# Integrating multi-ancestry common and rare variant mapping accelerates therapeutic target discovery

**DOI:** 10.64898/2026.06.25.26356239

**Authors:** Nobuyuki Enzan, Sean J. Jurgens, Seung Hoan Choi, Tetsushi Nakao, Satoshi Koyama, Patrick T. Ellinor

**Author notes:** **Correspondence** Satoshi Koyama, MD, PhD, Patrick T. Ellinor, MD, PhD. co-senior authors (Satoshi Koyama, Patrick T. Ellinor).

## Abstract

Integrating human genetics into therapeutic discovery accelerates drug development. However, ancestral biases in historical cohorts have left critical functional variation largely uncharted. Here, we leverage the diverse NIH *All of Us* Research Program to conduct comprehensive common- and rare-variant association analyses for 624 quantitative traits across 369,655 ancestrally diverse individuals. We identified 6,181 genome-wide significant locus-trait associations (526 novel) and 416 gene-trait associations (105 novel) via rare-variant burden testing. By integrating fine-mapping with computational variant-effect predictors, we systematically prioritized rare, likely causal variants driving these signals. Jointly modeling common and rare variation with protein-class annotations significantly improved the identification of known drug targets compared to common-variant analysis alone. Notably, we identified *NRG4* as a high-confidence candidate therapeutic target for preserving kidney function. Our findings demonstrate that characterization of rare and common variation across diverse populations enhances causal gene discovery and identifies novel, actionable therapeutic targets.

## Introduction

Genome-wide association studies (GWAS) and rare variant burden tests (RVAS or Rare Variant Association Studies) have identified thousands of loci/genes associated with human traits and diseases^1^. Although whole-genome sequencing (WGS) facilitates the comprehensive mapping of rare, trait-associated variants undetected by conventional arrays^2^, a critical gap remains: the vast majority of large-scale rare-variant analyses have been confined to populations of European descent^2^. Consequently, current genetic findings lack generalizability across diverse global populations^3^. Further, conventional GWAS have identified thousands of trait-associated loci, yet pinpointing the causal variants and genes underlying these associations remains challenging.

To address these limitations, we leverage the immense diversity of the *All of Us* Research Program (AoU) to perform both GWAS and RVAS for 624 quantitative traits across 369,655 participants from diverse ancestries, including individuals of African (AFR), Admixed American (AMR), and European (EUR) descent^4^. Using statistical fine-mapping together with enrichment analyses leveraging cis-regulatory element (CRE) annotations^5^ and multiple missense variant prediction tools^6–9^, we identified putatively causal and functional rare coding and noncoding variants associated with complex traits. Furthermore, we demonstrate that genes supported by both GWAS and RVAS signals show stronger links to drug development than genes identified by GWAS or RVAS alone, highlighting the value of integrating common and rare variant association analyses for the discovery of novel therapeutic targets.

## Results

### Comparison of Pooled and Meta-Analysis Frameworks in Diverse Cohorts

A defining feature of the AoU cohort is its substantial ancestral diversity, encompassing 77,584 AFR and 68,912 AMR participants. This diverse cohort allowed us to systematically evaluate the pooled multi-ancestry approach by comparing its performance against both a EUR-only GWAS and a traditional fixed-effects meta-analysis. We evaluated three distinct experimental designs: (1) 100% EUR; (2) a pooled analysis of ancestrally diverse individuals (composed of an equal 33%:33%:33% split of AFR, AMR, and EUR); and (3) a fixed-effects meta-analysis across these three groups. We observed no substantial differences in genomic inflation (λ_GC_) or statistical power among the three methods (**Supplementary Note 1**, **Methods**, **Supplementary Methods 1-5**, **Supplementary Figs. 1a-f**, and **Supplementary Data 1-3**).

We restricted our subsequent analyses to 624 phenotypes with data available for at least 1,000 individuals (**Methods** and **Supplementary Data 2**). Although the aggregate sample size of AoU is smaller than that of the UK Biobank (UKB), AoU provides greater statistical power for non-European populations due to its considerably larger AFR and AMR sample sizes (**Supplementary Fig. 2a**). Additionally, we confirmed that a pooled analysis inclusive of PCA outliers performed comparably to both pooled and meta-analyses that excluded them, validating the robustness of our approach (**Supplementary Note 2**, **Supplementary Figs. 2b-d**, **3a-f**, and **Supplementary Data 4**).

#### Multi-ancestry GWAS of 624 phenotypes yields 6,181 distinct phenotype-locus pairs

We performed GWAS for 624 phenotypes across six distinct genetic ancestry groups, as well as a comprehensive multi-ancestry pooled analysis leveraging the full study cohort (**Fig. 1**). Stringent quality control was performed as detailed in **Supplementary Note 3**, **Supplementary Figs. 4a-c**, and **Supplementary Data 5-7**. To assess the reliability of our findings, we cross-referenced our results with data from the UKB for overlapping phenotypes. In brief, 99.3% of the lead variants discovered in the AoU EUR cohort successfully replicated in the UKB EUR cohort (P < 0.05 with consistent effect directions). Notably, 95.8% of the lead variants identified in the smaller AoU AFR cohort were also successfully replicated within the UKB AFR population. Across all traits and ancestries, we identified a total of 9,871 distinct ancestry-phenotype-locus associations (**Fig. 2a** and **Supplementary Data 8**), corresponding to 6,181 unique phenotype-locus signals.

**Figure 1:**
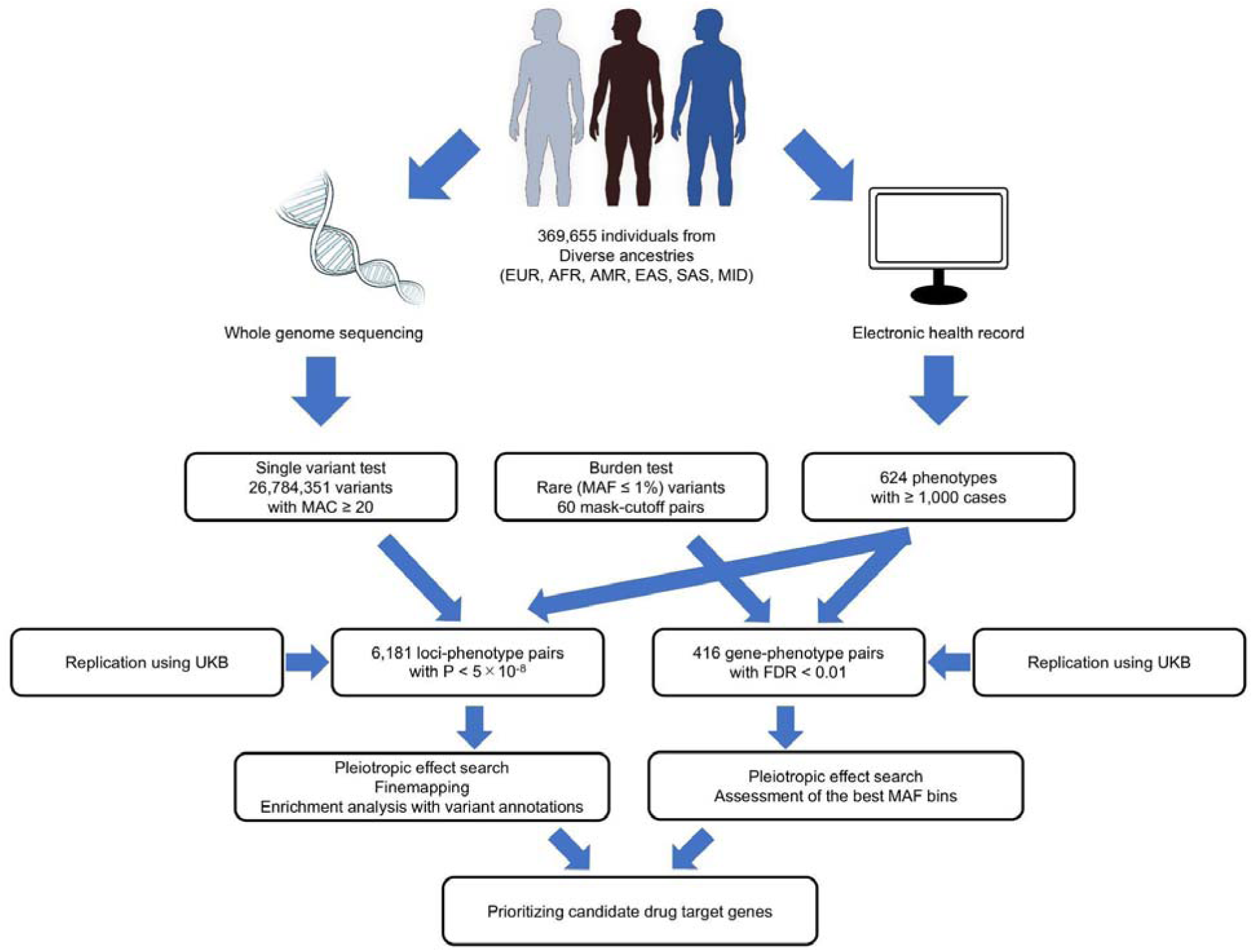
Study design. Flow diagram outlining the primary methodologies used for conducting the GWAS and RVAS. AFR, African American; AMR, Admixed American; EAS, East Asian; EUR, European; FDR, false discovery rate; MAC, minor allele count; MAF, minor allele frequency; MID, Middle Eastern; SAS, South Asian; UKB, the UK Biobank. This figure was created using Bioart (https://bioart.niaid.nih.gov/).

**Figure 2:**
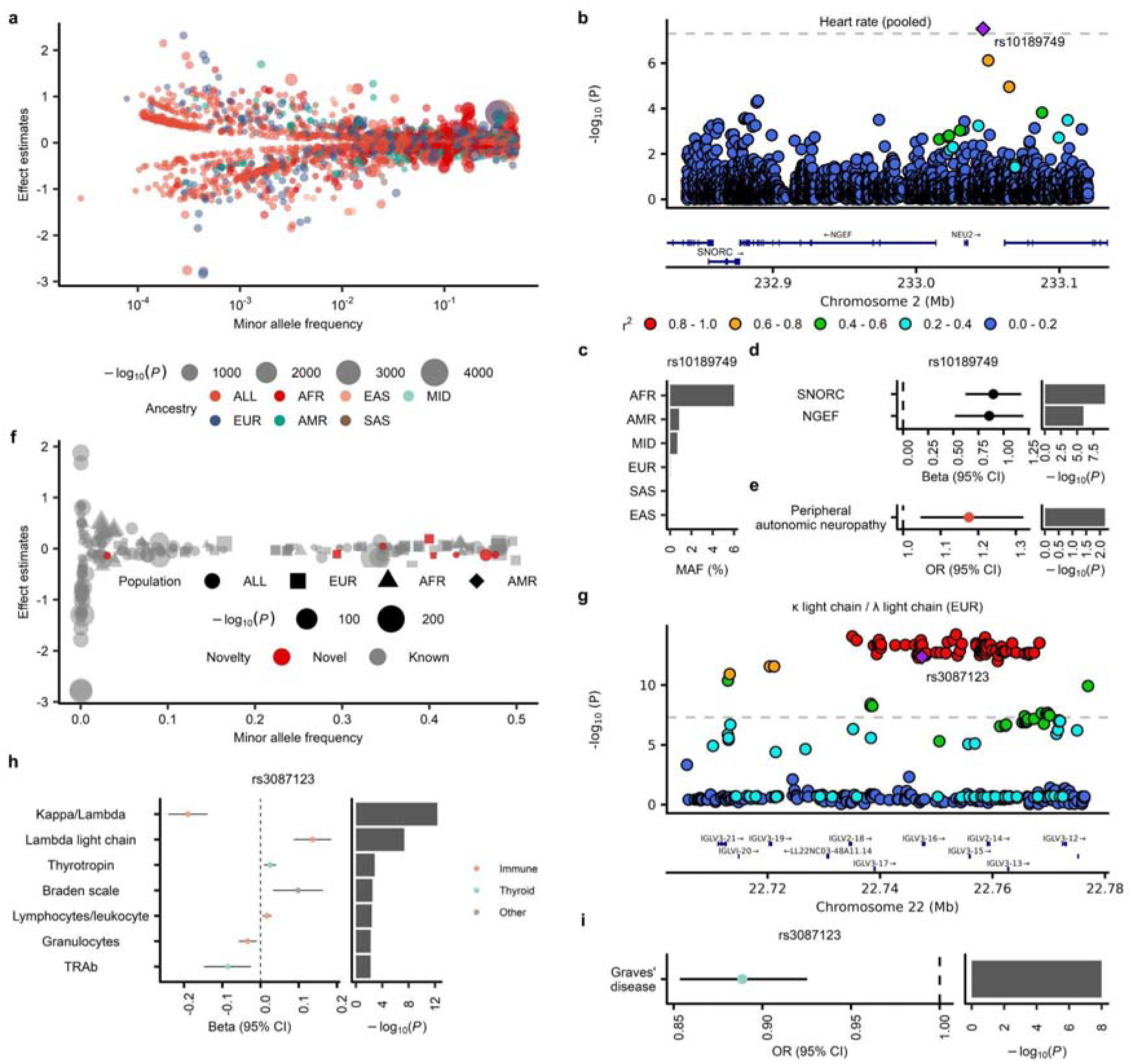
GWAS results for 624 quantitative traits. **a,** MAF and effect estimates for 9,871 ancestry–phenotype–lead SNPs. The x-axis shows the MAF, and the y-axis shows the effect estimates. **b**, Novel loci associated with heart rate. The purple square indicates the lead SNP rs10189749. **c,** MAF of rs10189749 in each ancestry, derived from gnomAD v4.1.1. **d,** The eQTL results from the GTEx project. The error bar indicates the 95% confidence intervals (CIs). **e,** The result from the Million Veterans Program (MVP). The error bar indicates the 95% CIs. **f,** MAF and effect estimates for 226 loss-of-function (LoF) variant-phenotype pairs (97 variants). The x-axis shows the MAF, and the y-axis shows the effect estimates. **g,** Locus plots for *IGLV3-16* locus. The purple square indicates the LoF variant rs3087123. The color scheme is the same as in **b**. **h,** Beta and P values for the top 7 QTL phenotypes associated with rs3087123 in AoU EUR. **i,** Top binary phenotype associated with rs3087123 in PLAtlas, along with its beta and P value. The color scheme is the same as in **h**. AFR, African American; AMR, Admixed American; AoU, All of Us; EAS, East Asian; eQTL, expression quantitative trait loci; EUR, European; MAF, minor allele frequency; MID, Middle Eastern; OR, odds ratio; SAS, South Asian; TRAb, Thyrotropin Receptor Antibody; SNP, single nucleotide polymorphism.

#### A multi-ancestry pooled approach uncovers rare variants in underrepresented populations

Out of 6,181 phenotype-locus pairs, 2,667 were discovered exclusively through the pooled approach (**Supplementary Fig. 4d**). A representative example of these pooled-specific findings is the AFR-specific *SNORC2* locus, which is significantly associated with heart rate (beta = 0.054; standard error [SE] = 0.010; P = 3.08 × 10^−8^; **Figs. 2b-c** and **Supplementary Data 9**). The effect estimate was consistent across ancestries and validated in an independent cohort^10^, MVP AFR (**Supplementary Fig. 5a** and **Supplementary Data 9**). Functionally, the lead variant, rs10189749, acts as an expression quantitative trait locus (eQTL) ^11^ for *SNORC* in the tibial nerve (beta = 0.90, SE = 0.14, P = 4.6 × 10^−10^; **Fig. 2d** and **Supplementary Data 10**). Given that *SNORC* is predominantly expressed in the nervous system (**Supplementary Fig. 5b**) and is linked to peripheral autonomic neuropathy (**Fig. 2e**), rs10189749 likely contributes to heart rate regulation via the autonomic nervous system.

#### Quantitative trait analysis identifies LoF variants elucidating disease mechanisms

Within the discovered locus-phenotype pairs, we prioritized 226 loss-of-function (LoF)-phenotype associations spanning 97 unique LoF variants (**Fig. 2f** and **Supplementary Data 11**). We highlighted rs3087123, a frameshift variant in *IGLV3-16*, which was significantly associated with a decreased kappa-to-lambda light chain ratio (beta = -0.19, SE = 0.03, P = 4.4 × 10^−13^) and increased lambda light chain levels (beta = 0.14, SE = 0.02, P = 4.8 × 10^−8^), denoting monoclonal lambda free light chain skewing (**Figs. 2g-h**, and **Supplementary Data 11**). The variant was additionally associated with thyrotropin and thyrotropin receptor antibody levels (**Fig. 2h and Supplementary Data 12**). Phenome-wide validation in PLAtlas^12^ confirmed that this allele protects against Graves’ disease (beta = -0.11, SE = 0.02, P = 1.0 × 10^−8^; **Fig. 2i**), pointing to a monoclonal lambda FLC-mediated protective mechanism driven by *IGLV3-16* disruption.

#### Functional characterization of causal noncoding variants

We conducted single- and multi-ancestry fine-mapping (**Supplementary Note 4**, **Supplementary Figs. 6a-c**, and **Supplementary Data 13**). Causal noncoding variants (posterior inclusion probability [PIP] > 0.9) were significantly enriched in promoters^5^ (promoter-like signature [PLS]: OR 4.47 [3.51 - 5.70], P = 1.1 × 10^−33^), proximal enhancers (proximal enhancer-like signature [pELS]: OR 2.17 [1.81 - 2.61], P = 1.5 × 10^−16^), and distal enhancers (distal enhancer-like signature [dELS]: OR 1.74 [1.54 - 1.96], P = 4.2 × 10^−20^) (**Fig. 3a**).

**Figure 3:**
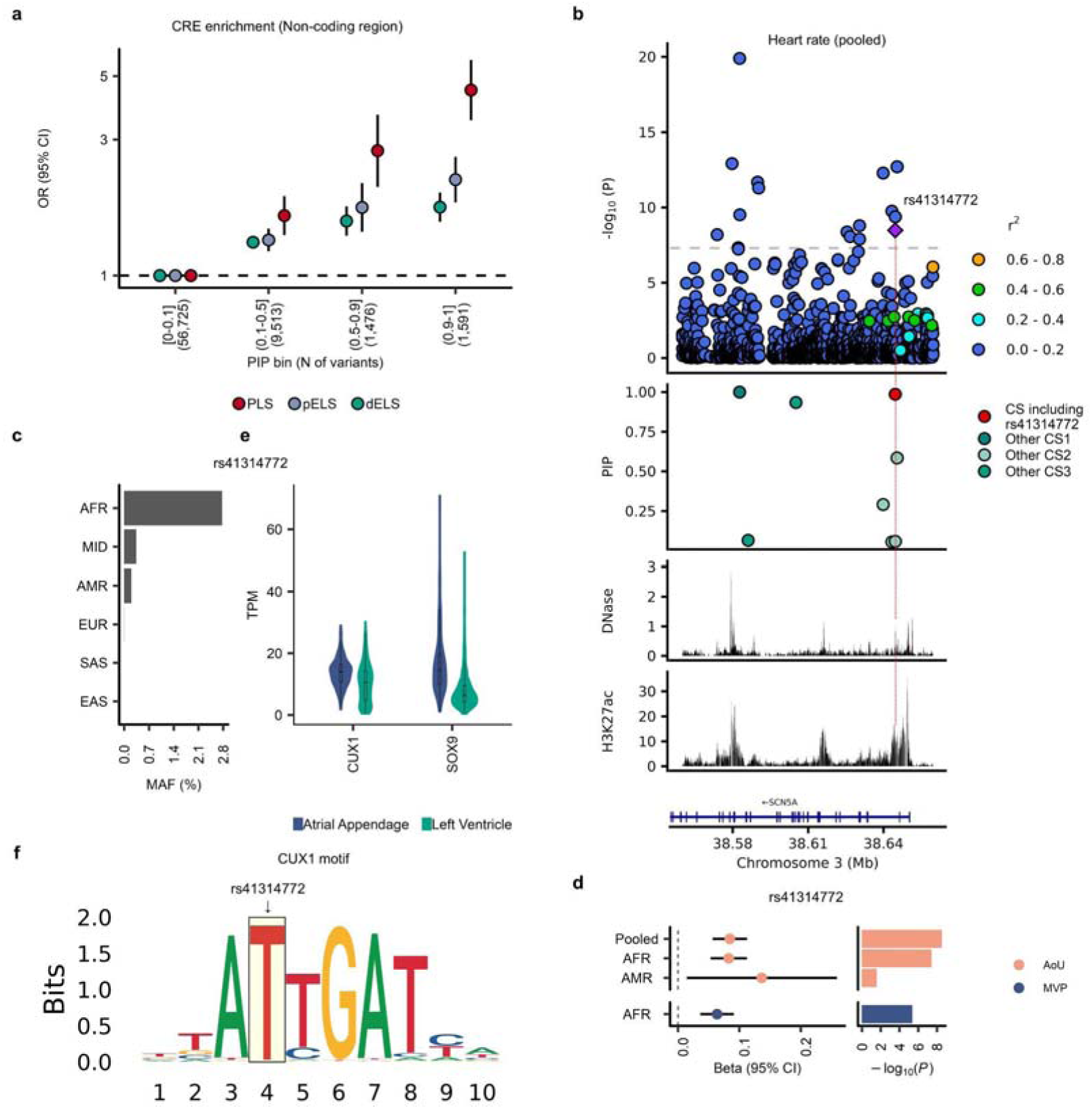
Likely causal variants are enriched in CRE. **a**, Enrichment of variants within the CRE across different PIP bins. The vertical axis indicates the OR of variants annotated as PLS, pELS, or dELS, compared to variants in the lowest PIP bin (0–0.1). The error bars indicate the 95% CIs. **b,** The locus plot, PIP, DNase peak and H3K27ac peaks for the *SCN5A* locus. The horizontal axis indicates the genomic coordinates. DNase (ENCSR214NLQ) and H3K27ac (ENCSR174GMQ) peaks are based on the right ventricle (**Supplementary Data 14**). **c,** The MAF of rs41314772 in each ancestry. The frequencies were derived from gnomAD v4.1.1. **d,** Effect estimates and P values of rs41314772. The error bar for the effect estimates indicates the 95% CI. **e,** Candidate transcription factors expressed in the atrial appendage or left ventricle. The vertical axis indicates TPM in each tissue. **f,** The motif of *CUX1*. The light-yellow shading indicates the position of rs41314772. This plot was based on the HOCOMOCO v13. AFR, African American; AMR, Admixed American; CI, confidence interval; CRE, cis-regulatory element; CS, credible set; dELS, distal enhancer-like signature; DNase, Deoxyribonuclease; EAS, East Asian; EUR, European; H3K27ac, Histone H3 Lysine 27 acetylation; MAF, minor allele frequency; MID, Middle Eastern; pELS, proximal enhancer-like signature; PIP, posterior inclusion probability; PLS, promoter-like signature; SAS, South Asian; TPM, Transcripts Per Million.

A notable example is rs41314772, a novel, AFR-specific noncoding variant in an *SCN5A* intron associated with heart rate. This variant resides within a cardiac muscle-specific dELS (**Fig. 3b**, **Supplementary Fig. 7a**, and **Supplementary Data 14**) and is more frequent in AFR (minor allele frequency [MAF] = 2.77%) than in EUR populations (MAF = 0.003%) (**Fig. 3c**). This variant acts independently of the lead variant rs3922844 (r^2^ = 0.008), exhibiting distinct credible sets (**Fig. 3b**). Effect sizes for rs41314772 were consistent across ancestries as well as in the independent MVP AFR cohort (**Fig. 3d**, **Supplementary Data 15**). In MVP, this variant was associated with atrial fibrillation (OR 1.22 [1.11 - 1.35]), paroxysmal ventricular tachycardia (OR 1.26 [1.03 - 1.54]), and cardiomyopathy (OR 1.12 [1.01 - 1.26]) (**Supplementary Fig. 7b** and **Supplementary Data 16**). Mechanistically, motifbreakR analysis indicated that rs41314772 disrupts 12 transcription factor binding motifs (**Supplementary Data 17**). Two of these, *CUX1* and *SOX9*, are expressed in the left atrial appendage and left ventricle (**Fig. 3e**). However, multiple lines of evidence and a larger allelic effect size suggest that *CUX1* is likely the primary binding motif disrupted at this locus (**Fig. 3f**).

### Multiple prediction tools identify likely causal missense variants

Within coding regions, high-PIP variants were enriched for damaging missense variants supported by at least one prediction tool (OR 11.6 [7.4 - 18.2], P = 1.3 × 10^−26^; **Fig. 4a**). A significant, albeit weaker, enrichment was also observed among “non-damaging” missense variants lacking support from any prediction tools (**Fig. 4a**). A representative causal damaging variant is EUR-specific rs145611185 in *MYH6*, which is associated with heart rate (beta = -0.30; SE = 0.05; P = 6.4 × 10^−9^; **Fig. 4b**, **Supplementary Figs. 8a-b,** and **Supplementary Data 18**). In UKB, rs145611185 was associated with bradycardia (OR = 2.64 [1.93 - 3.62]) and sick sinus syndrome (OR 4.94 [2.77 - 8.81]) (**Supplementary Fig. 8c** and **Supplementary Data 19**). Furthermore, the extensive WGS data for AMR participants in the AoU dataset enabled the detection of AMR-specific rare variant-phenotype associations (**Supplementary Figs. 9-10, Supplementary Data 20-22**, **Supplementary Notes 5-6)**.

**Figure 4:**
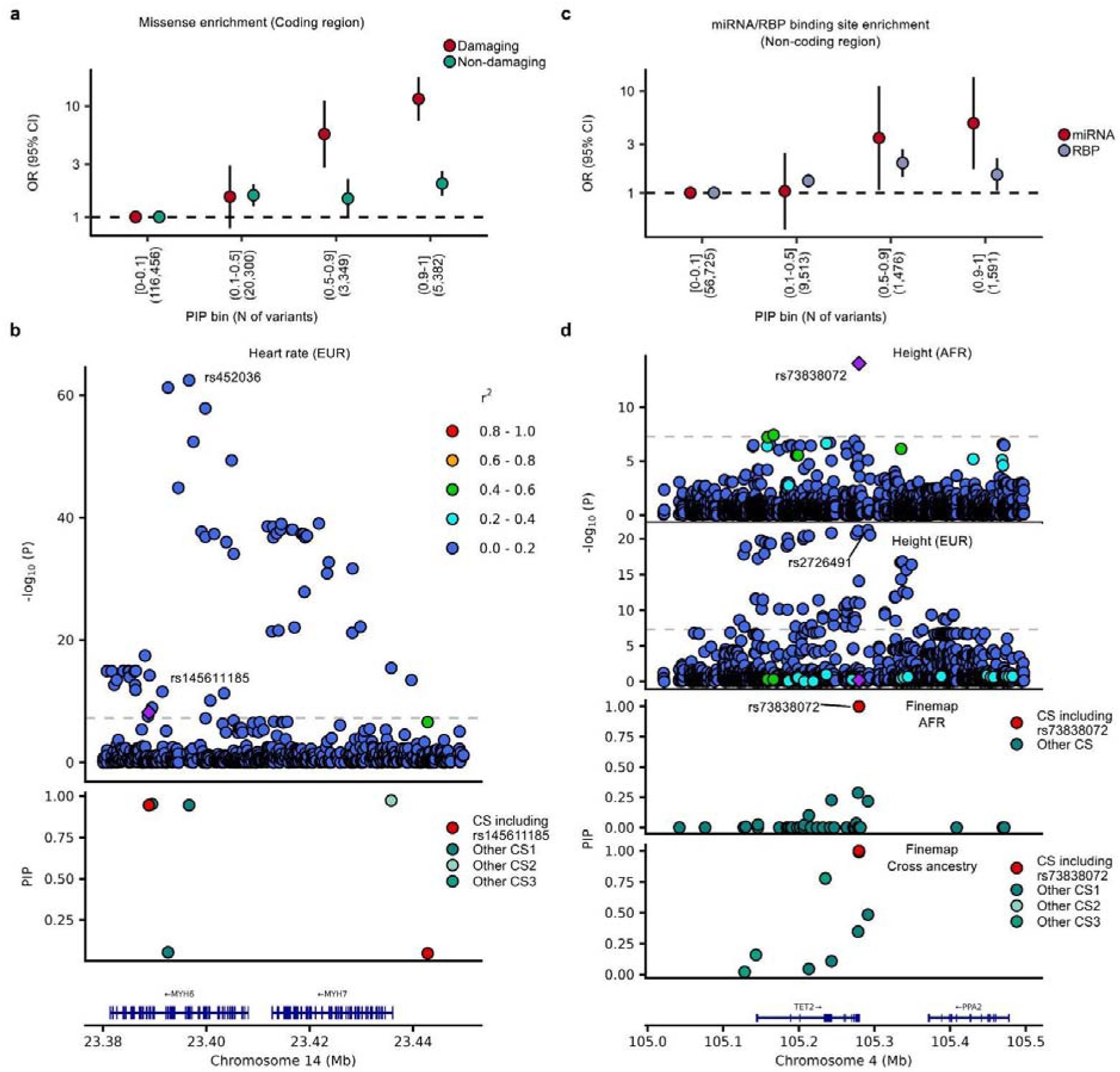
Likely causal variants are enriched in damaging missense and miRNA binding sites. **a**, Enrichment of damaging and non-damaging missense variants across different PIP bins. The vertical axis indicates the OR for variants within each PIP bin that are classified as damaging or non-damaging missense variants, relative to variants in the lowest PIP bin (0–0.1). The error bars indicate the 95% CIs. **b,** The locus plot and PIP for the *MYH6* locus. The horizontal axis indicates the genomic coordinates. **c**, Enrichment of variants within the binding sites of miRNA/RBP in variants with high PIP. The vertical axis indicates the OR for variants within each PIP bin located within miRNA or RBP binding sites, relative to variants in the lowest PIP bin (0–0.1). **d,** The locus plot and PIP for the *TET2* locus. The horizontal axis indicates the genomic coordinates. AFR, African American; CS, credible set; EUR, European; miRNA, MicroRNA; PIP, posterior inclusion probability; RBP, RNA-binding protein.

### Higher PIP variants are enriched in miRNA binding sites

Noncoding causal variants showed significant enrichment at the binding sites of microRNAs (miRNAs) and RNA-binding proteins (RBPs) (**Fig. 4c**). Specifically, among variants with a high PIP (> 0.5), we identified 11 variant-miRNA pairs located within miRNA binding sites (**Supplementary Data 23**). A representative example is rs73838072, a rare, AFR-specific variant associated with body height (MAF: 3.79% in AFR, 0.03% in EUR, and 0.81% in AMR; **Supplementary Figs. 11a-b** and **Supplementary Data 24**). rs73838072 is located in the 3’-UTR of *TET2* (**Fig. 4d**). Mechanistically, rs73838072 causes a mismatch in the seed region of *miR-210-3p* (**Supplementary Fig. 11c**). These findings highlight that miRNA binding sites should be systematically evaluated in the downstream analysis of GWAS results.

#### RVAS identifies 416 significant gene-phenotype pairs across 624 traits

We conducted RVAS across 624 quantitative traits using all available samples. By analyzing up to 60 distinct rare variant masks and combining their *P*-values via the Cauchy distribution test^13^ (**Methods**, **Supplementary Methods 6**, **Supplementary Fig. 12a**), we identified 416 significant gene-phenotype pairs in the AoU dataset (FDR < 0.01; **Fig. 5a**, **Supplementary Data 25**). Comprehensive quality checks of these results are detailed in the supplement (**Supplementary Notes 7**, **Supplementary Figs. 12b-f**, **Supplementary Data 26-30**). In brief, among the 72 phenotypes (out of a total of 106) previously analyzed by RVAS, 361 of the 363 significant gene-phenotype pairs (99.4%) were already reported in the Open Targets Platform or GeneBass. This indicates a high consistency with previous reports and confirms the validity of our approach.

**Figure 5:**
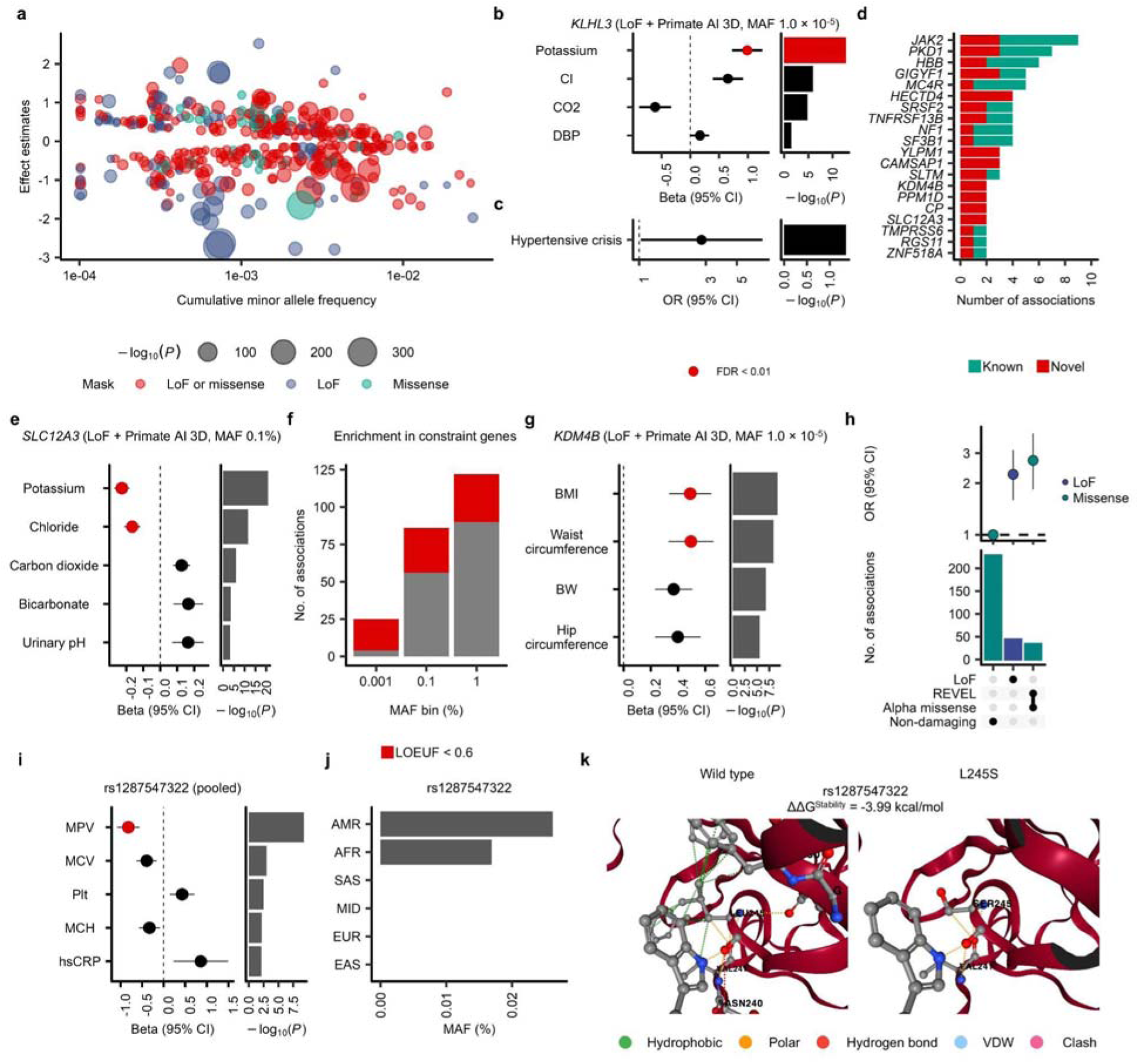
The rare variant analyses in exon regions revealed novel associations. **a**, Effect estimates plotted against cumulative MAF for 416 gene–phenotype pairs. **b, c,** Effect estimates and P values of *KLHL3* with the best mask for electrolytes-related traits (**b**; quantitative traits) and hypertension-related traits (**c;** binary traits) withing the AoU cohort. The error bars for the effect estimates indicate 95% confidence intervals (CIs) in **b**, **c**, **e**, **g**, **h**, and **i**. **d,** Number of significantly associated phenotypes per gene, showing genes with at least two associations. **e,** Effect estimates and P values of *SLC12A3* with the best mask on electrolytes- and acid-base balance-related traits within AoU. **f,** Number of significant genes classified by their best mask MAF thresholds (1%, 0.1%, or 0.001%). Constraint genes are defined by a LOEUF < 0.6. **g,** Effect estimates and P values of *KDM4B* with the best mask for obesity-related traits within AoU. **h,** Enrichment of LoF and damaging missense variants among significant variants with MAF < 1% in exon regions. The top panel shows the enrichment of indicated variant classes relative to non-damaging missense variants. The bottom panel shows the number of significant variants in the indicated variant classes. Only LoF, REVEL, Alpha missense, and non-damaging missense variants are shown (full data available in **Supplementary Fig.14a**). **i,** Effect estimates and P values of rs1287547322 in the pooled analysis. **j,** MAF of rs1287547322 across different ancestries, derived from gnomAD v4.1.1. **k,** Schematic representation of the *VNN1* variant. The left panel shows the wild-type structure, while the right panel illustrates how the rs1287547322 (L245S) mutation alters interactions with the surrounding amino acids. The figures were generated using DynaMut2. BMI, body mass index; BW, body weight; CI, confidence interval; hsCRP, high-sensitivity C-reactive protein; LOEUF, Loss-of-Function Observed/Expected Upper Bound Fraction; LoF, loss-of-funciton; MAF, minor allele frequency; MCH, mean corpuscular hemoglobin; MCV, mean corpuscular volume; MPV, mean platelet volume; OR, odds ratio; Plt, platelet; VDW, van der Waals.

#### Quantitative trait analysis identifies subclinical forms of monogenic disease

A notable finding from our RVAS was the association between *KLHL3* and potassium levels (**Fig. 5b**, **Supplementary Data 31**). *KLHL3* regulates the sodium-chloride cotransporter in the kidney and is a known causal gene for pseudohypoaldosteronism type II (PHAII). Clinically, PHAII is characterized by hyperkalemia, hyperchloremic metabolic acidosis, and hypertension, typically requiring thiazide diuretics for management^14^. Consistent with these established clinical features, our analysis showed that *KLHL3* variants were associated with increased blood potassium and chloride, alongside decreased carbon dioxide (**Fig. 5b**, **Supplementary Figs. 13c-d**, and **Supplementary Data 31**). While the exact prevalence of PHAII in the general population remains unknown, with only about 180 affected individuals and families reported to date, our results indicate that a less severe form of the condition is much more common than previously recognized (cumulative MAF [cMAF] = 0.02%). Importantly, despite its milder presentation, individuals carrying these variants remain at a significantly elevated risk for hypertensive crisis (OR 2.79 [1.03 - 7.61], P = 0.044; **Fig. 5c** and **Supplementary Data 31**).

#### Pleiotropic associations highlight genes of clinical significance

Of the 217 genes associated with at least one phenotype, 20 were associated with two or more (**Fig. 5d** and **Supplementary Data 25**). As expected, *JAK2* was associated with multiple blood cell-related traits, and *PKD1* with kidney-related traits. Our study identified novel associations between *SLC12A3* and two phenotypes: potassium and chloride levels (**Fig. 5e**, **Supplementary Fig. 13e**, and **Supplementary Data 25, 32**). *SLC12A3* is a protein-coding gene that regulates the thiazide-sensitive sodium-chloride cotransporter (NCC) and is a causal gene for Gitelman syndrome^15^. In our RVAS, *SLC12A3* was also associated with increased carbon dioxide, bicarbonate, and urinary pH (**Fig. 5e** and **Supplementary Data 32**), which was consistent with metabolic alkalosis. Given the 0.6% cMAF of *SLC12A3*, a less severe form of Gitelman syndrome might be relatively prevalent.

#### Lower MAF cutoffs yield stronger association signals for constrained genes

The statistical power of RVAS depends on the masking strategy. Hence, we sought to determine the optimal MAF cutoff for each gene. We defined the optimal MAF bin as the one yielding the lowest P-value for a given gene. To ensure robust comparisons between the < 0.001% MAF bin and other bins, we restricted this analysis to phenotypes with a sample size > 100,000, resulting in a total of 34 phenotypes and 176 genes.

Among the 176 genes, the lowest P-values were observed in the 1% MAF bin for 122 genes, the 0.1% bin for 86 genes, the 0.001% bin for 25 genes, and the singleton bin for 5 genes (the sum exceeds 176 due to ties across multiple bins) (**Fig. 5f** and **Supplementary Data 33**). Notably, the 0.001% MAF bin yielded the lowest P-values for one-third of the constrained genes, defined as having a Loss-of-Function Observed/Expected Upper Bound Fraction (LOEUF) < 0.6. This suggests that the optimal MAF bin for detecting gene-phenotype associations may vary depending on the level of gene constraint.

For example, using the 0.001% MAF bin, we identified an association between *KDM4B* (LOEUF = 0.371, the top decile among all protein-coding genes) and both higher BMI and increased waist circumference (**Fig. 5g** and **Supplementary Data 34**). Carriers of this *KDM4B* mask were at higher risk for pulmonary embolism, asthma, gallstones, and ischemic heart disease (**Supplementary Fig. 13f** and **Supplementary Data 34**), all of which are conditions with obesity as a known risk factor. These associations were successfully identified by utilizing multiple MAF bins and integrating the results via a Cauchy distribution.

#### Rare variant associations are driven by damaging missense variants

Advances in the scalability of WGS have enabled single-variant analyses even for rare variants. We performed single-variant association tests (minor allele count [MAC] > 20) across coding regions and identified 634 significant variant-phenotype pairs (P < 5 × 10^−8^) involving 364 unique variants (**Supplementary Data 35**). Of these, 44 were LoF variants and 320 were missense variants (**Supplementary Fig. 14a**). As expected, the significant rare variants were enriched for LoF variants (OR 2.25 [1.59 – 3.13], P = 7.4 × 10^−6^). Among missense variants, the significant hits were also enriched for those predicted to be damaging by AlphaMissense^6^ + REVEL^8^ with or without PrimateAI-3D^7^ (**Supplementary Fig. 14a** and **Supplementary Data 36**). Overall, exome-wide significant variants were more strongly enriched for missense variants supported by AlphaMissense and REVEL (compared to non-damaging missense variants; OR 2.72 [1.84 – 3.91], P = 1.1 × 10^−6^) than for LoF variants (**Fig. 5h** and **Supplementary Data 37**).

A notable example is rs1287547322, a missense variant in *VNN1* supported by both AlphaMissense and REVEL (**Supplementary Fig. 14b** and **Supplementary Data 35**), which was associated with decreased mean platelet volume (beta = -0.82, SE = 0.13; P = 5.0 × 10^−10^) (**Fig. 5i** and **Supplementary Data 38**). This variant is highly specific to the AMR and AFR populations (MAF: 0.026% and 0.017%, respectively, and absent in other ancestries) (**Fig. 5j**) and destabilizes the protein structure through the loss of hydrophobic bonds (**Fig. 5k**). Furthermore, rs1287547322 was associated with an increased platelet count and microcytic anemia (**Fig. 5i**, **Supplementary Fig. 14c**, and **Supplementary Data 38**). Given that *VNN1* is a known regulator of inflammation^17^ and rs1287547322 was also associated with elevated high-sensitivity CRP (hsCRP) (**Fig. 5i** and **Supplementary Data 38**), this variant may contribute to chronic inflammation, potentially explaining the observed association with thrombocytosis, microcytic anemia, and increased susceptibility to COVID-19 (**Supplementary Fig. 14c** and **Supplementary Data 38**).

#### GWAS and RVAS evidence additively predict approved drug targets

Next, we sought to identify candidate drug target genes using GWAS and RVAS results. We identified 2,785 genes based on GWAS proximity (nearest genes) or RVAS results. Of these, 2,568 were supported solely by GWAS, 140 by both GWAS and RVAS, and 77 exclusively by RVAS (**Fig. 6a** and **Supplementary Data 39**). We used the inverse rank-normalized LOEUF as a covariate in subsequent analyses (**Supplementary Note 8**, **Supplementary Fig. 15a**, and **Supplementary Data 40**). Genes supported by either GWAS or RVAS were significantly enriched for targets of phase 3 clinical trials and approved drugs (GWAS: OR 2.00 [1.66 - 2.42]; RVAS: OR 5.87 [3.98 - 8.66]), independent of LOEUF (**Fig. 6b** and **Supplementary Data 41**). Notably, genes supported by both GWAS and RVAS demonstrated stronger enrichment for approved drug targets compared to those supported by either method alone (**Fig. 6c** and **Supplementary Data 41**). Certain protein classes are known to possess higher druggability (**Supplementary Note 9**, **Supplementary Fig. 15b**, and **Supplementary Data 42-43**). We performed multivariable logistic regression to determine whether the enrichment of GWAS and RVAS genes among approved drug targets was independent of protein class and gene constraint. Both GWAS genes (OR 1.46 [1.16 - 1.84], P = 1.25 × 10^−3^) and RVAS genes (OR 2.09 [1.29 - 3.39], P = 2.66 × 10^−3^) were independently and additively associated with approved drug targets (**Fig. 6d** and **Supplementary Data 44**). Thus, integrating protein class information with GWAS and RVAS evidence enables more accurate identification of candidate drug targets. A prominent example is the novel RVAS association between *NRG4* (Neuregulin 4) and blood urea nitrogen (BUN) levels, driven by LoF and any missense predictors (beta = 0.62, SE = 0.11, P = 3.1 × 10^−9^; **Fig. 6e**). *NRG4* encodes a brown adipose tissue-secreted adipokine with beneficial metabolic effects in mice, and circulating NRG4 is independently associated with preserved renal function. The RVAS results for BUN and creatinine (Cre) were consistent across the AoU and UKB cohorts (**Supplementary Fig. 15c** and **Supplementary Data 45**). Within our framework, *NRG4* is classified as encoding a secreted protein and is supported by both GWAS and RVAS evidence. These findings suggest that *NRG4* may represent a potential therapeutic target for chronic kidney disease.

**Figure 6:**
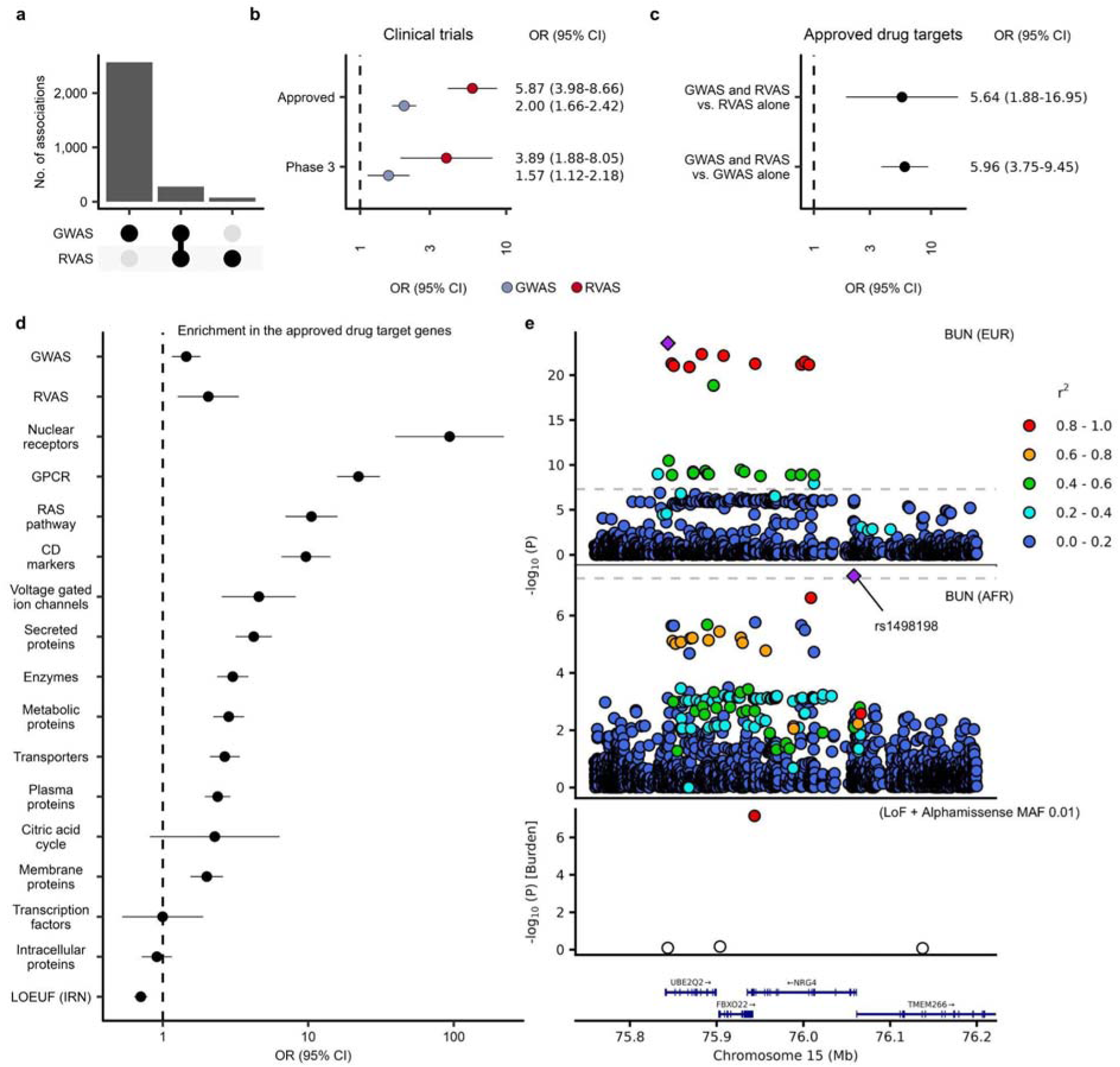
GWAS and RVAS genes additively contribute to drug discovery independent of protein classes. **a**, Number of genes identified as the nearest genes in GWAS, RVAS, and both methods. **b,** Enrichment of GWAS and RVAS genes among Phase 3 clinical trial targets and approved drug targets. **c,** Enrichment of genes identified by both GWAS and RVAS, compared to those identified by GWAS alone or RVAS alone, among approved drug targets. **d,** Multivariable logistic regression analysis for approved drug targets. All variables included in the model are displayed. For **b**, **c**, and **d**, inverse-rank-normalized LOEUF was included as a covariate, and error bars indicate 95% CIs. **e,** Representative example of a secreted protein supported by both GWAS and RVAS. The top two panels show the GWAS results for blood urea nitrogen (BUN) in EUR and AFR, respectively. The middle panel shows the corresponding RVAS results. P values for the RVAS were derived from the best mask (LoF + Missense, MAF < 1%). AFR, African American; CD, cluster of differentiation; CI, confidence interval; EUR, European; GPCR, G protein-coupled receptor; GWAS, genome-wide association study; IRN, inverse rank normalization; LOEUF, Loss-of-Function Observed/Expected Upper Bound Fraction; MAF, minor allele frequency; OR, odds ratio; RVAS, rare variant association study.

## Discussion

In this study, we performed multi-trait genetic analyses across diverse ancestries, including substantial representations of AFR and AMR populations. Our *in silico* functional analyses facilitated the identification of population-specific causal variants, highlighting the critical role of ancestral diversity in genetic discovery. Notably, beyond well-studied coding variants and CREs, we demonstrated that the disruption of miRNA binding sites represents an additional mechanism underlying causal noncoding associations. For RVAS, our comprehensive framework facilitated the discovery of novel gene-phenotype associations, demonstrating that the optimal MAF cutoff for variant masking varies depending on the degree of gene constraint. Finally, we established a systematic strategy for prioritizing potential therapeutic targets by integrating evidence from GWAS, RVAS, and protein functional class annotations.

Previous quantitative trait locus (QTL) studies have largely been limited to anthropometric traits and common laboratory measurements, such as blood cell counts and lipid levels. In contrast, the AoU cohort enables the utilization of rich phenotypic data extracted from electronic health records (EHRs), facilitating the investigation of biological mechanisms underlying complex diseases (**Fig. 2g**).

As sample sizes from non-European populations continue to increase, the optimal strategy for conducting multi-ancestry analyses remains a subject of ongoing discussion. While the validity of the pooled approach has been examined in previous studies, we also observed no substantial inflation of λ_GC_ in our pooled analysis compared with conventional meta-analysis (**Supplementary Fig. 1e**). Furthermore, the ancestry-specific novel variants identified through our pooled approach were thoroughly validated by multi-ancestry fine-mapping, the latest CRE datasets, and replication analysis in the independent cohorts, supporting the robustness of these findings. Consequently, these lines of evidence suggest that the pooled approach serves as a viable and robust strategy for the genetic analysis of ancestrally diverse cohorts.

While multiple *in silico* prediction tools are available to identify damaging missense variants, a consensus regarding their optimal utilization remains to be established. Nevertheless, our empirical observations suggest that deleterious missense variants supported by any of these prediction tools are more likely to be causal, which is consistent with our annotation-agnostic fine-mapping results. Conversely, high-PIP variants were also enriched among “non-damaging” missense variants (**Fig. 4a**), suggesting that certain functionally relevant missense variants may not be captured by the predictors employed in this study. For instance, current tools generally cannot predict gain-of-function variants. These findings highlight both the utility of combining fine-mapping with multiple missense predictors to identify candidate damaging variants, and the ongoing need for the development of more accurate predictive frameworks.

We demonstrated that, in addition to CREs, miRNA and RBP binding sites represent potential mechanisms underlying genome-wide significant variants in noncoding regions (**Fig. 4c**). Previous research has similarly shown that, apart from CREs and missense variants, likely causal variants are frequently located within the 3’-UTR, where miRNA binding sites are predominantly clustered. Taken together, these findings highlight that miRNA binding sites should be systematically evaluated alongside CREs in the downstream analysis of GWAS results.

In RVAS, the choice of appropriate MAF cutoffs and missense variant predictors is critical; however, the optimal cutoff and predictor vary by gene and cannot be uniformly defined. We demonstrated that for genes under stronger constraint, utilizing a lower MAF cutoff improves statistical power (**Fig. 5f**), as illustrated by the association between *KDM4B* and obesity (**Fig. 5g**). A potential explanation for this trend is that high-MAF missense variants in highly constrained genes may be more prone to false-positive deleterious annotations, whereas high-MAF missense variants in less constrained genes may represent genuinely functional alleles, thereby leading to distinct optimal MAF cutoffs. Furthermore, by integrating multiple variant masks, MAF cutoffs, and the Cauchy combination test, our framework systematically evaluated a wide range of analytical criteria, enabling the identification of novel gene–phenotype pairs that had previously been overlooked (**Supplementary Figs. 13a-b**).Finally, we investigated the utility of GWAS and RVAS evidence in prioritizing candidate drug targets. While previous studies have shown that genes identified through GWAS possess higher odds of becoming successful drug targets, our findings demonstrate that genes identified via RVAS similarly exhibit significantly elevated odds for therapeutic development. Importantly, GWAS and RVAS evidence contributed to drug target identification additively and independently of protein functional classes and gene constraint. These results suggest that integrating information on druggable protein classes with both GWAS and RVAS findings facilitates the more efficient and accurate identification of novel therapeutic candidates.

## Online Methods

### Study datasets

The AoU research program of the National Institutes of Health (NIH) is a longitudinal cohort study that aims to include 1 million racially, ancestrally and demographically diverse participants from across the USA, combining phenotypic data from various sources including patient-derived information and the EHR linkage^22^. As part of the release in February 2025, WGS was performed on 414,830 participants using Illumina NovaSeq 6000 machines following manufacturer’s best practices. The same protocol for library preparation (PCR Free Kapa HyperPrep) and software for variant calling (DRAGEN v.3.7.8) were used to keep consistent WGS data generated from different AoU Genome Centers. Detailed descriptions of the AoU data and the central quality control (QC) is described in the **Supplementary Methods 1**.

We performed further genotype, variant and sample QC procedures on both the Allele Count/Allele Frequency (ACAF) call set (MAF > 1% or population-specific MAC > 100) and the exome-region call set (contains variants that are within the exon regions of the Gencode v.42 basic transcripts, with padding of 15 bases on either side of each exon) released by the program, resulting in 410,400 eligible samples and 26,784,351 high-quality genetic variants for the ACAF call set. Details on the QC procedure, ancestry inference, principal component analysis and relatedness inference are described in the **Supplementary Methods 1**. All enrolled participants provided informed consent to AoU. Use of AoU data was approved under a data use agreement between the Massachusetts General Hospital and the AoU research program. We took the same approach for sample/variant QC of UKB (**Supplementary Methods 2**).

### Ancestry definitions

In all analyses, ancestry labels were based on inference from the genetic data. We defined labels for continental ancestries, namely EUR, AFR, AMR, EAS, SAS, and MID ancestries. Methodology for ancestry inference is described in the **Supplementary Methods 3**.

### Phenotype construction

We assessed all measurement phenotypes with more than 1,000 individuals. For each measurement, we selected the most frequently used unit. Ethnic groups with fewer than 200 individuals measured were excluded. When measurements were reported in multiple units (e.g., mg/dL and g/L), the most commonly used unit was adopted. To avoid potential confounding, we excluded measurements obtained in emergency, critical care, or inpatient settings; the excluded clinical settings are listed in **Supplementary Data 1**. We used both EHR data and participant-provided information for physical measurements. These included blood pressure, heart rate, height, weight, body mass index (BMI), and waist and hip circumference. For other measurements, we restricted the analysis to participants who had provided consent for the use of their EHR data.　The curation of Fitbit data and the mind datasets is described in **Supplementary Methods 4**. For each phenotype, values outside the median ± 6 MAD were excluded as outliers. For phenotypes with multiple measurements, the median value was used. We then regressed out age and sex, followed by inverse rank-based normalization within each ancestry group and in the combined all-ancestry sample. For the X chromosome analysis, GWAS was conducted separately in males and females followed by fixed-effect meta-analysis. Accordingly, for each phenotype, age was regressed out separately by sex, and inverse rank-based normalization was then applied within each ancestry group and in the combined all-ancestry sample. Phenotypes with a male proportion > 0.9 were defined as male-specific, whereas those with a male proportion < 0.1 were defined as female-specific phenotypes. Female participants were excluded from male-specific phenotypes, and male participants were excluded from female-specific phenotypes. The method to generate simulation datasets are described in **Supplementary Methods 5**.

### GWAS

GWAS was performed by Regenie v4.1^23^ with adjustment for age, age^2^, sex, the genotype center, the sample source (blood or saliva) (**Supplementary Methods 7**), and the top 20 PCAs. We selected variants with MAC > 20 in each phenotype. Of note, we used ACAF call set for GWAS, so the minimal MAF was 1% or population-specific MAC > 100 in the entire dataset. The genome-wide significance threshold was defined at P < 5.0 × 10^−8^. Adjacent genome-wide significant SNPs were grouped into one locus if they were within 1 Mb of each other. We defined a locus as follows: (1) extracted genome-wide significant variants (P < 5.0 × 10^−8^) from the association result, (2) added 500 kb to both sides of these variants, and (3) merged overlapping regions. If the locus did not contain coordinates with previously reported genome-wide significant variants (**Supplementary Methods 8-9**), the region was annotated as being novel.

### Pooled analysis and meta-analysis

For pooled analysis, a GWAS was performed on the entire dataset by combining all ancestry groups into a single analysis. Population structure was adjusted using the top 20 cross-ancestry PCAs derived from the full pooled dataset (**Supplementary Methods 1**), along with covariate adjustments as described above. For meta-analysis, a GWAS was conducted separately within each ancestry group. Top 20 ancestry-group-specific PCAs were used for population structure adjustment, along with the same covariates. Summary statistics from individual ancestry analyses were then combined using a fixed-effect meta-analysis approach with inverse-variance weighting.

### LDSC and heritability

To estimate confounding biases such as cryptic relatedness and population stratification, we performed ldscr, which was based on Linkage disequilibrium score regression (LDSC)^24^. We selected SNPs with MAF ≥ 0.01, and variants within the major histocompatibility complex region were excluded. For the regression, we used the LD scores provided by the Pan-UK Biobank.

### PheWAS analysis for novel variants found in our GWAS

For PheWAS analysis, we used several datasets. PLAtlas is the largest multi-ancestry meta-analysis for PheWAS studies^12^ (**Fig. 2i** and **Supplementary Fig. 11b**). Since PLAtlas is focused on low-frequency to common variants (MAF > 0.1%), AFR-specific or AMR-specific rare variants cannot be tested in PLAtlas. For these rare variants not listed in PLAtlas, we used MVP PheWAS results^10^ (**Fig. 2e**, **Fig. 3d**, **Supplementary Fig. 5a**, **Supplementary Fig. 7b**, and **Supplementary Figs. 9c-d**) and also we ran PheWAS using UKB WGS datasets (**Supplementary Methods 1**, **Supplementary Methods 10** and **Supplementary Fig. 8c**). Also, we checked the T2DKP portal for AMR-specific rare variants (**Supplementary Fig. 10d**).

### Fine-mapping

To identify causal variants within genome-wide significant loci, we used fine-mapping analyses with SuSiEx^25^ using an in-sample LD matrix. We generated 95% credible sets and calculated PIPs for both cross-ancestry and single-population analyses. We assigned to each variant the highest PIP across phenotypes and used this value for the enrichment analysis. Based on these highest PIP, variants were categorized into four groups: PIP bin [0 - 0.1], (0.1 - 0.5], (0.5 - 0.9] and (0.9 - 1.0)^21^.

### Cis-regulatory elements

We downloaded CRE, namely PLS (promoter-like signature), pELS (proximal enhancer-like signature) and dELS (distal enhancer-like signature), from SCREEN^5^. For variants in non-coding regions, we counted the overlap of variants with each CRE and calculated the OR of variants in each PPI bin to the lowest PPI bin. We tested the OR using a Fisher’s exact test.

### Motif differential affinity analysis

The motifBreakR^26^ tested the potential of each variant to disrupt binding affinity of any motif with the method = ‘ic’. The background probabilities of each nucleotide were set as A = C = G = T = 0.25. To identify relevant transcription factors, we utilized gene expression from GTEx v10.

### Variant annotation

We annotated genetic variants using the Variant Effect Predictor (VEP)^27^ with the LOFTEE plugin^28^, as well as the dbNSFP database^29^ and various additional prediction tools (**Supplementary Methods 6**). Our annotation pipeline first used VEP to identify rare coding variants affecting the ENSEMBL canonical transcripts of protein-coding genes, and then extracted missense variants (annotated as such by VEP) and high-confidence LoF (annotated as such by LOFTEE). Among LoFs, we distinguished two groups, including one irrespective of potential LOFTEE flags, and a more stringent group excluding LoFs with any LOFTEE flags^28^ (PTVnoflag; Supplementary Note). Among missense variants, various bioinformatic prediction tools were utilized to identify groups of variants with potentially damaging effects, namely PrimateAI-3D^7^, REVEL^8^, popEVE^9^, and AlphaMissense^6^ (**Supplementary Methods 6**).

### Enrichment analysis of likely causal variants in coding regions

We defined variants supported by at least one missense predictor (PrimateAI-3D, REVEL, popEVE, and AlphaMissense) as damaging missense, and those not supported by any of these predictors as non-damaging missense variants. We counted the overlap of variants with damaging/non-damaging missense variants separately, and calculated the OR of variants in each PPI bin to the lowest PPI bin. We tested the OR using a Fisher’s exact test.

### miRNA/RNA binding protein binding sites

We downloaded the miRNA binding sites from Target Scan Human and the RBP binding sites from RBP-Tar. All datasets were converted to genome coordinates GRCh38 (**Supplementary Methods 9**). For variants in non-coding regions, we counted the overlap of variants with miRNA/RBP binding site, and calculated the OR of variants in each PPI bin to the lowest PPI bin. We tested the OR using a Fisher’s exact test. The sequence of miRNA binding sites were obtained from miRNA SNP v4^30^.

### Masking strategy for rare variant testing

In RVAS, genetic variants are generally pooled to improve statistical power for rare variation. However, the optimal strategy for pooling of variants, or masking, is not known and likely not uniform across protein-coding genes^16,31^. From the various LoF, missense and frequency annotations, we therefore created many masking combinations, as shown in **Supplementary Fig. 12a**.

Based on LoFs only, we created up to two masks for each gene (LoF and LoFnoflag). For missense variants, for each prediction tool, we created a mask that included missense variants predicted as damaging by the given tool; we also created a mask that included missense variants predicted as damaging by any one of the four tools (miss1/4), and a strict masks including missense variants predicted as damaging by at least three of the four tools (miss3/4); as such, up to 6 missense masks were created per gene. We also created masks combining both LoF masks with each of the missense masks, yielding 2 × 6 = 12 additional LoF + missense masks. In total, therefore, 20 different masking schemes were created based on variant annotation.

In our masking, we also applied various filters determined by variant frequency. We essentially created masks for ultra-rare variants (MAFmax < 0.001%), masks for rare variants (MAFmax < 0.1%), and masks for rare and low-frequency variants (MAFmax < 1%). As such, for any given protein-coding gene, up to 20 × 3 = 60 different masks were created based on annotation and frequency.

### Burden association analyses

We performed RVAS with a dominant model using Regenie v4.1^23^. We used the same covariates as GWAS. After step 2, burden results were filtered based on the cMAC > 10. Of note, for all autosomal genes, the above approach was applied as described, combining both genetically-inferred males and genetically-inferred females in a single analysis. We did not analyze chromosome X.

### Cauchy combination

The RVAS would then produce up to 60 different mask-AF association results for each gene. To then produce a single test statistic for a gene, we used the Cauchy distribution test^13^ in a layered approach^32^. The Cauchy distribution allows for valid aggregation of many potentially correlated P-values, into a single omnibus P-value^13^. In our layered approach, for a given gene, we first combined P-values from all qualifying missense-only masks into a single ‘missense P-value’ using the Cauchy distribution; a similar approach was applied to combine LoF-only masks into a ‘LoF P-value’, and LoF+missense masks into a ‘LoF+missense’ P-value. Then, the missense Cauchy P-value, the LoF Cauchy P-value and the LoF+missense Cauchy P-value were combined in a second layer to produce the final Cauchy P-value for the given gene. All association and Cauchy P-values are two-sided unless otherwise specified. Significance threshold was set at FDR < 0.01 based on final Cauchy P-value. We defined novel gene-phenotype pairs when it has not been reported in the datasets described in **Supplementary Methods 11**.

### Protein structure stability

We used DynaMut2^33^ to assess the effects of variants on protein structures. We used PDB ID: 4CYF for *VNN1* and 6W25 for *MC4R*.

### Drug target and protein classification

We downloaded the Therapeutic Target Database^34^. We prioritized the development phases in the following order: Approved, Phase 4, NDA (New Drug Applications)/BLA (Biologics License Application), Phase 3, Phase 2, and Phase 1, and assigned each gene the highest phase available. We then grouped Approved and Phase 4 as “Approved”, NDA/BLA and Phase 3 as “Phase 3”, and Phase 2 and Phase 1 as “Phase 1–2”, with all remaining genes treated as the baseline. We downloaded the protein classification from the Human Protein Atlas^35^. We excluded those not related with protein function/localizations (“Cancer_related_genes”, “Candidate_cardiovascular_disease_genes”, “Disease_related_genes”, “FDA_approved_drug_targets”, “Human_disease_related_genes”, “Potential_drug_targets”, “Essential_proteins”). To assess the association of protein classes, GWAS genes, and RVAS genes with approved drug target genes, we performed a logistic regression model adjusted for inverse rank normalized LOEUF.

### Reporting summary

Further information on research design is available in the Nature Portfolio Reporting Summary linked to this article.

## Data availability

Summary results for the main analyses will be made available through the Cardiovascular Disease Knowledge Portal (https://cvd.hugeamp.org) upon publication. This study used data from the All of Us Research Program’s Registered/Controlled Tier

Dataset v8, available to authorized users on the Researcher Workbench. Access to individual level UK Biobank data, both phenotypic and genetic, is available to approved researchers through application on the UK Biobank website (https://www.ukbiobank.ac.uk). Additional information about registration for access to the data is available at http://www.ukbiobank.ac.uk/register-apply/. Use of UK Biobank data was performed under applications 17488 and 176602. The summary statistics of Pan-UKB can be downloaded using ukbb_pan_ancestry and ukb_common repositories. Results from Pan-UKB, the GIANT consortium, the Global Lipids Genetics Consortium, the blood pressure GWAS, and the whole-exome imputation study are shown in **Supplementary Data 46**.

## Code availability

The analysis was conducted using publicly available software. The URLs to obtain these software are described in **Supplementary Data 47**.

## Acknowledgements

We gratefully thank AoU participants, as this study would not have been possible without their contributions. This work was supported by grants from the National Institutes of Health to P.T.E. (1RO1HL092577, K24HL105780 and R01HL157635). This work was also supported by a grant from the American Heart Association to P.T.E. (18SFRN34110082 and 961045). N.E. was supported by the Uehara Memorial Foundation Fellowship, the Tsuchiya Memorial Foundation Fellowship, and the American Heart Association Postdoctoral Fellowship. S.J.J. was supported by the Junior Clinical Scientist Fellowship (03-007-2022-0035) from the Dutch Heart Foundation, and by an Amsterdam UMC Doctoral Fellowship. T.N. is supported by the National Heart, Lung, and Blood Institute (R00HL165024). S.K. is supported by the National Heart, Lung, and Blood Institute (R00HL169733). P.T.E. is supported by MAESTRIA (965286). S.H.C was supported by R01HL092577.

## Author Contributions

N.E., S.J.J, S.K. and P.T.E. conceived and designed the study. N.E. performed the main statistical analyses. N.E., S.J.J., S.H.C., and S.K. performed statistical data pre-processing within the AoU. S.J.J., and S.H.C. performed statistical data pre-processing within the UKB.T.N. contributed critically to the analysis plan. N.E. and P.T.E wrote the manuscript. All authors critically revised and approved the manuscript.

