## Supplementary Materials for "Integrating multi-ancestry common and rare variant mapping accelerates therapeutic target discovery"

References ………………...................................................................................................	48

**Supplementary Notes**

#### Supplementary Notes 1. Comparison of the pooled approach with the fixed effect meta-analysis in the simulation

The quality control and processing of the All of Us (AoU) are described in the **Methods**, **Supplementary Methods 1-3**, and **Supplementary Data 1**. To assess the genomic inflation factor (λ_GC_) and statistical power in GWAS of samples including mixed individuals, we simulated two groups: 100% EUR (pure EUR) and mixed individuals (33%:33%:33% AFR/AMR/EUR), each with 30,000 (44 phenotypes) and 150,000 subjects (10 phenotypes) (**Supplementary Fig.1a**, **Supplementary Data 3**, **Supplementary Methods 4-5**). We did not observe inflation in λ_GC_ in mixed individuals compared to pure EUR (**Supplementary Fig.1b** and **Supplementary Data 3**). The Linkage disequilibrium (LD) score regression (LDSC)[^1^](https://web.endnote.com/citations/eyJkaXNwbGF5VGV4dCI6IjEiLCJjaXRhdGlvbnMiOlt7ImJpYmxpb0NvbnRlbnQiOlt7ImF1dGhvcnMiOlsiQnVsaWstU3VsbGl2YW4sIEJyZW5kYW4gSyIsIkxvaCwgUG8tUnUiLCJGaW51Y2FuZSwgSGlsYXJ5IEsiLCJSaXBrZSwgU3RlcGhhbiIsIllhbmcsIEppYW4iLCJQYXR0ZXJzb24sIE5pY2siLCJEYWx5LCBNYXJrIEoiLCJQcmljZSwgQWxrZXMgTCIsIk5lYWxlLCBCZW5qYW1pbiBNIl0sImlzYm4iOiIxMDYxLTQwMzYiLCJ2b2x1bWUiOiI0NyIsInJzeG1sIjoiPHJlY29yZD48cmVmLXR5cGU%2BMTc8L3JlZi10eXBlPjxjb250cmlidXRvcnM%2BPGF1dGhvcnM%2BPGF1dGhvcj5CdWxpay1TdWxsaXZhbiwgQnJlbmRhbiBLPC9hdXRob3I%2BPGF1dGhvcj5Mb2gsIFBvLVJ1PC9hdXRob3I%2BPGF1dGhvcj5GaW51Y2FuZSwgSGlsYXJ5IEs8L2F1dGhvcj48YXV0aG9yPlJpcGtlLCBTdGVwaGFuPC9hdXRob3I%2BPGF1dGhvcj5ZYW5nLCBKaWFuPC9hdXRob3I%2BPGF1dGhvcj5QYXR0ZXJzb24sIE5pY2s8L2F1dGhvcj48YXV0aG9yPkRhbHksIE1hcmsgSjwvYXV0aG9yPjxhdXRob3I%2BUHJpY2UsIEFsa2VzIEw8L2F1dGhvcj48YXV0aG9yPk5lYWxlLCBCZW5qYW1pbiBNPC9hdXRob3I%2BPC9hdXRob3JzPjwvY29udHJpYnV0b3JzPjx0aXRsZXM%2BPHRpdGxlPkxEIFNjb3JlIHJlZ3Jlc3Npb24gZGlzdGluZ3Vpc2hlcyBjb25mb3VuZGluZyBmcm9tIHBvbHlnZW5pY2l0eSBpbiBnZW5vbWUtd2lkZSBhc3NvY2lhdGlvbiBzdHVkaWVzPC90aXRsZT48c2Vjb25kYXJ5LXRpdGxlPk5hdHVyZSBHZW5ldGljczwvc2Vjb25kYXJ5LXRpdGxlPjwvdGl0bGVzPjxkYXRlcz48eWVhcj4yMDE1PC95ZWFyPjwvZGF0ZXM%2BPHBhZ2VzPjI5MS0yOTU8L3BhZ2VzPjx2b2x1bWU%2BNDc8L3ZvbHVtZT48aXNibj4xMDYxLTQwMzY8L2lzYm4%2BPHVybHM%2BPHJlbGF0ZWQtdXJscz48dXJsPmh0dHBzOi8vZXNjaG9sYXJzaGlwLm9yZy9jb250ZW50L3F0NjdiNzEwc2ovcXQ2N2I3MTBzai5wZGY%2FdD1wdG52dmk8L3VybD48L3JlbGF0ZWQtdXJscz48L3VybHM%2BPGVsZWN0cm9uaWMtcmVzb3VyY2UtbnVtPjxzdHlsZSBmYWNlPVwidW5kZXJsaW5lXCI%2BMTAuMTAzOC9uZy4zMjExPC9zdHlsZT48L2VsZWN0cm9uaWMtcmVzb3VyY2UtbnVtPjxudW1iZXI%2BMzwvbnVtYmVyPjxyZWMtZ3VpZD42OGFhNTEzZi0xOTAzLTQzZTItYmExYi1hYmI0ZTQxYTQyNGE8L3JlYy1ndWlkPjxyZWMtdXNuPjI1PC9yZWMtdXNuPjwvcmVjb3JkPiIsInBhZ2VzIjoiMjkxLTI5NSIsInJlZmVyZW5jZVR5cGUiOiIxNyIsIm51bWJlciI6IjMiLCJ5ZWFyIjoiMjAxNSIsInRpdGxlIjoiTEQgU2NvcmUgcmVncmVzc2lvbiBkaXN0aW5ndWlzaGVzIGNvbmZvdW5kaW5nIGZyb20gcG9seWdlbmljaXR5IGluIGdlbm9tZS13aWRlIGFzc29jaWF0aW9uIHN0dWRpZXMiLCJndWlkIjoiNjhhYTUxM2YtMTkwMy00M2UyLWJhMWItYWJiNGU0MWE0MjRhIiwiZWxlY3Ryb25pY1Jlc291cmNlTnVtYmVyIjoiMTAuMTAzOC9uZy4zMjExIiwidXJsIjpbIm) intercept for all 44 phenotypes from 30,000 individuals was smaller than 1.0, which means there was no population stratification (**Supplementary Fig. 1b** and **Supplementary Data 3**). The LDSC intercept for all 10 phenotypes from 150,000 individuals exceeded 1.0, however, the all attenuation ratios were less than 0.2, suggesting no evidence of significant confounding (**Supplementary Fig. 1b** and **Supplementary Data 3**). The λ_GC_ derived from the pooled approach were nearly identical or slightly smaller than that from pure EUR. The λ_GC_ derived from the pooled approach and the meta-analysis were mostly concordant (**Supplementary Fig. 1c** and **Supplementary Data 3**). Out of 635 loci found in either the pooled analysis or meta-analysis, 485 were common, 87 reached significance were found only in the pooled analysis, and 63 only in the meta-analysis (**Supplementary Fig 1d**). P values from pooled analysis and meta-analysis were consistent (**Supplementary Fig 1e**). Similarly, we ran Rare Variant Association Studies (RVAS), and compared λ_GC_ derived from pooled analysis and pure EUR analysis. Again, the λ_GC_ derived from the pooled approach was identical or slightly smaller than that from pure EUR (**Supplementary Fig 1f**). Taken together, we concluded that pooled analysis is a controlled and powerful strategy for multi-ancestry GWASs and RVASs, and we employed this approach hereafter.

#### Supplementary Notes 2. Comparison of the pooled approach with the fixed effect meta-analysis for 624 quantitative traits

Our inclusive strategy (pooled analysis) allowed us to incorporate 24,290 PCA outliers (**Supplementary Fig.2b-c**), who had been conventionally excluded from genetic analysis. There are 415 phenotypes with sample size 1,000 - 10,000, 152 with 10,000 - 50,000, 21 with 50,000 - 100,000, and 36 with 100,000 or more (**Supplementary Fig.2d** and **Supplementary Data 4**). We assessed the genetic correlation for all phenotype combinations (194,376 pairs), and 481 phenotype pairs showed genetic correlation > 0.5 with P < 0.05 (Supplementary Data 4). For example, the coefficient between “Diastolic blood pressure” and “Computed diastolic blood pressure, mean of 2nd and 3rd measures” is 0.99.

Similar to what we did in the simulation study, we compared pooled analysis with meta-analysis for 624 phenotypes with the same sample size and variant sets. Out of 5,193 loci found in these analyses, 3,578 were common, 1,242 were unique to pooled analysis, and 375 were unique to the meta-analysis (**Supplementary Fig 3a**). P-values and beta were highly correlated between the two approaches (**Supplementary Fig 3b-c**), indicating that pooled analysis is a valid strategy in the real research settings. Next we compared pooled approaches for individuals before and after PCA outliers since we wanted to maximize the sample size (**Supplementary Fig 2b**). Including PCA outliers increased 566 loci compared to results from no PCA outliers (**Supplementary Fig 3d**) while beta and P-values were reasonably comparable (**Supplementary Fig 3e-f**).

#### Supplementary Notes 3. The quality check of GWAS results

Out of 1,982 ancestry-phenotype pairs, 1,932 ancestry-phenotype pairs showed λ_GC_ < 1.05, indicating that there was no inflation in most of the ancestry-phenotype pairs (**Supplementary Data 5**). Among the 50 remaining ancestry-phenotype pairs, leukocytes (measurement concept ID: 3000905) in AFR showed high attenuation ratio (0.5) while the λ_GC_ was 1.07 (**Supplementary Fig 4a** and **Supplementary Data 5**). Since the previous report showed blood cell traits in AFR showed quite high attenuation ratio[^2^](https://web.endnote.com/citations/eyJkaXNwbGF5VGV4dCI6IjIiLCJjaXRhdGlvbnMiOlt7Imd1aWQiOiJmM2IxMjA0ZS1kYTA2LTQ1YzMtYWE1OS05M2U2ZDEzMGE0MjEiLCJyZWNvcmQiOnsibnVtYmVyIjoiNjcwNiIsInJlYy1ndWlkIjoiZjNiMTIwNGUtZGEwNi00NWMzLWFhNTktOTNlNmQxMzBhNDIxIiwicmVjLXVzbiI6IjEyIiwiZGF0ZXMiOnsieWVhciI6IjIwMjQifSwicmVmLXR5cGUiOiIxNyIsInRpdGxlcyI6eyJzZWNvbmRhcnktdGl0bGUiOiJTY2llbmNlIiwidGl0bGUiOiJEaXZlcnNpdHkgYW5kIHNjYWxlOiBHZW5ldGljIGFyY2hpdGVjdHVyZSBvZiAyMDY4IHRyYWl0cyBpbiB0aGUgVkEgTWlsbGlvbiBWZXRlcmFuIFByb2dyYW0ifSwiZWxlY3Ryb25pYy1yZXNvdXJjZS1udW0iOnsic3R5bGUiOnsiIyI6IjEwLjExMjYvc2NpZW5jZS5hZGoxMTgyIiwiQGZhY2UiOiJ1bmRlcmxpbmUifX0sImlzYm4iOiIwMDM2LTgwNzUiLCJ2b2x1bWUiOiIzODUiLCJjb250cmlidXRvcnMiOnsiYXV0aG9ycyI6eyJhdXRob3IiOlsiVmVybWEsIEFudXJhZyIsIkh1ZmZtYW4sIEplbm5pZmVyIEUuIiwiUm9kcmlndWV6LCBBbGV4IiwiQ29uZXJ5LCBNaXRjaGVsbCIsIkxpdSwgTW9sZWkiLCJIbywgWXVrLUxhbSIsIktpbSwgWW91bmdkYWUiLCJIZWlzZSwgRGF2aWQgQS4iLCJHdWFyZSwgTGluZHNheSIsIlBhbmlja2FuLCBWaWR1bCBBeWFrdWxhbmdhcmEiLCJHYXJjb24sIEhlbGVuZSIsIkxpbmFyZXMsIEZyYW5jaWVsIiwiQ29zdGEsIExhdXJlbiIsIkdvZXRoZXJ0LCBJYW4iLCJUaXB0b24sIFJ5YW4iLCJIb25lcmxhdywgSmFjcXVlbGluZSIsIkRhdmllcywgTGF1cmEiLCJXaGl0Ym91cm5lLCBTdGFjZXkiLCJDb2hlbiwgSmVyZW15IiwiUG9zbmVyLCBEYW5pZWwgQy4iLCJTYW5nYXIsIFJhaHVsIiwiTXVycmF5LCBNaWNoYWVsIiwiV2FuZywgWHVhbiIsIkRvY2h0ZXJtYW5uLCBEYW5pZWwgUi4iLCJEZXZpbmVuaSwgUG9vcm5pbWEiLCJTaGksIFl1bmxpbmciLCJOYW5kaSwgVGFyYWsgTmF0aCIsIkFzc2ltZXMsIFRoZW1pc3RvY2xlcyBMLiIsIkJydW5ldHRlLCBDaGFybGVzIEEuIiwiQ2Fycm9sbCwgUm9iZXJ0IEouIiwiQ2xpZmZvcmQsIFJveWNlIiwiRHV2YWxsLCBTY290dCIsIkdlbGVybnRlciwgSm9lbCIsIkh1bmcsIEFkcmlhbmEiLCJJeWVuZ2FyLCBTdWRoYSBLLiIsIkpvc2VwaCwgSmFjb2IiLCJLZW1iZXIsIFJhY2hlbCIsIktyYW56bGVyLCBIZW5yeSIsIktyaXBrZSwgQ29sbGVlbiBNLiIsIkxldmV5LCBEYW5pZWwiLCJMdW9oLCBTaGl1aC1XZW4iLCJNZXJyaXR0LCBWaWN0b3JpYSBDLiIsIk92ZXJzdHJlZXQsIENhc3NpZSIsIkRlYWssIEpvc2VwaCBELiIsIkdyYW50LCBTdHJ1YW4gRi4gQS4iLCJQb2xpbWFudGksIFJlbmF0byIsIlJvdXNzb3MsIFBhbm9zIiwiU2hha3QsIEdhYnJpZWxsZSIsIlN1biwgWWFuIFYuIiwiVHNhbywgTm9haCIsIlZlbmthdGVzaCwgU2FuYW4iLCJWb2xvdWRha2lzLCBHZW9yZ2lvcyIsIkp1c3RpY2UsIEFteSIsIkJlZ29saSwgRWRtb24iLCJSYW1vbmksIFJhY2hlbCIsIlRvdXJhc3NpLCBHZW), and this ancestry-phenotype pair showed only a modest increase in λ_GC_, we kept this ancestry-phenotype pair. Then we ran GWAS for 70 matched phenotypes (**Supplementary Data 6**) in EUR and AFR in the UKB, and compared the effect estimates in EUR and AFR between AoU and UKB. Among 2,382 loci found in EUR, 2,240 reached genome-wide significance with concordant effect directions in UKB, 126 did not reach genome-wide significance threshold but was replicated with P < 0.05 and concordant effect directions, and 16 with P > 0.05 and discordant effect directions (**Supplementary Fig 4b** and **Supplementary Data 7**). Among 144 loci found in AFR, 103 reached genome-wide significance with concordant effect directions in UKB, 35 did not reach genome-wide significance threshold but was replicated with P < 0.05 and concordant effect directions, and 6 with P > 0.05 and discordant effect directions (**Supplementary Fig 4b** and **Supplementary Data 7**). Overall, most of the lead variants were replicated in UKB where phenotypes were common in both AoU and UKB. We noted one locus showed P < 1 × 10^-1000^. This was the known intron variant overlapping with *UGT1A4* associated with total bilirubin (**Supplementary Fig 4c**).

#### Supplementary Notes 4. Quality of the single- and multi-ancestry finemapping

We ran multi-ancestry finemapping on 6,181 phenotype-locus pairs, which reached genome-wide significance in at least one ancestry or the pooled analysis. Single-ancestry finemapping was conducted for loci that reached genome-wide significance within each ancestry: 2,899 loci in EUR, 883 loci in AFR, and 456 loci in AMR. After finemapping, we excluded credible sets with minimum purity of 0, resulting in 6,255 credible sets from 4,473 loci for multi-ancestry finemapping, 4,304 credible sets from 2,816 loci for EUR, 1,245 credible sets from 862 loci for AFR, and 582 credible sets from 445 for AMR. We compared phenotype-loci pairs with at least one credible sets with minimum purity > 0 in both single- and multi-ancestry finemapping. The credible set size decreased from a median of 5 (IQR 2–16) in EUR single-ancestry analyses to a median of 3 (IQR 1–9) in multi-ancestry analyses (**Supplementary Fig 6a**). Similar reductions were observed for AFR (from median 2, IQR 1–7 to median 2, IQR 1–5) and AMR (from median 4, IQR 1–10 to median 2, IQR 1–4). The maximum PIP within credible sets increased from a median of 0.437 (IQR 0.189–0.925) in EUR single-ancestry analyses to 0.60 (IQR 0.300–0.986) in multi-ancestry analyses(**Supplementary Fig 6b**). Similar trends were observed in AFR (from median 0.767, IQR 0.392–0.997 to median 0.874, IQR 0.468–1.000) and AMR (from median 0.584, IQR 0.305–0.977 to median 0.901, IQR 0.480–1.000). The number of credible sets with a best PIP > 0.95 was higher in the multi-ancestry analysis than in the EUR single-ancestry analysis (1,319 vs. 968) (**Supplementary Fig 6c**). Similar patterns were observed in AFR (752 vs. 449) and AMR (537 vs. 153).

#### Supplementary Notes 5. The AMR-specific missense variant associated with obesity

Rs79783591 is a AMR-specific, likely pathogenic missense variant of *MC4R*, supported by the Primate AI 3D score 0.805. Rs79783591 was not linkage disequilibrium with the known variant rs72982988, and their credible sets are also distinct (**Supplementary Fig.9a**). The MAF of rs79783591 was 0.098% for AMR and did not exist in EUR (**Supplementary Fig.9b**). In AoU AMR, rs79783591 was associated with increased body weight (beta 0.37, SE 0.047; P = 3.7 × 10^-15^) and obesity-related phenotypes (**Supplementary Fig.9c** and **Supplementary Data 20**). The replication analysis using MVP AMR showed that effect estimates were consistent between AoU and MVP (**Supplementary Fig.9c** and **Supplementary Data 20**). Rs79783591 was also associated with hypertension (OR 1.79; 95% CI 1.30 - 2.47), chronic venous insufficiency (OR 2.37; 95% CI 1.43 - 3.94), and other forms of chronic heart disease (OR 3.60; 95% CI 1.67 - 7.75) presumably caused by obesity (**Supplementary Fig.9d** and **Supplementary Data 21**). The ΔΔG^Stability^ was -0.82 kcal/mol (< -0.5 kcal/mol), indicating unstabilized structure, supposedly by lost hydrophobic bond with surrounding amino acids (**Supplementary Fig.9e**).

#### Supplementary Notes 6. The AMR-specific missense variant associated with height

Another example for the AMR-specific missense variant was rs367991535, a missense variant of *ADAMTS17* associated with height (beta -0.54, SE 0.08; P = 2.8 × 10-11) (**Supplementary Fig.10a** and **Supplementary Data 22**). Rs367991535 was independent from the lead variant rs4369638 (**Supplementary Fig.10a**).The MAF of rs367991535 was 0.04% for AMR, and did not exist for other ancestries (**Supplementary Fig.10b**). The effect direction of this variant on height in the T2D knowledge portal (T2DKP)[^3^](https://web.endnote.com/citations/eyJkaXNwbGF5VGV4dCI6IjMiLCJjaXRhdGlvbnMiOlt7InJlY29yZCI6eyJyZWYtdHlwZSI6IjE3IiwicmVjLXVzbiI6IjE1IiwibnVtYmVyIjoiNCIsInJlYy1ndWlkIjoiM2ZhMTA0ZGMtNWRmMi00OTlmLThlZDEtZTcwOWFhYjQzODBlIiwiZGF0ZXMiOnsieWVhciI6IjIwMjMifSwicGFnZXMiOiI2OTUtNzEwLmU2IiwiY29udHJpYnV0b3JzIjp7ImF1dGhvcnMiOnsiYXV0aG9yIjpbIkNvc3RhbnpvLCBNYXJpYSBDLiIsIkdyb3R0aHVzcywgTWFyY2luIFZvbiIsIk1hc3N1bmcsIEplZmZyZXkiLCJKYW5nLCBEb25na2V1biIsIkNhdWxraW5zLCBMaXp6IiwiS29lc3RlcmVyLCBSeWFuIiwiR2lsYmVydCwgQ2xpbnQiLCJXZWxjaCwgUnlhbiBQLiIsIkt1ZHRhcmthciwgUGFydWwiLCJIb2FuZywgUXV5IiwiQm91Z2h0b24sIEFuZHJldyBQLiIsIlNpbmdoLCBQcmVldGkiLCJTdW4sIFlpbmciLCJEdWJ5LCBNYXJjIiwiTW9yaW9uZG8sIEFubmllIiwiTmd1eWVuLCBUcmFuZyIsIlNtYWRiZWNrLCBQYXRyaWNrIiwiQWxleGFuZGVyLCBCZW5qYW1pbiBSLiIsIkJyYW5kZXMsIE1hY2tlbnppZSIsIkNhcm1pY2hhZWwsIE1hcnkiLCJEb3JuYm9zLCBQZXRlciIsIkdyZWVuLCBUb2RkIiwiSHVlbGxhcy1CcnVza2lld2ljeiwgS2VubmV0aCBDLiIsIkppLCBZdWUiLCJLbHVnZSwgQWxleGFuZHJpYSIsIk1jbWFob24sIEFvaWZlIEMuIiwiTWVyY2FkZXIsIEpvc2VwIE0uIiwiUnVlYmVuYWNrZXIsIE9saXZlciIsIlNlbmd1cHRhLCBTZWJhbnRpIiwiU3BhbGRpbmcsIER5bGFuIiwiVGFsaXVuLCBEYW5pZWwiLCJBYmVjYXNpcywgR29uw6dhbG8iLCJBa29sa2FyLCBCZWVuYSIsIkFsZXhhbmRlciwgQmVuamFtaW4gUi4iLCJBbGxyZWQsIE5pY2hvbGV0dGUgRC4iLCJBbHRzaHVsZXIsIERhdmlkIiwiQmVsb3csIEplbm5pZmVyIEUuIiwiQmVyZ21hbiwgUmljaGFyZCIsIkJldWxlbnMsIEpvbGluZSBXLkouIiwiQmxhbmdlcm8sIEpvaG4iLCJCb2VobmtlLCBNaWNoYWVsIiwiQm9rdmlzdCwgS3Jpc3RlciIsIkJvdHRpbmdlciwgRXJ3aW4iLCJCb3VnaHRvbiwgQW5kcmV3IFAuIiwiQm93ZGVuLCBEb25hbGQiLCJCcm9zbmFuLCBNLiBKdWxpYSIsIkJyb3duLCBDaHJpc3RvcGhlciIsIkJydXNraWV3aWN6LCBLZW5uZXRoIiwiQnVydHQsIE5vw6tsIFAuIiwiQ2FybWljaGFlbCwgTWFyeSIsIkNhdWxraW5zLCBMaXp6IiwiQ2Vib2xhLCBJbsOqcyIsIkNoYW1iZXJzLCBKb2huIiwiQ2hlbiwgWWlpLURlciBJZGEiLCJDaGVya2FzLCBBbmRyaXkiLCJDaHUsIEF1ZHJleSBZLiIsIkNsYXJrLCBDaHJpc3RvcGhlciIsIkNsYXVzc25pdHplciwgTWVsaW5hIiwiQ29zdGFuem8sIE1hcmlhIEMuIiwiQ294LCBOYW5jeSBKLiIsIkhvZWQsIE1hcmNlbCBEZW4iLCJEb25nLCBEdWMiLCJEdWJ5LCBNYXJjIiwiRHVnZ2lyYWxhLCBSYXZpbmRyYW5hdGgiLCJEdXB1aXMsIEpvc8OpZSIsIkVsZGVycywgUGV0cmEgSi5NLiIsIkVuZ3JlaXR6LCBKZXNzZSBNLiIsIkZhdW1hbiwgRXJpYyIsIkZlcnJlciwgSm9yZ2UiLCJGbGFubmljaywgSm) was consistent with that in AoU (**Supplementary Fig.10c-d** and **Supplementary Data 22**). These findings of AMR-specific rare variants highlighted the need for further WGS studies for AMR.

#### Supplementary Notes 7. The quality check of RVAS results

We found 8 phenotypes with inflation (n = 8 phenotypes with λ_95%,RVAS_ > 1.2, one phenotype with λ_95%_,_RVAS_ > 1.5; **Supplementary Fig 12b** and **Supplementary Data 26**). The same phenomenon was observed when we restricted it to variants with MAF_max_ < 0.1% (**Supplementary Fig 12b** and **Supplementary Data 26**). The λ_95%,RVAS_ were generally proportional to the λ_GC,GWAS_ (**Supplementary Fig 12c**), which was consistent with the previous report[^4^](https://web.endnote.com/citations/eyJkaXNwbGF5VGV4dCI6IjQiLCJjaXRhdGlvbnMiOlt7Imd1aWQiOiIwN2U1M2IwZC0xYzdmLTQwM2ItYjBlZS1mYmExNjE1NTI3MzAiLCJyZWNvcmQiOnsiaXNibiI6IjAwMjgtMDgzNiIsImVsZWN0cm9uaWMtcmVzb3VyY2UtbnVtIjp7InN0eWxlIjp7IiMiOiIxMC4xMDM4L3M0MTU4Ni0wMjItMDU2ODQteiIsIkBmYWNlIjoidW5kZXJsaW5lIn19LCJ0aXRsZXMiOnsidGl0bGUiOiJQb2x5Z2VuaWMgYXJjaGl0ZWN0dXJlIG9mIHJhcmUgY29kaW5nIHZhcmlhdGlvbiBhY3Jvc3MgMzk0LDc4MyBleG9tZXMiLCJzZWNvbmRhcnktdGl0bGUiOiJOYXR1cmUifSwidm9sdW1lIjoiNjE0IiwicGFnZXMiOiI0OTItNDk5IiwiY29udHJpYnV0b3JzIjp7ImF1dGhvcnMiOnsiYXV0aG9yIjpbIldlaW5lciwgRGFuaWVsIEouIiwiTmFkaWcsIEFqYXkiLCJKYWdhZGVlc2gsIEthcnRoaWsgQS4iLCJEZXksIEt1c2hhbCBLLiIsIk5lYWxlLCBCZW5qYW1pbiBNLiIsIlJvYmluc29uLCBFbGlzZSBCLiIsIkthcmN6ZXdza2ksIEtvbnJhZCBKLiIsIk%2FigJlDb25ub3IsIEx1a2UgSi4iXX19LCJyZWMtdXNuIjoiMzYiLCJyZWMtZ3VpZCI6IjA3ZTUzYjBkLTFjN2YtNDAzYi1iMGVlLWZiYTE2MTU1MjczMCIsIm51bWJlciI6Ijc5NDgiLCJkYXRlcyI6eyJ5ZWFyIjoiMjAyMyJ9LCJyZWYtdHlwZSI6IjE3In0sImJpYmxpb0NvbnRlbnQiOlt7InBhZ2VzIjoiNDkyLTQ5OSIsInJlZmVyZW5jZVR5cGUiOiIxNyIsImF1dGhvcnMiOlsiV2VpbmVyLCBEYW5pZWwgSi4iLCJOYWRpZywgQWpheSIsIkphZ2FkZWVzaCwgS2FydGhpayBBLiIsIkRleSwgS3VzaGFsIEsuIiwiTmVhbGUsIEJlbmphbWluIE0uIiwiUm9iaW5zb24sIEVsaXNlIEIuIiwiS2FyY3pld3NraSwgS29ucmFkIEouIiwiT%2BKAmUNvbm5vciwgTHVrZSBKLiJdLCJpc2JuIjoiMDAyOC0wODM2Iiwidm9sdW1lIjoiNjE0IiwicnN4bWwiOiI8cmVjb3JkPjxyZWYtdHlwZT4xNzwvcmVmLXR5cGU%2BPGNvbnRyaWJ1dG9ycz48YXV0aG9ycz48YXV0aG9yPldlaW5lciwgRGFuaWVsIEouPC9hdXRob3I%2BPGF1dGhvcj5OYWRpZywgQWpheTwvYXV0aG9yPjxhdXRob3I%2BSmFnYWRlZXNoLCBLYXJ0aGlrIEEuPC9hdXRob3I%2BPGF1dGhvcj5EZXksIEt1c2hhbCBLLjwvYXV0aG9yPjxhdXRob3I%2BTmVhbGUsIEJlbmphbWluIE0uPC9hdXRob3I%2BPGF1dGhvcj5Sb2JpbnNvbiwgRWxpc2UgQi48L2F1dGhvcj48YXV0aG9yPkthcmN6ZXdza2ksIEtvbnJhZCBKLjwvYXV0aG9yPjxhdXRob3I%2BT%2BKAmUNvbm5vciwgTHVrZSBKLjwvYXV0aG9yPjwvYXV0aG9ycz48L2NvbnRyaWJ1dG9ycz48dGl0bGVzPjx0aXRsZT5Qb2x5Z2VuaWMgYXJjaGl0ZWN0dXJlIG9mIHJhcmUgY29kaW5nIHZhcmlhdGlvbiBhY3Jvc3MgMzk0LDc4MyBleG9tZXM8L3RpdGxlPjxzZWNvbmRhcnktdGl0bGU%2BTmF0dXJlPC9zZWNvbmRhcnktdGl0bGU%2BPC90aXRsZXM%2BPGRhdGVzPjx5ZWFyPjIwMjM8L3llYXI%2BPC9kYXRlcz48cGFnZXM%2BNDkyLTQ5OTwvcGFnZXM%2BPHZvbHVtZT42MTQ8L3). Notable inflations were observed in body height (λ_95%,RVAS_ 1.678; λ_GC,GWAS_ 1.88) and body weight (λ_95%,RVAS_ 1.37; λ_GC,GWAS_ 1.70). Of note, all of the significant gene-phenotype associations in these eight phenotypes have been reported previously, indicating that these inflations were acceptable.

We compared these gene-phenotype pairs with previous reports. To this end, we checked if the gene-phenotype pairs were (1) genome-wide significant in the GeneBass (2) genome-wide significant in the Open Targets Platform (3) P < 0.05 in the GeneBass or (4) Not reported in the Open Targets Platform and P > 0.05 in the GeneBass. Out of 106 phenotypes which had at least one significant gene, 72 phenotypes were reported to have at least one significant gene in Open Targets Platform or GeneBass (363 gene-phenotype pairs). Out of 363 gene-phenotype pairs, 361 have been reported in Open Targets Platform or GeneBass (**Supplementary Fig 12d** and **Supplementary Data 27**). Two novel associations were *TNFRSF13B* - Albumin/Globulin ratio and *MRC1* - AST. Since we used different masks from the previous reports, we re-analyzed UKB using the same analysis pipeline. All the effect estimates from 363 gene-phenotype pairs in the same masks were concordant between AoU and UKB (**Supplementary Fig 12e** and **Supplementary Data 28**). The cumulative MAF (cMAF) were also correlated between AoU and UKB (**Supplementary Fig 12f** and **Supplementary Data 28**). As an example of novel associations, we showed that *MRC1* (the best mask in UKB was LoF + Primate AI 3D, MAF 1%) had strong association with AST (AoU: beta 1.49, SE 0.10, P = 4.7 × 10-51; UKB: beta 1.53, SE 0.06, P = 6.2 × 10-138) (**Supplementary Fig 13a** and **Supplementary Data 29**), and mild association with ALT. *MRC1* was also associated with alcoholic hepatitis, fatty liver disease and chronic liver disease in UKB (**Supplementary Fig 13b** and **Supplementary Data 30**). Taken together, although some phenotypes showed an inflated λ_95%,RVAS_, the significant gene-phenotype pairs, including novel associations, are reasonable results. In addition, we found two novel gene-phenotype pairs using our extensive analytical framework.

#### Supplementary Notes 8. Association between constraint and druggability

As we expected, genes supported by GWAS or RVAS were enriched in lower LOEUF (Loss-of-Function Observed/Expected Upper Bound Fraction), indicating these were enriched in constraint genes (**Supplementary Fig.15a** and **Supplementary Data 40**). The approved drug target genes were also enriched in the constraint genes (**Supplementary Fig.15a** and **Supplementary Data 40**).

#### Supplementary Notes 9. The association between protein classes and druggability

Certain classes of proteins are known to have higher druggability. For example, nuclear receptors (e.g. Peroxisome proliferator-activated receptor gamma [PPAR-γ inhibitors as hypoglycemic agents]) constitute targets for 15–20% of all pharmacologic drugs[^5^](https://web.endnote.com/citations/eyJkaXNwbGF5VGV4dCI6IjUiLCJjaXRhdGlvbnMiOlt7ImJpYmxpb0NvbnRlbnQiOlt7InJlY29yZFN0YXR1cyI6ImFjdGl2ZSIsInNlY29uZGFyeVRpdGxlIjoiU2lnbmFsIFRyYW5zZHVjdGlvbiBhbmQgVGFyZ2V0ZWQgVGhlcmFweSIsImVsZWN0cm9uaWNSZXNvdXJjZU51bWJlciI6IjEwLjEwMzgvczQxMzkyLTAyNS0wMjI3MC0zIiwiZ3JvdXBHdWlkcyI6W10sInllYXIiOiIyMDI1IiwibnVtYmVyIjoiMSIsInRpdGxlIjoiTnVjbGVhciByZWNlcHRvcnMgaW4gaGVhbHRoIGFuZCBkaXNlYXNlOiBzaWduYWxpbmcgcGF0aHdheXMsIGJpb2xvZ2ljYWwgZnVuY3Rpb25zIGFuZCBwaGFybWFjZXV0aWNhbCBpbnRlcnZlbnRpb25zIiwiZ3VpZCI6ImM0MjQwZTM3LTkxNTctNGM4ZS04OTY0LTlmYTZmZjhhYWE1NiIsInJlZmVyZW5jZVR5cGUiOiIxNyIsImF1dGhvcnMiOlsiSmluLCBQaW5nIiwiRHVhbiwgWGlydWkiLCJIdWFuZywgWmhhbyIsIkRvbmcsIFl1YW4iLCJaaHUsIEppYW5tZWkiLCJHdW8sIEh1aW1pbmciLCJUaWFuLCBIdWkiLCJab3UsIENoZW5nLUdhbmciLCJYaWUsIEtlIl0sImlzYm4iOiIyMDU5LTM2MzUiLCJ2b2x1bWUiOiIxMCIsInJzeG1sIjoiPHJlY29yZD48cmVmLXR5cGU%2BMTc8L3JlZi10eXBlPjxjb250cmlidXRvcnM%2BPGF1dGhvcnM%2BPGF1dGhvcj5KaW4sIFBpbmc8L2F1dGhvcj48YXV0aG9yPkR1YW4sIFhpcnVpPC9hdXRob3I%2BPGF1dGhvcj5IdWFuZywgWmhhbzwvYXV0aG9yPjxhdXRob3I%2BRG9uZywgWXVhbjwvYXV0aG9yPjxhdXRob3I%2BWmh1LCBKaWFubWVpPC9hdXRob3I%2BPGF1dGhvcj5HdW8sIEh1aW1pbmc8L2F1dGhvcj48YXV0aG9yPlRpYW4sIEh1aTwvYXV0aG9yPjxhdXRob3I%2BWm91LCBDaGVuZy1HYW5nPC9hdXRob3I%2BPGF1dGhvcj5YaWUsIEtlPC9hdXRob3I%2BPC9hdXRob3JzPjwvY29udHJpYnV0b3JzPjx0aXRsZXM%2BPHRpdGxlPk51Y2xlYXIgcmVjZXB0b3JzIGluIGhlYWx0aCBhbmQgZGlzZWFzZTogc2lnbmFsaW5nIHBhdGh3YXlzLCBiaW9sb2dpY2FsIGZ1bmN0aW9ucyBhbmQgcGhhcm1hY2V1dGljYWwgaW50ZXJ2ZW50aW9uczwvdGl0bGU%2BPHNlY29uZGFyeS10aXRsZT5TaWduYWwgVHJhbnNkdWN0aW9uIGFuZCBUYXJnZXRlZCBUaGVyYXB5PC9zZWNvbmRhcnktdGl0bGU%2BPC90aXRsZXM%2BPGRhdGVzPjx5ZWFyPjIwMjU8L3llYXI%2BPC9kYXRlcz48dm9sdW1lPjEwPC92b2x1bWU%2BPGlzYm4%2BMjA1OS0zNjM1PC9pc2JuPjxlbGVjdHJvbmljLXJlc291cmNlLW51bT48c3R5bGUgZmFjZT1cInVuZGVybGluZVwiPjEwLjEwMzgvczQxMzkyLTAyNS0wMjI3MC0zPC9zdHlsZT48L2VsZWN0cm9uaWMtcmVzb3VyY2UtbnVtPjxudW1iZXI%2BMTwvbnVtYmVyPjxyZWMtZ3VpZD5jNDI0MGUzNy05MTU3LTRjOGUtODk2NC05ZmE2ZmY4YWFhNTY8L3JlYy1ndWlkPjxyZWMtdXNuPjE5PC9yZWMtdXNuPjwvcmVjb3JkPiJ9XSwiZ3VpZCI6ImM0MjQwZTM3LTkxNTctNGM4ZS04OTY0LTlmYTZmZjhhYWE1NiIsInJlY29yZCI6eyJ2b2x1bWUiOi). Indeed, nuclear receptors and other known classes of druggable proteins, such as G protein-coupled receptors (GPCRs)[^6^](https://web.endnote.com/citations/eyJkaXNwbGF5VGV4dCI6IjYiLCJjaXRhdGlvbnMiOlt7ImJpYmxpb0NvbnRlbnQiOlt7ImVsZWN0cm9uaWNSZXNvdXJjZU51bWJlciI6IjEwLjEwMzgvczQxNTczLTAyNS0wMTEzOS15IiwiZ3JvdXBHdWlkcyI6W10sInJlY29yZFN0YXR1cyI6ImFjdGl2ZSIsInNlY29uZGFyeVRpdGxlIjoiTmF0dXJlIFJldmlld3MgRHJ1ZyBEaXNjb3ZlcnkiLCJhdXRob3JzIjpbIkxvcmVudGUsIEphdmllciBTw6FuY2hleiIsIlNva29sb3YsIEFsZWtzYW5kciBWLiIsIkZlcmd1c29uLCBHYXZpbiIsIlNjaGnDtnRoLCBIZWxnaSBCLiIsIkhhdXNlciwgQWxleGFuZGVyIFMuIiwiR2xvcmlhbSwgRGF2aWQgRS4iXSwiaXNibiI6IjE0NzQtMTc3NiIsInZvbHVtZSI6IjI0IiwicnN4bWwiOiI8cmVjb3JkPjxyZWYtdHlwZT4xNzwvcmVmLXR5cGU%2BPGNvbnRyaWJ1dG9ycz48YXV0aG9ycz48YXV0aG9yPkxvcmVudGUsIEphdmllciBTw6FuY2hlejwvYXV0aG9yPjxhdXRob3I%2BU29rb2xvdiwgQWxla3NhbmRyIFYuPC9hdXRob3I%2BPGF1dGhvcj5GZXJndXNvbiwgR2F2aW48L2F1dGhvcj48YXV0aG9yPlNjaGnDtnRoLCBIZWxnaSBCLjwvYXV0aG9yPjxhdXRob3I%2BSGF1c2VyLCBBbGV4YW5kZXIgUy48L2F1dGhvcj48YXV0aG9yPkdsb3JpYW0sIERhdmlkIEUuPC9hdXRob3I%2BPC9hdXRob3JzPjwvY29udHJpYnV0b3JzPjx0aXRsZXM%2BPHRpdGxlPkdQQ1IgZHJ1ZyBkaXNjb3Zlcnk6IG5ldyBhZ2VudHMsIHRhcmdldHMgYW5kIGluZGljYXRpb25zPC90aXRsZT48c2Vjb25kYXJ5LXRpdGxlPk5hdHVyZSBSZXZpZXdzIERydWcgRGlzY292ZXJ5PC9zZWNvbmRhcnktdGl0bGU%2BPC90aXRsZXM%2BPGRhdGVzPjx5ZWFyPjIwMjU8L3llYXI%2BPC9kYXRlcz48cGFnZXM%2BNDU4LTQ3OTwvcGFnZXM%2BPHZvbHVtZT4yNDwvdm9sdW1lPjxpc2JuPjE0NzQtMTc3NjwvaXNibj48ZWxlY3Ryb25pYy1yZXNvdXJjZS1udW0%2BPHN0eWxlIGZhY2U9XCJ1bmRlcmxpbmVcIj4xMC4xMDM4L3M0MTU3My0wMjUtMDExMzkteTwvc3R5bGU%2BPC9lbGVjdHJvbmljLXJlc291cmNlLW51bT48bnVtYmVyPjY8L251bWJlcj48cmVjLWd1aWQ%2BY2I3OTY5NWQtM2I3My00ZDlmLWIxMWYtNTM5ZTllOGJhZTQyPC9yZWMtZ3VpZD48cmVjLXVzbj4yMDwvcmVjLXVzbj48L3JlY29yZD4iLCJwYWdlcyI6IjQ1OC00NzkiLCJyZWZlcmVuY2VUeXBlIjoiMTciLCJudW1iZXIiOiI2IiwieWVhciI6IjIwMjUiLCJ0aXRsZSI6IkdQQ1IgZHJ1ZyBkaXNjb3Zlcnk6IG5ldyBhZ2VudHMsIHRhcmdldHMgYW5kIGluZGljYXRpb25zIiwiZ3VpZCI6ImNiNzk2OTVkLTNiNzMtNGQ5Zi1iMTFmLTUzOWU5ZThiYWU0MiJ9XSwicmVjb3JkIjp7ImNvbnRyaWJ1dG9ycyI6eyJhdXRob3JzIjp7ImF1dGhvciI6WyJMb3JlbnRlLCBKYXZpZXIgU8OhbmNoZXoiLCJTb2tvbG92LCBBbGVrc2FuZHIgVi4iLCJGZXJndXNvbiwgR2F2aW4iLCJTY2hpw7Z0aCwgSGVsZ2kgQi4iLCJIYXVzZXIsIEFsZXhhbmRlciBTLiIsIkdsb3JpYW0sIERhdmlkIEUuIl19fSwicGFnZXMiOi) and the voltage gated ion channels[^7^](https://web.endnote.com/citations/eyJkaXNwbGF5VGV4dCI6IjciLCJjaXRhdGlvbnMiOlt7ImJpYmxpb0NvbnRlbnQiOlt7InNlY29uZGFyeVRpdGxlIjoiRnJvbnRpZXJzIGluIFBoYXJtYWNvbG9neSIsInJlY29yZFN0YXR1cyI6ImFjdGl2ZSIsImd1aWQiOiI2NGIyYzc3Ny1iN2I0LTQ1ZjktYTQxNy0zYTA3NzFjYWJhNjEiLCJ0aXRsZSI6IkRydWdnYWJpbGl0eSBvZiBWb2x0YWdlLUdhdGVkIFNvZGl1bSBDaGFubmVsc%2BKAlEV4cGxvcmluZyBPbGQgYW5kIE5ldyBEcnVnIFJlY2VwdG9yIFNpdGVzIiwieWVhciI6IjIwMjIiLCJncm91cEd1aWRzIjpbXSwicmVmZXJlbmNlVHlwZSI6IjE3IiwiZWxlY3Ryb25pY1Jlc291cmNlTnVtYmVyIjoiMTAuMzM4OS9mcGhhci4yMDIyLjg1ODM0OCIsInZvbHVtZSI6IjEzIiwicnN4bWwiOiI8cmVjb3JkPjxyZWYtdHlwZT4xNzwvcmVmLXR5cGU%2BPGNvbnRyaWJ1dG9ycz48YXV0aG9ycz48YXV0aG9yPldpc2VkY2hhaXNyaSwgR29yYWdvdDwvYXV0aG9yPjxhdXRob3I%2BRWwtRGluLCBUYW1lciBNLiBHYW1hbDwvYXV0aG9yPjwvYXV0aG9ycz48L2NvbnRyaWJ1dG9ycz48dGl0bGVzPjx0aXRsZT5EcnVnZ2FiaWxpdHkgb2YgVm9sdGFnZS1HYXRlZCBTb2RpdW0gQ2hhbm5lbHPigJRFeHBsb3JpbmcgT2xkIGFuZCBOZXcgRHJ1ZyBSZWNlcHRvciBTaXRlczwvdGl0bGU%2BPHNlY29uZGFyeS10aXRsZT5Gcm9udGllcnMgaW4gUGhhcm1hY29sb2d5PC9zZWNvbmRhcnktdGl0bGU%2BPC90aXRsZXM%2BPGRhdGVzPjx5ZWFyPjIwMjI8L3llYXI%2BPC9kYXRlcz48dm9sdW1lPjEzPC92b2x1bWU%2BPGlzYm4%2BMTY2My05ODEyPC9pc2JuPjxlbGVjdHJvbmljLXJlc291cmNlLW51bT4xMC4zMzg5L2ZwaGFyLjIwMjIuODU4MzQ4PC9lbGVjdHJvbmljLXJlc291cmNlLW51bT48cmVjLWd1aWQ%2BNjRiMmM3NzctYjdiNC00NWY5LWE0MTctM2EwNzcxY2FiYTYxPC9yZWMtZ3VpZD48cmVjLXVzbj4yMTwvcmVjLXVzbj48L3JlY29yZD4iLCJpc2JuIjoiMTY2My05ODEyIiwiYXV0aG9ycyI6WyJXaXNlZGNoYWlzcmksIEdvcmFnb3QiLCJFbC1EaW4sIFRhbWVyIE0uIEdhbWFsIl19XSwicmVjb3JkIjp7InZvbHVtZSI6IjEzIiwidGl0bGVzIjp7InRpdGxlIjoiRHJ1Z2dhYmlsaXR5IG9mIFZvbHRhZ2UtR2F0ZWQgU29kaXVtIENoYW5uZWxz4oCURXhwbG9yaW5nIE9sZCBhbmQgTmV3IERydWcgUmVjZXB0b3IgU2l0ZXMiLCJzZWNvbmRhcnktdGl0bGUiOiJGcm9udGllcnMgaW4gUGhhcm1hY29sb2d5In0sImVsZWN0cm9uaWMtcmVzb3VyY2UtbnVtIjoiMTAuMzM4OS9mcGhhci4yMDIyLjg1ODM0OCIsImlzYm4iOiIxNjYzLTk4MTIiLCJjb250cmlidXRvcnMiOnsiYXV0aG9ycyI6eyJhdXRob3IiOlsiV2lzZWRjaGFpc3JpLCBHb3JhZ290IiwiRWwtRGluLCBUYW1lciBNLiBHYW1hbCJdfX0sImRhdGVzIjp7InllYXIiOiIyMDIyIn0sInJlYy11c24iOiIyMSIsInJlYy1ndWlkIjoiNjRiMmM3NzctYjdiNC00NWY5LWE0MTctM2EwNzcxY2FiYTYxIiwicmVmLXR5cGUiOiIxNyJ9LCJndWlkIjoiNjRiMmM3NzctYjdiNC), were enriched in the target genes of the approved drugs (**Supplementary Fig.15b** and **Supplementary Data 42**). The GWAS/RVAS genes were enriched in some of these protein classes. For example, both GWAS genes and RVAS genes were enriched in CD markers, which showed strong enrichment in the drug target genes (**Supplementary Fig.15b** and **Supplementary Data 43**). This suggests that the ability of GWAS/RVAS genes to identify highly druggable targets is partly driven by their enrichment in druggable protein classes. Hence, we performed multivariable logistic regression to test whether GWAS genes and RVAS genes were enriched in the approved drug targets independently of protein classes as well as constraints.

### **Supplementary Methods**

#### Supplementary Methods 1. Quality control and processing of the All of Us program

*Central quality control of All of Us* *data*

One of the goals set by AoU was to recruit individuals that have been, and continue to be, underrepresented in biomedical research because of limited access to healthcare[^8,9^](https://web.endnote.com/citations/eyJkaXNwbGF5VGV4dCI6IjgsOSIsImNpdGF0aW9ucyI6W3siYmlibGlvQ29udGVudCI6W3siZ3JvdXBHdWlkcyI6W10sImVsZWN0cm9uaWNSZXNvdXJjZU51bWJlciI6IjEwLjEwMTYvai5wYXR0ZXIuMjAyMi4xMDA1NzAiLCJzZWNvbmRhcnlUaXRsZSI6IlBhdHRlcm5zIiwicmVjb3JkU3RhdHVzIjoiYWN0aXZlIiwidm9sdW1lIjoiMyIsInJzeG1sIjoiPHJlY29yZD48cmVmLXR5cGU%2BMTc8L3JlZi10eXBlPjxjb250cmlidXRvcnM%2BPGF1dGhvcnM%2BPGF1dGhvcj5SYW1pcmV6LCBBbmRyZWEgSC48L2F1dGhvcj48YXV0aG9yPlN1bGllbWFuLCBMaW5hPC9hdXRob3I%2BPGF1dGhvcj5TY2hsdWV0ZXIsIERhdmlkIEouPC9hdXRob3I%2BPGF1dGhvcj5IYWx2b3Jzb24sIEFsZXNlPC9hdXRob3I%2BPGF1dGhvcj5RaWFuLCBKdW48L2F1dGhvcj48YXV0aG9yPlJhdHNpbWJhemFmeSwgRnJhbmNpczwvYXV0aG9yPjxhdXRob3I%2BTG9wZXJlbmEsIFJveGFuYTwvYXV0aG9yPjxhdXRob3I%2BTWF5bywgS2Vsc2V5PC9hdXRob3I%2BPGF1dGhvcj5CYXNmb3JkLCBNZWxpc3NhPC9hdXRob3I%2BPGF1dGhvcj5EZWZsYXV4LCBOaWNvbGU8L2F1dGhvcj48YXV0aG9yPk11dGh1cmFtYW4sIEthcnRoaWsgTi48L2F1dGhvcj48YXV0aG9yPk5hdGFyYWphbiwgS2FydGhpazwvYXV0aG9yPjxhdXRob3I%2BS2hvLCBBYmVsPC9hdXRob3I%2BPGF1dGhvcj5YdSwgSHVhPC9hdXRob3I%2BPGF1dGhvcj5XaWxraW5zLCBDb25zdWVsbzwvYXV0aG9yPjxhdXRob3I%2BQW50b24tQ3VsdmVyLCBIb2RhPC9hdXRob3I%2BPGF1dGhvcj5Cb2Vyd2lua2xlLCBFcmljPC9hdXRob3I%2BPGF1dGhvcj5DaWNlaywgTWluZTwvYXV0aG9yPjxhdXRob3I%2BQ2xhcmssIENoZXJ5bCBSLjwvYXV0aG9yPjxhdXRob3I%2BQ29obiwgRWxpemFiZXRoPC9hdXRob3I%2BPGF1dGhvcj5PaG5vLU1hY2hhZG8sIEx1Y2lsYTwvYXV0aG9yPjxhdXRob3I%2BU2NodWxseSwgU2hlcmkgRC48L2F1dGhvcj48YXV0aG9yPkFobWVkYW5pLCBCcmlhbiBLLjwvYXV0aG9yPjxhdXRob3I%2BQXJnb3MsIE1hcmlhPC9hdXRob3I%2BPGF1dGhvcj5Dcm9uaW4sIFJvYmVydCBNLjwvYXV0aG9yPjxhdXRob3I%2BT%2BKAmURvbm5lbGwsIENocmlzdG9waGVyPC9hdXRob3I%2BPGF1dGhvcj5Gb3VhZCwgTW9uYTwvYXV0aG9yPjxhdXRob3I%2BR29sZHN0ZWluLCBEYXZpZCBCLjwvYXV0aG9yPjxhdXRob3I%2BR3JlZW5sYW5kLCBQaGlsaXA8L2F1dGhvcj48YXV0aG9yPkhlYmJyaW5nLCBTY290dCBKLjwvYXV0aG9yPjxhdXRob3I%2BS2FybHNvbiwgRWxpemFiZXRoIFcuPC9hdXRob3I%2BPGF1dGhvcj5LaGF0cmksIFBhcmluZGE8L2F1dGhvcj48YXV0aG9yPktvcmYsIEJydWNlPC9hdXRob3I%2BPGF1dGhvcj5TbW9sbGVyLCBKb3JkYW4gVy48L2F1dGhvcj48YXV0aG9yPlNvZGVrZSwgU3RlcGhlbjwvYXV0aG9yPjxhdXRob3I%2BV2lsYmFua3MsIEpvaG48L2F1dGhvcj48YXV0aG9yPkhlbnRnZXMsIEp1c3RpbjwvYXV0aG9yPjxhdXRob3I%2BTW9ja3JpbiwgU3). Consistently, AoU prioritized underrepresented participants for genome sequencing and data collection, resulting in a diverse research population with rich phenotypic data. A stringent central quality control (QC) procedure was applied, as described in the program’s genomic quality report (<https://support.researchallofus.org/hc/en-us/articles/29390274413716-All-of-Us-Genomic-Quality-Report>). Each AoU Genome Center performed QC of the specimens obtained from the AoU Biobank. Sample preparation, normalization and DNA library construction have been reported previously[^10^](https://web.endnote.com/citations/eyJkaXNwbGF5VGV4dCI6IjEwIiwiY2l0YXRpb25zIjpbeyJndWlkIjoiMjA4NjIyYjAtYmM1ZS00MmRkLWJjODktNWI5YTYwMjZhOGE2IiwicmVjb3JkIjp7ImNvbnRyaWJ1dG9ycyI6eyJhdXRob3JzIjp7ImF1dGhvciI6WyJWZW5uZXIsIEVyaWMiLCJNdXpueSwgRG9ubmEiLCJTbWl0aCwgSm9zaHVhIEQuIiwiV2Fsa2VyLCBLaW1iZXJseSIsIk5lYmVuLCBDeW50aGlhIEwuIiwiTG9ja3dvb2QsIENocmlzdGluYSBNLiIsIkVtcGV5LCBQaGlsbGlwIEUuIiwiTWV0Y2FsZiwgR2luZ2VyIEEuIiwiS2FjaHVsaXMsIENocmlzIiwiTWlhbiwgU2FuYSIsIk11c2ljaywgQW5qZW5lIiwiUmVobSwgSGVpZGkgTC4iLCJIYXJyaXNvbiwgU3RldmVuIiwiR2FicmllbCwgU3RhY2V5IiwiR2liYnMsIFJpY2hhcmQgQS4iLCJOaWNrZXJzb24sIERlYm9yYWgiLCJaaG91LCBBbGljaWEgWS4iLCJEb2hlbnksIEtpbWJlcmx5IiwiT3plbmJlcmdlciwgQnJhZGxleSIsIlRvcHBlciwgU2NvdHQgRS4iLCJMZW5ub24sIE5pYWxsIEouIl19fSwiZWxlY3Ryb25pYy1yZXNvdXJjZS1udW0iOnsic3R5bGUiOnsiIyI6IjEwLjExODYvczEzMDczLTAyMi0wMTAzMS16IiwiQGZhY2UiOiJ1bmRlcmxpbmUifX0sInRpdGxlcyI6eyJzZWNvbmRhcnktdGl0bGUiOiJHZW5vbWUgTWVkaWNpbmUiLCJ0aXRsZSI6Ildob2xlLWdlbm9tZSBzZXF1ZW5jaW5nIGFzIGFuIGludmVzdGlnYXRpb25hbCBkZXZpY2UgZm9yIHJldHVybiBvZiBoZXJlZGl0YXJ5IGRpc2Vhc2UgcmlzayBhbmQgcGhhcm1hY29nZW5vbWljIHJlc3VsdHMgYXMgcGFydCBvZiB0aGUgQWxsIG9mIFVzIFJlc2VhcmNoIFByb2dyYW0ifSwiaXNibiI6IjE3NTYtOTk0WCIsInZvbHVtZSI6IjE0IiwicmVmLXR5cGUiOiIxNyIsInJlYy11c24iOiI0MCIsIm51bWJlciI6IjEiLCJyZWMtZ3VpZCI6IjIwODYyMmIwLWJjNWUtNDJkZC1iYzg5LTViOWE2MDI2YThhNiIsImRhdGVzIjp7InllYXIiOiIyMDIyIn19LCJiaWJsaW9Db250ZW50IjpbeyJncm91cEd1aWRzIjpbXSwiZWxlY3Ryb25pY1Jlc291cmNlTnVtYmVyIjoiMTAuMTE4Ni9zMTMwNzMtMDIyLTAxMDMxLXoiLCJyZWNvcmRTdGF0dXMiOiJhY3RpdmUiLCJzZWNvbmRhcnlUaXRsZSI6Ikdlbm9tZSBNZWRpY2luZSIsInJzeG1sIjoiPHJlY29yZD48cmVmLXR5cGU%2BMTc8L3JlZi10eXBlPjxjb250cmlidXRvcnM%2BPGF1dGhvcnM%2BPGF1dGhvcj5WZW5uZXIsIEVyaWM8L2F1dGhvcj48YXV0aG9yPk11em55LCBEb25uYTwvYXV0aG9yPjxhdXRob3I%2BU21pdGgsIEpvc2h1YSBELjwvYXV0aG9yPjxhdXRob3I%2BV2Fsa2VyLCBLaW1iZXJseTwvYXV0aG9yPjxhdXRob3I%2BTmViZW4sIEN5bnRoaWEgTC48L2F1dGhvcj48YXV0aG9yPkxvY2t3b29kLCBDaHJpc3RpbmEgTS48L2F1dGhvcj48YXV0aG9yPkVtcGV5LCBQaGlsbGlwIEUuPC9hdXRob3I%2BPGF1dGhvcj5NZXRjYWxmLCBHaW5nZXIgQS48L2F1dGhvcj48YXV0aG9yPkthY2h1bGlzLCBDaHJpczwvYXV0aG9yPjxhdXRob3I%2BTWlhbiwgU2FuYTwvYXV0aG9yPjxhdX), after which samples underwent WGS. Details on sequencing, variant calling and quality control are described in the program’s genomic data quality report [https://support.researchallofus.org/hc/en-us/articles/29390274413716-All-of-Us-Genomic-Quality-Report]. In brief, processing consisted of an initial per-sample QC, including fingerprint concordance (array vs. WGS data), sex concordance (genetically determined vs. self-reported), cross-individual contamination rate and coverage to detect major errors, such as sample swaps or contamination. Participants who failed these tests were removed from the release. The WGS variants were then called jointly to reduce systematic biases. Additional sample QC procedures were then performed on the joint callsets, including hard threshold flagging (e.g., number of SNPs: < 2.4M and > 5.0M) and population outlier flagging. Variants QC was performed after sample QC, flagging specific variants in the callset. Processes included hard threshold filters (e.g., ExcessHet, QUAL score) and Allele-Specific Variant Quality Score Recalibration (AS-VQSR or VQSR).

*Additional quality control of All of Us data*

Using the centrally QCed genomic data, we performed additional QC steps at the genotype and variant level. Specifically, we obtained genotypes in PLINK binary format from the jointly called ACAF (an Allele Count/Allele Frequency) call set (v8) provided by AoU. This dataset includes only high-quality genotypes, with multi-allelic variants already split into bi-allelic variants and variants with allele frequency (AF) = 0 removed, as described. We further filtered variants based on the following criteria: (1) monomorphic variants, (2) call rate < 90%, (3) Hardy-Weinberg Equilibrium (HWE) P value < 1 × 10^-15^, (4) low complexity region, and (5) blacklist region (**Supplementary Data 46**). HWE was calculated within each ancestry for ancestry-specific analyses and across all ancestries for pooled analyses. For sample-level quality control, we excluded individuals flagged or identified as having known issues by AoU. We retained only samples with dragen_sex_ploidy equal to "XX" or "XY", excluded samples with genotype missingness > 5%, and removed potential duplicates. Duplicate resolution prioritized retaining individuals with linked electronic health record (EHR) data, followed by higher call rate (with greater weight given to EHR availability). Potential duplicates were identified using KING v2.3.2, with one sample from each pair flagged for removal if the heterozygous concordance exceeded 0.8. It resulted in 410,400 participants in the current analysis dataset. Out of these, 315,536 were linked with the electronic health record (EHR) data.

*Centrally provided principal component analysis*

AoU applied the analysis pipeline used in the gnomAD v3.1 release [https://gnomad.broadinstitute.org/news/2020-10-gnomad-v3-1-new-content-methods-annotations-and-data-availability/] to calculate principal components (PCs) of ancestry and assign ancestry labels. Specifically, AoU first identified 130,660 high-quality sites that can be called accurately in both the Human Genome Diversity Project (HGDP) and 1000 Genomes (1000G) dataset (training sample) and the AoU dataset. High-quality sites were defined as autosomal, bi-allelic single nucleotide variants (SNVs) with a MAF > 0.1% and a call rate > 99%. These sites were then LD-pruned with a cut-off of r^2^ = 0.1. AoU calculated the first 16 PCs in the training sample (HGDP and 1000G, using the hwe_normalized_pca() Hail function) with high-quality SNVs and projected the AoU sample into the PCA space to generate the first 16 PCs. The number of PCs was determined in the gnomAD resource that the first 16 PCs captured global ancestry variation well.

*Principal component analysis using internal datasets*

To infer population structure across all samples, we conducted principal component analysis (PCA), as the components provided by the AoU were based on external reference datasets (HGDP and 1000 Genomes Project) as described above. We first generated a filtered set of autosomal variants for both relatedness estimation and PCA. Variants were required to have a MAF > 1% and genotype missingness < 1%. LD pruning was performed using PLINK with the parameters --indep-pairwise 500 200 0.1 and --indep-pairwise 2000 400 0.1. Variants located in long-range LD regions were excluded, and 100,000 variants were randomly selected from the remaining set. Based on this variant set, we estimated pairwise relatedness with KING v2.3.2, and identified unrelated individuals as those with a kinship coefficient < 0.042. PCA was then performed using flashPCA v2.0. PCs were computed on the unrelated individuals, and the resulting components were projected onto the remaining samples. We calculated PCs using all individuals, including those in the “Other” category, as described in the “*Ancestry assignment*” section for the pooled analysis. For ancestry-specific analyses, PCs were recalculated within each ancestry after excluding individuals in the “Other” category.

#### Supplementary Methods 2. Whole-genome sequencing data from the UK Biobank

We used the WGS data released to the DNAnexus platform. To this end, we used the pVCF files released in 2024, processed through the DRAGEN pipeline (https://www.ukbiobank.ac.uk/media/c3zpw015/uk-biobank-final-whole-genome-sequencing-release-faqs_v3.pdf). We processed the >150,000 pVCF files through various steps, which included genotype-level, variant-level, and sample-level QC procedures. After central filtering, we identified WGS data for 490,542 samples, which entered the pipeline described below.

First, we processed the pVCF files by performing genotype-level refinement, and subsequent trimming of the VCF meta-data to reduce file size. To this end, bcftools (v1.16) with the setGT plugin (v1.20) was used to set low-quality genotypes to missing; genotypes flagged as ‘lowDepth’ or that failed the genotype-level DRAGEN machine-learning algorithm were set to missing. For details, see https://github.com/seanjosephjurgens/vcf_trimmer_genotypeqcer.

Next, we applied variant-level QC on the processed pVCF files. Using bcftools (v1.16), we removed variants that failed the variant-level DRAGEN machine-learning algorithm, or failed any other filter (ie INFO=/=PASS), after which multi-allelic variants were split to represent several biallelic variants. For details, see https://github.com/seanjosephjurgens/vcf_splitter_variantqcer. The pVCF files were then merged into larger pVCF files (50-300 per chromosome; see https://github.com/seanjosephjurgens/vcf_merger), after which we converted these pVCFs to PLINK2 format (see https://github.com/seanjosephjurgens/vcf2gds/tree/PLINK2), after which these chunks were merged into a single PLINK2 file per chromosome (see https://github.com/seanjosephjurgens/plink_merger). We then used the minrep() function from hail (v0.2.116) to normalize variant IDs and to convert the IDs to minimal representation. Finally, we used PLINK2 (vLinux_avx2_20240818) to remove variants that failed specific filters, including those with >0.1 missingness, those with minor allele count <1, and those with ExcessHet P < 1 × 10^-20^ (or HWE test P < 1 × 10^-20^ for X-chromosome variants). This process resulted in 1,195,517,695 autosomal, 47,666,140 non-autosomal X-chromosal, and 1,626,190 pseudo-autosomal X-chromosomal variants after variant-level QC.

We then defined a filtered genomic dataset, of pruned high-quality autosomal variants, for several downstream sample-QC procedures (using PLINK2 v-linux_x86_64_20240818). This filtered dataset was made by filtering to variants with MAF > 1%, missingness <1%, LD-pruned (--indep-pairwise 500 200 0.1 and --indep-pairwise 2000 400 0.1), excluding long-range LD regions, and finally taking 100k random markers.

Using this filtered dataset, we computed sample relatedness, ancestry assignments, and to perform PCA for the UKB samples: First, king2 (v2.3.2) was used to estimate heterozygote concordance rates and kinship coefficients for each sample-sample combination. Then, flashPCA (v2.0) was used to perform PCA on unrelated (kinship < 0.042) samples, with subsequent projection of PCs onto the remaining samples. Ancestry assignments were then performed using ADMIXTURE (v.1.3.0), by i) merging the data with the 1KG dataset, ii) sequently pruning variants in each continental superpopulation within the 1KG samples, iii) learning ancestry probabilities from the 1KG samples, and iv) projecting ancestry assignments onto the UKB samples; samples were assigned to a continental ancestry if the probability was >=0.8.

We then applied sample-level QC filtering. We first removed all samples with revoked consents (N = 245 removed). We then flagged individuals with ambiguous or mismatched genetically-inferred sex, as compared to their self-reported sex (N = 210). To this end, we used PLINK2 (v-linux_x86_64_20240818) to identify sex-mismatched samples, by computing X-chromosomal inbreeding coefficients (F) for each sample (using non-PAR X-chromosomal variants with MAF > 0.5%, missingness <1%, pruned using –indep-pairwise 500 200 0.1, and then pruned again using --indep-pairwise 2000 400 0.1); genetically-determined female sex was assigned at F < 0.5, while genetically-determined male sex was assigned at F > 0.8. We then removed one sample from duplicated pairs (using a heterozygote concordance rate > 0.8; N = 235 flagged). We then flagged samples that did not pass a centrally-computed metric for DNA quality (N = 239 flagged); samples that had a DNA contamination score of >= 2 (N = 67 flagged); samples that had high missingness (> 1% across all autosomal variants; N = 0 flagged); and samples that were outliers for various other metrics, including the PC-adjusted Ti/Tv ratio, PC-adjusted Het/Hom ratio, PC-adjusted SNV/indel ratio, and the PC-adjusted number of singletons (N = 1653 flagged). Overall, of the 490,542 initial samples, 490,296 had appropriate consents, of which 487,917 remained after removing all flagged samples.

#### Supplementary Methods 3. Ancestry assignment

To assign ancestry categories to the WGS participants, AoU trained a random forest classifier on the HGDP and the 1000G samples with known ancestry labels using the 16 PCs and applied the model to the AoU sample. Therefore, available ancestry categories were those used in the gnomAD, HGDP and 1000G resources, including AFR, AMR, EAS, MID, EUR, SAS, and Other. Ancestry categories were assigned to participants based on the probability generated by the random forest classifier. A cut-off of 75% was used, and all remaining samples were assigned to the "Other" group. For GWAS pooled analyses and RVAT, we included all populations, including the “Other” category, whereas for population-specific analyses, the “Other” category was excluded.

#### Supplementary Methods 4. Phenotype definition

*Fitbit data*

For number of steps from the Fitbit data, a valid day was defined as a participant wearing the Fitbit for at least 10 hours per day and reporting 100 - 45,000 steps per day[^11^](https://web.endnote.com/citations/eyJkaXNwbGF5VGV4dCI6IjExIiwiY2l0YXRpb25zIjpbeyJndWlkIjoiMmNiZDU0ZjItY2RkMS00NjhkLWE0ZWMtZjc2M2QwYmQwOGI4IiwicmVjb3JkIjp7InBhZ2VzIjoiMjMwMS0yMzA4IiwiY29udHJpYnV0b3JzIjp7ImF1dGhvcnMiOnsiYXV0aG9yIjpbIk1hc3RlciwgSGlyYWwiLCJBbm5pcywgSmVmZnJleSIsIkh1YW5nLCBTaGkiLCJCZWNrbWFuLCBKb3NodWEgQS4iLCJSYXRzaW1iYXphZnksIEZyYW5jaXMiLCJNYXJnaW5lYW4sIEtheWxhIiwiQ2Fycm9sbCwgUm9iZXJ0IiwiTmF0YXJhamFuLCBLYXJ0aGlrIiwiSGFycmVsbCwgRnJhbmsgRS4iLCJSb2RlbiwgRGFuIE0uIiwiSGFycmlzLCBQYXVsIiwiQnJpdHRhaW4sIEV2YW4gTC4iXX19LCJpc2JuIjoiMTA3OC04OTU2IiwidGl0bGVzIjp7InNlY29uZGFyeS10aXRsZSI6Ik5hdHVyZSBNZWRpY2luZSIsInRpdGxlIjoiQXNzb2NpYXRpb24gb2Ygc3RlcCBjb3VudHMgb3ZlciB0aW1lIHdpdGggdGhlIHJpc2sgb2YgY2hyb25pYyBkaXNlYXNlIGluIHRoZSBBbGwgb2YgVXMgUmVzZWFyY2ggUHJvZ3JhbSJ9LCJlbGVjdHJvbmljLXJlc291cmNlLW51bSI6eyJzdHlsZSI6eyIjIjoiMTAuMTAzOC9zNDE1OTEtMDIyLTAyMDEyLXciLCJAZmFjZSI6InVuZGVybGluZSJ9fSwidm9sdW1lIjoiMjgiLCJyZWYtdHlwZSI6IjE3IiwicmVjLWd1aWQiOiIyY2JkNTRmMi1jZGQxLTQ2OGQtYTRlYy1mNzYzZDBiZDA4YjgiLCJudW1iZXIiOiIxMSIsInJlYy11c24iOiI0MSIsImRhdGVzIjp7InllYXIiOiIyMDIyIn19LCJiaWJsaW9Db250ZW50IjpbeyJlbGVjdHJvbmljUmVzb3VyY2VOdW1iZXIiOiIxMC4xMDM4L3M0MTU5MS0wMjItMDIwMTItdyIsImdyb3VwR3VpZHMiOltdLCJzZWNvbmRhcnlUaXRsZSI6Ik5hdHVyZSBNZWRpY2luZSIsInJlY29yZFN0YXR1cyI6ImFjdGl2ZSIsImlzYm4iOiIxMDc4LTg5NTYiLCJhdXRob3JzIjpbIk1hc3RlciwgSGlyYWwiLCJBbm5pcywgSmVmZnJleSIsIkh1YW5nLCBTaGkiLCJCZWNrbWFuLCBKb3NodWEgQS4iLCJSYXRzaW1iYXphZnksIEZyYW5jaXMiLCJNYXJnaW5lYW4sIEtheWxhIiwiQ2Fycm9sbCwgUm9iZXJ0IiwiTmF0YXJhamFuLCBLYXJ0aGlrIiwiSGFycmVsbCwgRnJhbmsgRS4iLCJSb2RlbiwgRGFuIE0uIiwiSGFycmlzLCBQYXVsIiwiQnJpdHRhaW4sIEV2YW4gTC4iXSwicnN4bWwiOiI8cmVjb3JkPjxyZWYtdHlwZT4xNzwvcmVmLXR5cGU%2BPGNvbnRyaWJ1dG9ycz48YXV0aG9ycz48YXV0aG9yPk1hc3RlciwgSGlyYWw8L2F1dGhvcj48YXV0aG9yPkFubmlzLCBKZWZmcmV5PC9hdXRob3I%2BPGF1dGhvcj5IdWFuZywgU2hpPC9hdXRob3I%2BPGF1dGhvcj5CZWNrbWFuLCBKb3NodWEgQS48L2F1dGhvcj48YXV0aG9yPlJhdHNpbWJhemFmeSwgRnJhbmNpczwvYXV0aG9yPjxhdXRob3I%2BTWFyZ2luZWFuLCBLYXlsYTwvYXV0aG9yPjxhdXRob3I%2BQ2Fycm9sbCwgUm9iZXJ0PC9hdXRob3I%2BPGF1dGhvcj5OYXRhcmFqYW4sIEthcnRoaWs8L2F1dGhvcj48YXV0aG9yPkhhcnJlbGwsIEZyYW5rIEUuPC9hdX). A valid sleep was defined as the `is_main_sleep` being TRUE, sleep time > 180 minutes, and not missing details of sleep information (such as REM, non-REM, light sleep and deep sleep).

*The Exploring The Mind dataset*

AoU provided four “the Exploring The Mind (EtM) tasks”: City of Mountain, Now or Later, Left or Right, and Guess the Emotion. The City of Mountain is called a gradual onset continuous performance task (gradCPT). It tests how fast participants respond, or how well participants resist responding, to a changing scene. The Now or Later task, a delay discounting task, measures a participant’s individual level of temporal discounting or how long they are willing to wait for a certain reward. The Left or Right task, a Flanker test, measures how well participants can focus in distracting environments. The Guess the Emotion task, a version of a Facial Emotional Recognition task, asks participants to look at pictures of people and name the emotions based on their facial expressions.

#### Supplementary Methods 5. Generating simulation datasets

To prove the validity of the pooled approach, we simulated two groups: 100% European (pure EUR) and mixed individuals (33%:33%:33% AFR/AMR/EUR), with sample sizes of 30,000 and 150,000 for each group. We selected phenotypes for which both the pure EUR and mixed individuals datasets could be generated; that is, for the 30,000-sample dataset, the phenotype was measured in 30,000 Europeans and in 10,000 individuals from each of EUR, AFR, and AMR populations. The EUR individuals in the pure EUR group and in the mixed individuals group were the same. As a result, 44 phenotypes were selected for the 30,000-sample dataset, and 10 phenotypes for the 150,000-sample dataset.

#### Supplementary Methods 6. Annotation and rare variant burden testing

The Variant Effect Predictor (VEP, v105)[^12^](https://web.endnote.com/citations/eyJkaXNwbGF5VGV4dCI6IjEyIiwiY2l0YXRpb25zIjpbeyJyZWNvcmQiOnsiZGF0ZXMiOnsieWVhciI6IjIwMTYifSwidXJscyI6eyJyZWxhdGVkLXVybHMiOnsidXJsIjoiaHR0cDovL2V1cm9wZXBtYy5vcmcvYXJ0aWNsZXMvcG1jNDg5MzgyNT9wZGY9cmVuZGVyIn19LCJudW1iZXIiOiIxIiwicmVjLWd1aWQiOiJhNGY0M2EyMy03YTcxLTRiZWItYjA5ZS02MzYzNzA5NDhiNjYiLCJyZWMtdXNuIjoiMjgiLCJyZWYtdHlwZSI6IjE3Iiwidm9sdW1lIjoiMTciLCJlbGVjdHJvbmljLXJlc291cmNlLW51bSI6eyJzdHlsZSI6eyIjIjoiMTAuMTE4Ni9zMTMwNTktMDE2LTA5NzQtNCIsIkBmYWNlIjoidW5kZXJsaW5lIn19LCJ0aXRsZXMiOnsic2Vjb25kYXJ5LXRpdGxlIjoiR2Vub21lIEJpb2xvZ3kiLCJ0aXRsZSI6IlRoZSBFbnNlbWJsIFZhcmlhbnQgRWZmZWN0IFByZWRpY3RvciJ9LCJpc2JuIjoiMTQ3NC03NjBYIiwiY29udHJpYnV0b3JzIjp7ImF1dGhvcnMiOnsiYXV0aG9yIjpbIk1jbGFyZW4sIFdpbGxpYW0iLCJHaWwsIExhdXJlbnQiLCJIdW50LCBTYXJhaCBFLiIsIlJpYXQsIEhhcnByZWV0IFNpbmdoIiwiUml0Y2hpZSwgR3JhaGFtIFIuIFMuIiwiVGhvcm1hbm4sIEFuamEiLCJGbGljZWssIFBhdWwiLCJDdW5uaW5naGFtLCBGaW9uYSJdfX19LCJndWlkIjoiYTRmNDNhMjMtN2E3MS00YmViLWIwOWUtNjM2MzcwOTQ4YjY2IiwiYmlibGlvQ29udGVudCI6W3sic2Vjb25kYXJ5VGl0bGUiOiJHZW5vbWUgQmlvbG9neSIsInJlY29yZFN0YXR1cyI6ImFjdGl2ZSIsImVsZWN0cm9uaWNSZXNvdXJjZU51bWJlciI6IjEwLjExODYvczEzMDU5LTAxNi0wOTc0LTQiLCJncm91cEd1aWRzIjpbXSwidXJsIjpbImh0dHA6Ly9ldXJvcGVwbWMub3JnL2FydGljbGVzL3BtYzQ4OTM4MjU%2FcGRmPXJlbmRlciJdLCJ0aXRsZSI6IlRoZSBFbnNlbWJsIFZhcmlhbnQgRWZmZWN0IFByZWRpY3RvciIsInllYXIiOiIyMDE2IiwibnVtYmVyIjoiMSIsImd1aWQiOiJhNGY0M2EyMy03YTcxLTRiZWItYjA5ZS02MzYzNzA5NDhiNjYiLCJyZWZlcmVuY2VUeXBlIjoiMTciLCJhdXRob3JzIjpbIk1jbGFyZW4sIFdpbGxpYW0iLCJHaWwsIExhdXJlbnQiLCJIdW50LCBTYXJhaCBFLiIsIlJpYXQsIEhhcnByZWV0IFNpbmdoIiwiUml0Y2hpZSwgR3JhaGFtIFIuIFMuIiwiVGhvcm1hbm4sIEFuamEiLCJGbGljZWssIFBhdWwiLCJDdW5uaW5naGFtLCBGaW9uYSJdLCJpc2JuIjoiMTQ3NC03NjBYIiwidm9sdW1lIjoiMTciLCJyc3htbCI6IjxyZWNvcmQ%2BPHJlZi10eXBlPjE3PC9yZWYtdHlwZT48Y29udHJpYnV0b3JzPjxhdXRob3JzPjxhdXRob3I%2BTWNsYXJlbiwgV2lsbGlhbTwvYXV0aG9yPjxhdXRob3I%2BR2lsLCBMYXVyZW50PC9hdXRob3I%2BPGF1dGhvcj5IdW50LCBTYXJhaCBFLjwvYXV0aG9yPjxhdXRob3I%2BUmlhdCwgSGFycHJlZXQgU2luZ2g8L2F1dGhvcj48YXV0aG9yPlJpdGNoaWUsIEdyYWhhbSBSLiBTLjwvYXV0aG9yPjxhdXRob3I%2BVGhvcm1hbm4sIEFuamE8L2F1dGhvcj48YXV0aG9yPkZsaWNlaywgUGF1) with the LOFTEE plugin[^13^](https://web.endnote.com/citations/eyJkaXNwbGF5VGV4dCI6IjEzIiwiY2l0YXRpb25zIjpbeyJyZWNvcmQiOnsicmVmLXR5cGUiOiIxNyIsImRhdGVzIjp7InllYXIiOiIyMDIwIn0sInJlYy11c24iOiIyOSIsInJlYy1ndWlkIjoiNzcxNTU0OWYtNTcxOC00ZWFhLTk3NDUtNjI5YTI3OWFlMmI4IiwibnVtYmVyIjoiNzgwOSIsImNvbnRyaWJ1dG9ycyI6eyJhdXRob3JzIjp7ImF1dGhvciI6WyJLYXJjemV3c2tpLCBLb25yYWQgSi4iLCJGcmFuY2lvbGksIExhdXJlbnQgQy4iLCJUaWFvLCBHcmFjZSIsIkN1bW1pbmdzLCBCZXJ5bCBCLiIsIkFsZsO2bGRpLCBKZXNzaWNhIiwiV2FuZywgUWluZ2JvIiwiQ29sbGlucywgUnlhbiBMLiIsIkxhcmljY2hpYSwgS3Jpc3RlbiBNLiIsIkdhbm5hLCBBbmRyZWEiLCJCaXJuYmF1bSwgRGFuaWVsIFAuIiwiR2F1dGhpZXIsIExhdXJhIEQuIiwiQnJhbmQsIEhhcnJpc29uIiwiU29sb21vbnNvbiwgTWF0dGhldyIsIldhdHRzLCBOaWNob2xhcyBBLiIsIlJob2RlcywgRGFuaWVsIiwiU2luZ2VyLUJlcmssIE1vcmllbCIsIkVuZ2xhbmQsIEVsZWluYSBNLiIsIlNlYWJ5LCBFbGVhbm9yIEcuIiwiS29zbWlja2ksIEphY2sgQS4iLCJXYWx0ZXJzLCBSYXltb25kIEsuIiwiVGFzaG1hbiwgS2F0aGVyaW5lIiwiRmFyam91biwgWW9zc2kiLCJCYW5rcywgRXJpYyIsIlBvdGVyYmEsIFRpbW90aHkiLCJXYW5nLCBBcmN0dXJ1cyIsIlNlZWQsIENvdHRvbiIsIldoaWZmaW4sIE5pY29sYSIsIkNob25nLCBKZXNzaWNhIFguIiwiU2Ftb2NoYSwgS2FpdGxpbiBFLiIsIlBpZXJjZS1Ib2ZmbWFuLCBFbW1hIiwiWmFwcGFsYSwgWmFjaGFyeSIsIk%2FigJlEb25uZWxsLUx1cmlhLCBBbm5lIEguIiwiTWluaWtlbCwgRXJpYyBWYWxsYWJoIiwiV2Vpc2J1cmQsIEJlbiIsIkxlaywgTW9ua29sIiwiV2FyZSwgSmFtZXMgUy4iLCJWaXR0YWwsIENocmlzdG9waGVyIiwiQXJtZWFuLCBJcmluYSBNLiIsIkJlcmdlbHNvbiwgTG91aXMiLCJDaWJ1bHNraXMsIEtyaXN0aWFuIiwiQ29ubm9sbHksIEtyaXN0ZW4gTS4iLCJDb3ZhcnJ1YmlhcywgTWlndWVsIiwiRG9ubmVsbHksIFN0YWNleSIsIkZlcnJpZXJhLCBTdGV2ZW4iLCJHYWJyaWVsLCBTdGFjZXkiLCJHZW50cnksIEplZmYiLCJHdXB0YSwgTmFtcmF0YSIsIkplYW5kZXQsIFRoaWJhdWx0IiwiS2FwbGFuLCBEaWFuZSIsIkxsYW53YXJuZSwgQ2hyaXN0b3BoZXIiLCJNdW5zaGksIFJ1Y2hpIiwiTm92b2QsIFNhbSIsIlBldHJpbGxvLCBOaWtlbGxlIiwiUm9hemVuLCBEYXZpZCIsIlJ1YW5vLVJ1YmlvLCBWYWxlbnRpbiIsIlNhbHR6bWFuLCBBbmRyZWEiLCJTY2hsZWljaGVyLCBNb2xseSIsIlNvdG8sIEpvc2UiLCJUaWJiZXR0cywgS2F0aGxlZW4iLCJUb2xvbmVuLCBDaGFybG90dGUiLCJXYWRlLCBHb3Jkb24iLCJUYWxrb3dza2ksIE1pY2hhZWwgRS4iLCJTYWxpbmFzLCBDYXJsb3MgQS4gQWd1aWxhciIsIkFobWFkLCBUYXJpcSIsIkFsYmVydCwgQ2hyaXN0aW5lIE0uIiwiQXJkaXNzaW5vLCBEaWVnbyIsIkF0em1vbiwgR2lsIiwiQmFybmFyZCwgSm9obiIsIkJlYXVnZXJpZSwg), as well as the dbNSFP database (v4.2)[^14^](https://web.endnote.com/citations/eyJkaXNwbGF5VGV4dCI6IjE0IiwiY2l0YXRpb25zIjpbeyJiaWJsaW9Db250ZW50IjpbeyJyZWZlcmVuY2VUeXBlIjoiMTciLCJ2b2x1bWUiOiIxMiIsInJzeG1sIjoiPHJlY29yZD48cmVmLXR5cGU%2BMTc8L3JlZi10eXBlPjxjb250cmlidXRvcnM%2BPGF1dGhvcnM%2BPGF1dGhvcj5MaXUsIFhpYW9taW5nPC9hdXRob3I%2BPGF1dGhvcj5MaSwgQ2hhbmc8L2F1dGhvcj48YXV0aG9yPk1vdSwgQ2hlbmdjaGVuZzwvYXV0aG9yPjxhdXRob3I%2BRG9uZywgWWlibzwvYXV0aG9yPjxhdXRob3I%2BVHUsIFlpY2hlbmc8L2F1dGhvcj48L2F1dGhvcnM%2BPC9jb250cmlidXRvcnM%2BPHRpdGxlcz48dGl0bGU%2BZGJOU0ZQIHY0OiBhIGNvbXByZWhlbnNpdmUgZGF0YWJhc2Ugb2YgdHJhbnNjcmlwdC1zcGVjaWZpYyBmdW5jdGlvbmFsIHByZWRpY3Rpb25zIGFuZCBhbm5vdGF0aW9ucyBmb3IgaHVtYW4gbm9uc3lub255bW91cyBhbmQgc3BsaWNlLXNpdGUgU05WczwvdGl0bGU%2BPHNlY29uZGFyeS10aXRsZT5HZW5vbWUgTWVkaWNpbmU8L3NlY29uZGFyeS10aXRsZT48L3RpdGxlcz48ZGF0ZXM%2BPHllYXI%2BMjAyMDwveWVhcj48L2RhdGVzPjx2b2x1bWU%2BMTI8L3ZvbHVtZT48aXNibj4xNzU2LTk5NFg8L2lzYm4%2BPGVsZWN0cm9uaWMtcmVzb3VyY2UtbnVtPjxzdHlsZSBmYWNlPVwidW5kZXJsaW5lXCI%2BMTAuMTE4Ni9zMTMwNzMtMDIwLTAwODAzLTk8L3N0eWxlPjwvZWxlY3Ryb25pYy1yZXNvdXJjZS1udW0%2BPG51bWJlcj4xPC9udW1iZXI%2BPHJlYy1ndWlkPjE2MjgxNDQxLWVkYTYtNGQ1Yi05YmZiLWI0YzdkMDk0YTYxNTwvcmVjLWd1aWQ%2BPHJlYy11c24%2BMzA8L3JlYy11c24%2BPC9yZWNvcmQ%2BIiwiYXV0aG9ycyI6WyJMaXUsIFhpYW9taW5nIiwiTGksIENoYW5nIiwiTW91LCBDaGVuZ2NoZW5nIiwiRG9uZywgWWlibyIsIlR1LCBZaWNoZW5nIl0sImlzYm4iOiIxNzU2LTk5NFgiLCJndWlkIjoiMTYyODE0NDEtZWRhNi00ZDViLTliZmItYjRjN2QwOTRhNjE1IiwieWVhciI6IjIwMjAiLCJudW1iZXIiOiIxIiwidGl0bGUiOiJkYk5TRlAgdjQ6IGEgY29tcHJlaGVuc2l2ZSBkYXRhYmFzZSBvZiB0cmFuc2NyaXB0LXNwZWNpZmljIGZ1bmN0aW9uYWwgcHJlZGljdGlvbnMgYW5kIGFubm90YXRpb25zIGZvciBodW1hbiBub25zeW5vbnltb3VzIGFuZCBzcGxpY2Utc2l0ZSBTTlZzIiwiZ3JvdXBHdWlkcyI6W10sImVsZWN0cm9uaWNSZXNvdXJjZU51bWJlciI6IjEwLjExODYvczEzMDczLTAyMC0wMDgwMy05IiwicmVjb3JkU3RhdHVzIjoiYWN0aXZlIiwic2Vjb25kYXJ5VGl0bGUiOiJHZW5vbWUgTWVkaWNpbmUifV0sInJlY29yZCI6eyJ2b2x1bWUiOiIxMiIsImlzYm4iOiIxNzU2LTk5NFgiLCJ0aXRsZXMiOnsidGl0bGUiOiJkYk5TRlAgdjQ6IGEgY29tcHJlaGVuc2l2ZSBkYXRhYmFzZSBvZiB0cmFuc2NyaXB0LXNwZWNpZmljIGZ1bmN0aW9uYWwgcHJlZGljdGlvbnMgYW5kIGFubm90YXRpb25zIGZvciBodW1hbiBub25zeW5vbnltb3VzIGFuZCBzcGxpY2Utc2l0ZSBTTlZzIiwic2Vj), were used to annotate genetic variants. We first used VEP to identify rare coding variants affecting the ENSEMBL canonical gene transcripts. From these variants, high-confidence loss-of-function (LoF) variants, including protein-truncating variants and high-impact splice variants, were identified using LOFTEE; we distinguished and analyzed two broad groups of LoF variants, one irrespective of potential flags, and a more stringent group excluding LoFs with any flags (LoFnoflag). Such LOFTEE flags may include variants affecting NAGNAG sites, variants affecting noncanonical splice sites, variants affecting non-conserved exons, or variants affecting single-exon transcripts.

Missense variants were also identified using VEP, and functional bioinformatic-predicted consequences were then inferred using various prediction tools. For the REVEL prediction tool[^15^](https://web.endnote.com/citations/eyJkaXNwbGF5VGV4dCI6IjE1IiwiY2l0YXRpb25zIjpbeyJiaWJsaW9Db250ZW50IjpbeyJyZWZlcmVuY2VUeXBlIjoiMTciLCJwYWdlcyI6Ijg3Ny04ODUiLCJ2b2x1bWUiOiI5OSIsInJzeG1sIjoiPHJlY29yZD48cmVmLXR5cGU%2BMTc8L3JlZi10eXBlPjxjb250cmlidXRvcnM%2BPGF1dGhvcnM%2BPGF1dGhvcj5Jb2FubmlkaXMsIE5pbGFoIE0uPC9hdXRob3I%2BPGF1dGhvcj5Sb3Roc3RlaW4sIEpvc2VwaCBILjwvYXV0aG9yPjxhdXRob3I%2BUGVqYXZlciwgVmlrYXM8L2F1dGhvcj48YXV0aG9yPk1pZGRoYSwgU3VtaXQ8L2F1dGhvcj48YXV0aG9yPk1jZG9ubmVsbCwgU2hhbm5vbiBLLjwvYXV0aG9yPjxhdXRob3I%2BQmFoZXRpLCBTYXVyYWJoPC9hdXRob3I%2BPGF1dGhvcj5NdXNvbGYsIEFudGhvbnk8L2F1dGhvcj48YXV0aG9yPkxpLCBRaW5nPC9hdXRob3I%2BPGF1dGhvcj5Ib2x6aW5nZXIsIEVtaWx5PC9hdXRob3I%2BPGF1dGhvcj5LYXJ5YWRpLCBEYW5pZWxsZTwvYXV0aG9yPjxhdXRob3I%2BQ2Fubm9uLUFsYnJpZ2h0LCBMaXNhIEEuPC9hdXRob3I%2BPGF1dGhvcj5UZWVybGluaywgQ3JhaWcgQy48L2F1dGhvcj48YXV0aG9yPlN0YW5mb3JkLCBKYW5ldCBMLjwvYXV0aG9yPjxhdXRob3I%2BSXNhYWNzLCBXaWxsaWFtIEIuPC9hdXRob3I%2BPGF1dGhvcj5YdSwgSmlhbmZlbmc8L2F1dGhvcj48YXV0aG9yPkNvb25leSwgS2F0aGxlZW4gQS48L2F1dGhvcj48YXV0aG9yPkxhbmdlLCBFdGhhbiBNLjwvYXV0aG9yPjxhdXRob3I%2BU2NobGV1dGtlciwgSm9oYW5uYTwvYXV0aG9yPjxhdXRob3I%2BQ2FycHRlbiwgSm9obiBELjwvYXV0aG9yPjxhdXRob3I%2BUG93ZWxsLCBJc2FhYyBKLjwvYXV0aG9yPjxhdXRob3I%2BQ3Vzc2Vub3QsIE9saXZpZXI8L2F1dGhvcj48YXV0aG9yPkNhbmNlbC1UYXNzaW4sIEdlcmFsZGluZTwvYXV0aG9yPjxhdXRob3I%2BR2lsZXMsIEdyYWhhbSBHLjwvYXV0aG9yPjxhdXRob3I%2BTWFjaW5uaXMsIFJvYmVydCBKLjwvYXV0aG9yPjxhdXRob3I%2BTWFpZXIsIENocmlzdGlhbmU8L2F1dGhvcj48YXV0aG9yPkhzaWVoLCBDaGloLUxpbjwvYXV0aG9yPjxhdXRob3I%2BV2lrbHVuZCwgRnJlZHJpazwvYXV0aG9yPjxhdXRob3I%2BQ2F0YWxvbmEsIFdpbGxpYW0gSi48L2F1dGhvcj48YXV0aG9yPkZvdWxrZXMsIFdpbGxpYW0gRC48L2F1dGhvcj48YXV0aG9yPk1hbmRhbCwgRGlwdGFzcmk8L2F1dGhvcj48YXV0aG9yPkVlbGVzLCBSb3NhbGluZCBBLjwvYXV0aG9yPjxhdXRob3I%2BS290ZS1KYXJhaSwgWnNvZmlhPC9hdXRob3I%2BPGF1dGhvcj5CdXN0YW1hbnRlLCBDYXJsb3MgRC48L2F1dGhvcj48YXV0aG9yPlNjaGFpZCwgRGFuaWVsIEouPC9hdXRob3I%2BPGF1dGhvcj5IYXN0aWUsIFRyZXZvcjwvYXV0aG9yPjxhdXRob3I%2BT3N0cmFuZGVyLCBFbGFpbmUgQS48L2F1dGhvcj48YXV0aG9yPkJhaWxleS1XaWxzb24sIEpvYW4gRS48L2F1dGhvcj48YXV0aG9yPlJhZGl2b2phYywgUHJlZHJhZzwvYXV0aG9yPjxhdXRob3I%2BVGhpYm9kZWF1LCBTdGVw), we used the variant prediction associated with the canonical transcript annotated through the dbNSFP database; we considered missense variants with a REVEL score of >=0.7 as predicted-damaging missense-REVEL variants.

We also curated additional, newer bioinformatic prediction tools. We downloaded the AlphaMissense predictions[^16^](https://web.endnote.com/citations/eyJkaXNwbGF5VGV4dCI6IjE2IiwiY2l0YXRpb25zIjpbeyJyZWNvcmQiOnsidGl0bGVzIjp7InNlY29uZGFyeS10aXRsZSI6IlNjaWVuY2UiLCJ0aXRsZSI6IkFjY3VyYXRlIHByb3Rlb21lLXdpZGUgbWlzc2Vuc2UgdmFyaWFudCBlZmZlY3QgcHJlZGljdGlvbiB3aXRoIEFscGhhTWlzc2Vuc2UifSwiZWxlY3Ryb25pYy1yZXNvdXJjZS1udW0iOiIxMC4xMTI2L3NjaWVuY2UuYWRnNzQ5MiIsImlzYm4iOiIwMDM2LTgwNzUiLCJ2b2x1bWUiOiIzODEiLCJjb250cmlidXRvcnMiOnsiYXV0aG9ycyI6eyJhdXRob3IiOlsiQ2hlbmcsIEp1biIsIk5vdmF0aSwgR3VpZG8iLCJQYW4sIEpvc2h1YSIsIkJ5Y3JvZnQsIENsYXJlIiwixb1lbWd1bHl0xJcsIEFrdmlsxJciLCJBcHBsZWJhdW0sIFRheWxvciIsIlByaXR6ZWwsIEFsZXhhbmRlciIsIldvbmcsIExhaSBIb25nIiwiWmllbGluc2tpLCBNaWNoYWwiLCJTYXJnZWFudCwgVG9iaWFzIiwiU2NobmVpZGVyLCBSb3NhbGlhIEcuIiwiU2VuaW9yLCBBbmRyZXcgVy4iLCJKdW1wZXIsIEpvaG4iLCJIYXNzYWJpcywgRGVtaXMiLCJLb2hsaSwgUHVzaG1lZXQiLCJBdnNlYywgxb1pZ2EiXX19LCJudW1iZXIiOiI2NjY0IiwicmVjLWd1aWQiOiJhZDZjYThhMC1lYmE2LTRjNzEtYThkNS1hNjZkYTYwMDFlODYiLCJyZWMtdXNuIjoiOCIsImRhdGVzIjp7InllYXIiOiIyMDIzIn0sInJlZi10eXBlIjoiMTcifSwiZ3VpZCI6ImFkNmNhOGEwLWViYTYtNGM3MS1hOGQ1LWE2NmRhNjAwMWU4NiIsImJpYmxpb0NvbnRlbnQiOlt7Imdyb3VwR3VpZHMiOltdLCJlbGVjdHJvbmljUmVzb3VyY2VOdW1iZXIiOiIxMC4xMTI2L3NjaWVuY2UuYWRnNzQ5MiIsInJlY29yZFN0YXR1cyI6ImFjdGl2ZSIsInNlY29uZGFyeVRpdGxlIjoiU2NpZW5jZSIsInJlZmVyZW5jZVR5cGUiOiIxNyIsInZvbHVtZSI6IjM4MSIsInJzeG1sIjoiPHJlY29yZD48cmVmLXR5cGU%2BMTc8L3JlZi10eXBlPjxjb250cmlidXRvcnM%2BPGF1dGhvcnM%2BPGF1dGhvcj5DaGVuZywgSnVuPC9hdXRob3I%2BPGF1dGhvcj5Ob3ZhdGksIEd1aWRvPC9hdXRob3I%2BPGF1dGhvcj5QYW4sIEpvc2h1YTwvYXV0aG9yPjxhdXRob3I%2BQnljcm9mdCwgQ2xhcmU8L2F1dGhvcj48YXV0aG9yPsW9ZW1ndWx5dMSXLCBBa3ZpbMSXPC9hdXRob3I%2BPGF1dGhvcj5BcHBsZWJhdW0sIFRheWxvcjwvYXV0aG9yPjxhdXRob3I%2BUHJpdHplbCwgQWxleGFuZGVyPC9hdXRob3I%2BPGF1dGhvcj5Xb25nLCBMYWkgSG9uZzwvYXV0aG9yPjxhdXRob3I%2BWmllbGluc2tpLCBNaWNoYWw8L2F1dGhvcj48YXV0aG9yPlNhcmdlYW50LCBUb2JpYXM8L2F1dGhvcj48YXV0aG9yPlNjaG5laWRlciwgUm9zYWxpYSBHLjwvYXV0aG9yPjxhdXRob3I%2BU2VuaW9yLCBBbmRyZXcgVy48L2F1dGhvcj48YXV0aG9yPkp1bXBlciwgSm9objwvYXV0aG9yPjxhdXRob3I%2BSGFzc2FiaXMsIERlbWlzPC9hdXRob3I%2BPGF1dGhvcj5Lb2hsaSwgUHVzaG1lZXQ8L2F1dGhvcj48YXV0aG9yPkF2c2VjLCDFvWlnYTwvYXV0aG9yPjwvYXV0aG9ycz48L2Nv), using the ‘AlphaMissense_hg38.tsv.gz’ file released in August 2023. We used gencode v45 to map the ENSEMBL transcript IDs to ENSEMBL gene IDs. We found discrepancies between the transcript used by the developers, and the ENSEMBL canonical transcript; as such, we mapped the variant predictions to variants affecting the canonical transcript from the VEP output, using the ENSEMBL gene IDs. We considered missense variants with ‘predicted_pathogenic’ classification by AlphaMissense as being predicted-damaging missense-AM variants.

We similarly received predictions for the PrimateAI-3D (PAI3D) tool[^17^](https://web.endnote.com/citations/eyJkaXNwbGF5VGV4dCI6IjE3IiwiY2l0YXRpb25zIjpbeyJiaWJsaW9Db250ZW50IjpbeyJndWlkIjoiYjdjOTcyMWUtZDJhZC00NWVkLTk4NzktY2ZkOTAxYWNlYmE0IiwidGl0bGUiOiJUaGUgbGFuZHNjYXBlIG9mIHRvbGVyYXRlZCBnZW5ldGljIHZhcmlhdGlvbiBpbiBodW1hbnMgYW5kIHByaW1hdGVzIiwieWVhciI6IjIwMjMiLCJudW1iZXIiOiI2NjQ4IiwicnN4bWwiOiI8cmVjb3JkPjxyZWYtdHlwZT4xNzwvcmVmLXR5cGU%2BPGNvbnRyaWJ1dG9ycz48YXV0aG9ycz48YXV0aG9yPkdhbywgSG9uZzwvYXV0aG9yPjxhdXRob3I%2BSGFtcCwgVG9iaWFzPC9hdXRob3I%2BPGF1dGhvcj5FZGUsIEplZmZyZXk8L2F1dGhvcj48YXV0aG9yPlNjaHJhaWJlciwgSm9zaHVhIEcuPC9hdXRob3I%2BPGF1dGhvcj5NY3JhZSwgSmVyZW15PC9hdXRob3I%2BPGF1dGhvcj5TaW5nZXItQmVyaywgTW9yaWVsPC9hdXRob3I%2BPGF1dGhvcj5ZYW5nLCBZYW5zaGVuPC9hdXRob3I%2BPGF1dGhvcj5EaWV0cmljaCwgQW5hc3Rhc2lhIFMuIEQuPC9hdXRob3I%2BPGF1dGhvcj5GaXppZXYsIFBldGtvIFAuPC9hdXRob3I%2BPGF1dGhvcj5LdWRlcm5hLCBMdWthcyBGLiBLLjwvYXV0aG9yPjxhdXRob3I%2BU3VuZGFyYW0sIExha3NzaG1hbjwvYXV0aG9yPjxhdXRob3I%2BV3UsIFlpYmluZzwvYXV0aG9yPjxhdXRob3I%2BQWRoaWthcmksIEFhc2hpc2g8L2F1dGhvcj48YXV0aG9yPkZpZWxkLCBZYWlyPC9hdXRob3I%2BPGF1dGhvcj5DaGVuLCBDaGVuPC9hdXRob3I%2BPGF1dGhvcj5CYXR6b2dsb3UsIFNlcmFmaW08L2F1dGhvcj48YXV0aG9yPkFndWV0LCBGcmFuY29pczwvYXV0aG9yPjxhdXRob3I%2BTGVtaXJlLCBHYWJyaWVsbGU8L2F1dGhvcj48YXV0aG9yPlJlaW1lcnMsIFJlYmVjY2E8L2F1dGhvcj48YXV0aG9yPkJhbGljaywgRGFuaWVsPC9hdXRob3I%2BPGF1dGhvcj5KYW5pYWssIE1hcmVpa2UgQy48L2F1dGhvcj48YXV0aG9yPkt1aGx3aWxtLCBNYXJ0aW48L2F1dGhvcj48YXV0aG9yPk9ya2luLCBKb3NlcGggRC48L2F1dGhvcj48YXV0aG9yPk1hbnUsIFNoaXZha3VtYXJhPC9hdXRob3I%2BPGF1dGhvcj5WYWxlbnp1ZWxhLCBBbGVqYW5kcm88L2F1dGhvcj48YXV0aG9yPkJlcmdtYW4sIEp1cmFqPC9hdXRob3I%2BPGF1dGhvcj5Sb3Vzc2VsbGUsIE1hcmpvbGFpbmU8L2F1dGhvcj48YXV0aG9yPlNpbHZhLCBGZWxpcGUgRW5uZXM8L2F1dGhvcj48YXV0aG9yPkFndWVkYSwgTGlkaWE8L2F1dGhvcj48YXV0aG9yPkJsYW5jLCBKdWxpZTwvYXV0aG9yPjxhdXRob3I%2BR3V0LCBNYXJ0YTwvYXV0aG9yPjxhdXRob3I%2BVnJpZXMsIERvcmllbiBEZTwvYXV0aG9yPjxhdXRob3I%2BR29vZGhlYWQsIElhbjwvYXV0aG9yPjxhdXRob3I%2BSGFycmlzLCBSLiBBbGFuPC9hdXRob3I%2BPGF1dGhvcj5SYXZlZW5kcmFuLCBNdXRodXN3YW15PC9hdXRob3I%2BPGF1dGhvcj5KZW5zZW4sIEF4ZWw8L2F1dGhvcj48YXV0aG9yPkNodW1hLCBJZHJpc3MgUy48L2F1dGhvcj48YXV0aG9yPkhvcnZhdGgsIEp1bGllIEUuPC), by requesting access in April 2024 (we received the ‘PrimateAI-3D_scores.csv.gz’ file). As above, we used gencode to map transcripts to ENSEMBL gene IDs and used these to match predictions to canonical gene transcripts from VEP. We considered missense variants with PAI3D score >=0.803 as predicted-damaging missense-PAI3D variants.

We also downloaded the popEVE predictions[^18^](https://web.endnote.com/citations/eyJkaXNwbGF5VGV4dCI6IjE4IiwiY2l0YXRpb25zIjpbeyJndWlkIjoiN2E3ZmY3ZmMtYzk5OC00YjQ3LTliNjUtY2UxYThmMTNhMGRiIiwicmVjb3JkIjp7ImVsZWN0cm9uaWMtcmVzb3VyY2UtbnVtIjp7InN0eWxlIjp7IiMiOiIxMC4xMDM4L3M0MTU4OC0wMjUtMDI0MDAtMSIsIkBmYWNlIjoidW5kZXJsaW5lIn19LCJ0aXRsZXMiOnsidGl0bGUiOiJQcm90ZW9tZS13aWRlIG1vZGVsIGZvciBodW1hbiBkaXNlYXNlIGdlbmV0aWNzIiwic2Vjb25kYXJ5LXRpdGxlIjoiTmF0dXJlIEdlbmV0aWNzIn0sImlzYm4iOiIxMDYxLTQwMzYiLCJ2b2x1bWUiOiI1NyIsInBhZ2VzIjoiMzE2NS0zMTc0IiwiY29udHJpYnV0b3JzIjp7ImF1dGhvcnMiOnsiYXV0aG9yIjpbIk9yZW5idWNoLCBSb3NlIiwiU2hlYXJlciwgQ291cnRuZXkgQS4iLCJLb2xsYXNjaCwgQWFyb24gVy4iLCJTcGlubmVyLCBBdml2IEQuIiwiSG9wZiwgVGhvbWFzIiwiTmlla2VyaywgTG9vZCBWYW4iLCJGcmFuY2VzY2hpLCBEaW5rbyIsIkRpYXMsIE1hZmFsZGEiLCJGcmF6ZXIsIEpvbmF0aGFuIiwiTWFya3MsIERlYm9yYSBTLiJdfX0sInJlYy11c24iOiIxMSIsIm51bWJlciI6IjEyIiwicmVjLWd1aWQiOiI3YTdmZjdmYy1jOTk4LTRiNDctOWI2NS1jZTFhOGYxM2EwZGIiLCJkYXRlcyI6eyJ5ZWFyIjoiMjAyNSJ9LCJyZWYtdHlwZSI6IjE3In0sImJpYmxpb0NvbnRlbnQiOlt7Imdyb3VwR3VpZHMiOltdLCJlbGVjdHJvbmljUmVzb3VyY2VOdW1iZXIiOiIxMC4xMDM4L3M0MTU4OC0wMjUtMDI0MDAtMSIsInJlY29yZFN0YXR1cyI6ImFjdGl2ZSIsInNlY29uZGFyeVRpdGxlIjoiTmF0dXJlIEdlbmV0aWNzIiwicnN4bWwiOiI8cmVjb3JkPjxyZWYtdHlwZT4xNzwvcmVmLXR5cGU%2BPGNvbnRyaWJ1dG9ycz48YXV0aG9ycz48YXV0aG9yPk9yZW5idWNoLCBSb3NlPC9hdXRob3I%2BPGF1dGhvcj5TaGVhcmVyLCBDb3VydG5leSBBLjwvYXV0aG9yPjxhdXRob3I%2BS29sbGFzY2gsIEFhcm9uIFcuPC9hdXRob3I%2BPGF1dGhvcj5TcGlubmVyLCBBdml2IEQuPC9hdXRob3I%2BPGF1dGhvcj5Ib3BmLCBUaG9tYXM8L2F1dGhvcj48YXV0aG9yPk5pZWtlcmssIExvb2QgVmFuPC9hdXRob3I%2BPGF1dGhvcj5GcmFuY2VzY2hpLCBEaW5rbzwvYXV0aG9yPjxhdXRob3I%2BRGlhcywgTWFmYWxkYTwvYXV0aG9yPjxhdXRob3I%2BRnJhemVyLCBKb25hdGhhbjwvYXV0aG9yPjxhdXRob3I%2BTWFya3MsIERlYm9yYSBTLjwvYXV0aG9yPjwvYXV0aG9ycz48L2NvbnRyaWJ1dG9ycz48dGl0bGVzPjx0aXRsZT5Qcm90ZW9tZS13aWRlIG1vZGVsIGZvciBodW1hbiBkaXNlYXNlIGdlbmV0aWNzPC90aXRsZT48c2Vjb25kYXJ5LXRpdGxlPk5hdHVyZSBHZW5ldGljczwvc2Vjb25kYXJ5LXRpdGxlPjwvdGl0bGVzPjxkYXRlcz48eWVhcj4yMDI1PC95ZWFyPjwvZGF0ZXM%2BPHBhZ2VzPjMxNjUtMzE3NDwvcGFnZXM%2BPHZvbHVtZT41Nzwvdm9sdW1lPjxpc2JuPjEwNjEtNDAzNjwvaXNibj48ZWxlY3Ryb25pYy1yZXNvdXJjZS1udW0%2BPHN0eWxlIGZhY2U9XCJ1bm) from December 2023 (file ‘2023-12-19_popeve_bulk.tar.gz’). Since the popEVE predictions were linked to RefSeq protein IDs, we first mapped the RefSeq protein IDs to ENSEMBL gene IDs (using https://ftp.ncbi.nlm.nih.gov/pub/datasets/command-line/v2/linux-amd64/datasets). In contrast to AlphaMissense and PAI3D, popEVE predictions were on the amino-acid change level, instead of the variant level. We therefore mapped popEVE predictions to missense genetic variants, using the VEP-predicted protein changes. We considered missense variants with popEVE score < -5.056 as predicted-damaging missense-popEVE variants.

#### Supplementary Methods 7. Batch variables

We adjusted for ‘genotype center’ and ‘sample source’ variables in all genetic association analyses. In AoU, Baylor College of Medicine, Broad Institute, and University of Washington performed genotyping using the same protocol for library construction (PCR Free Kapa HyperPrep), sequencer (NovaSeq 6000), software (DRAGEN v3.7.8), and software configuration. This was chosen in particular given slight differences in the sample selection might cause batch effects. The sample source includes blood or saliva. Previous assessment showed that there were regions where batch effects were caused by the sample source (<https://support.researchallofus.org/hc/en-us/articles/40462586515732-Additional-benchmarking-and-quality-analyses-on-the-All-of-Us-short-read-WGS-SNP-Indel-dataset>). Hence, we included the genotype center and sample source as covariates for all association analyses.

#### Supplementary Methods 8. Definition of known/novel loci in GWAS

We downloaded the Pan-UKB[^19^](https://web.endnote.com/citations/eyJkaXNwbGF5VGV4dCI6IjE5IiwiY2l0YXRpb25zIjpbeyJiaWJsaW9Db250ZW50IjpbeyJhdXRob3JzIjpbIkthcmN6ZXdza2ksIEtvbnJhZCBKLiIsIkd1cHRhLCBSYWh1bCIsIkthbmFpLCBNYXNhaGlybyIsIkx1LCBXZW5oYW4iLCJUc3VvLCBLcmlzdGluIiwiV2FuZywgWWluZyIsIldhbHRlcnMsIFJheW1vbmQgSy4iLCJUdXJsZXksIFBhdHJpY2siLCJDYWxsaWVyLCBTaGF3bmVlcXVhIiwiU2hhaCwgTmlyYXYgTi4iLCJCYXlhLCBOaWtvbGFzIiwiUGFsbWVyLCBEdW5jYW4gUy4iLCJHb2xkc3RlaW4sIEphY3F1ZWxpbmUgSS4iLCJTYXJtYSwgR29wYWwiLCJTb2xvbW9uc29uLCBNYXR0aGV3IiwiQ2hlbmcsIE5hdGhhbiIsIkJyeWFudCwgU2FtIiwiQ2h1cmNoaG91c2UsIENsYWlyZSIsIkN1c2ljaywgQ2Fyb2xpbmUgTS4iLCIuLi4iLCJNYXJ0aW4sIEFsaWNpYSBSLiJdLCJpc2JuIjoiMTA2MS00MDM2Iiwidm9sdW1lIjoiNTciLCJyc3htbCI6IjxyZWNvcmQ%2BPHJlZi10eXBlPjE3PC9yZWYtdHlwZT48Y29udHJpYnV0b3JzPjxhdXRob3JzPjxhdXRob3I%2BS2FyY3pld3NraSwgS29ucmFkIEouPC9hdXRob3I%2BPGF1dGhvcj5HdXB0YSwgUmFodWw8L2F1dGhvcj48YXV0aG9yPkthbmFpLCBNYXNhaGlybzwvYXV0aG9yPjxhdXRob3I%2BTHUsIFdlbmhhbjwvYXV0aG9yPjxhdXRob3I%2BVHN1bywgS3Jpc3RpbjwvYXV0aG9yPjxhdXRob3I%2BV2FuZywgWWluZzwvYXV0aG9yPjxhdXRob3I%2BV2FsdGVycywgUmF5bW9uZCBLLjwvYXV0aG9yPjxhdXRob3I%2BVHVybGV5LCBQYXRyaWNrPC9hdXRob3I%2BPGF1dGhvcj5DYWxsaWVyLCBTaGF3bmVlcXVhPC9hdXRob3I%2BPGF1dGhvcj5TaGFoLCBOaXJhdiBOLjwvYXV0aG9yPjxhdXRob3I%2BQmF5YSwgTmlrb2xhczwvYXV0aG9yPjxhdXRob3I%2BUGFsbWVyLCBEdW5jYW4gUy48L2F1dGhvcj48YXV0aG9yPkdvbGRzdGVpbiwgSmFjcXVlbGluZSBJLjwvYXV0aG9yPjxhdXRob3I%2BU2FybWEsIEdvcGFsPC9hdXRob3I%2BPGF1dGhvcj5Tb2xvbW9uc29uLCBNYXR0aGV3PC9hdXRob3I%2BPGF1dGhvcj5DaGVuZywgTmF0aGFuPC9hdXRob3I%2BPGF1dGhvcj5CcnlhbnQsIFNhbTwvYXV0aG9yPjxhdXRob3I%2BQ2h1cmNoaG91c2UsIENsYWlyZTwvYXV0aG9yPjxhdXRob3I%2BQ3VzaWNrLCBDYXJvbGluZSBNLjwvYXV0aG9yPjxhdXRob3I%2BUG90ZXJiYSwgVGltb3RoeTwvYXV0aG9yPjxhdXRob3I%2BQ29tcGl0ZWxsbywgSm9objwvYXV0aG9yPjxhdXRob3I%2BS2luZywgRGFuaWVsPC9hdXRob3I%2BPGF1dGhvcj5aaG91LCBXZWk8L2F1dGhvcj48YXV0aG9yPlNlZWQsIENvdHRvbjwvYXV0aG9yPjxhdXRob3I%2BRmludWNhbmUsIEhpbGFyeSBLLjwvYXV0aG9yPjxhdXRob3I%2BRGFseSwgTWFyayBKLjwvYXV0aG9yPjxhdXRob3I%2BTmVhbGUsIEJlbmphbWluIE0uPC9hdXRob3I%2BPGF1dGhvcj5BdGtpbnNvbiwgRWxpemFiZXRoIEcuPC9hdXRob3I%2BPGF1dGhvcj5NYXJ0aW4sIEFsaWNpYSBSLjwvYXV0aG9yPjwvYXV0aG9ycz48L2NvbnRy) results for all ancestries, the height GWAS summary statistics from the GIANT consortium[^20^](https://web.endnote.com/citations/eyJkaXNwbGF5VGV4dCI6IjIwIiwiY2l0YXRpb25zIjpbeyJyZWNvcmQiOnsidm9sdW1lIjoiNjEwIiwiZWxlY3Ryb25pYy1yZXNvdXJjZS1udW0iOiIxMC4xMDM4L3M0MTU4Ni0wMjItMDUyNzUteSIsInRpdGxlcyI6eyJ0aXRsZSI6IkEgc2F0dXJhdGVkIG1hcCBvZiBjb21tb24gZ2VuZXRpYyB2YXJpYW50cyBhc3NvY2lhdGVkIHdpdGggaHVtYW4gaGVpZ2h0Iiwic2Vjb25kYXJ5LXRpdGxlIjoiTmF0dXJlIn0sImlzYm4iOiIwMDI4LTA4MzYiLCJjb250cmlidXRvcnMiOnsiYXV0aG9ycyI6eyJhdXRob3IiOlsiWWVuZ28sIExvw69jIiwiVmVkYW50YW0sIFNhaWxhamEiLCJNYXJvdWxpLCBFaXJpbmkiLCJTaWRvcmVua28sIEp1bGlhIiwiQmFydGVsbCwgRXJpYyIsIlNha2F1ZSwgU2FvcmkiLCJHcmFmZiwgTWFyaWVsaXNhIiwiRWxpYXNlbiwgQW5kZXJzIFUuIiwiSmlhbmcsIFl1bnh1YW4iLCJSYWdoYXZhbiwgU3JpZGhhcmFuIiwiTWlhbywgSmVua2FpIiwiQXJpYXMsIEpvc2h1YSBELiIsIkdyYWhhbSwgU2FyYWggRS4iLCJNdWthbWVsLCBSb25lbiBFLiIsIlNwcmFja2xlbiwgQ2Fzc2FuZHJhIE4uIiwiWWluLCBYaWFueW9uZyIsIkNoZW4sIFNoeWgtSHVlaSIsIkZlcnJlaXJhLCBUZXJlc2EiLCJIaWdobGFuZCwgSGVhdGhlciBILiIsIkppLCBZaW5namllIiwiS2FyYWRlcmksIFR1Z2NlIiwiTGluLCBLdWFuZyIsIkzDvGxsLCBLcmVldGUiLCJNYWxkZW4sIERlYm9yYWggRS4iLCJNZWRpbmEtR29tZXosIENhcm9saW5hIiwiTWFjaGFkbywgTW9hcmEiLCJNb29yZSwgQW15IiwiUsO8ZWdlciwgU2luYSIsIlNpbSwgWHVlbGluZyIsIlZyaWV6ZSwgU2NvdHQiLCJBaGx1d2FsaWEsIFRhcnVudmVlciBTLiIsIkFraXlhbWEsIE1hc2F0byIsIkFsbGlzb24sIE1hdHRoZXcgQS4iLCJBbHZhcmV6LCBNYXJjdXMiLCJBbmRlcnNlbiwgTWV0dGUgSy4iLCJBbmksIEFsaXJlemEiLCJBcHBhZHVyYWksIFZpdmVrIiwiQXJiZWV2YSwgTGl1Ym92IiwiQmhhc2thciwgU2VlbWEiLCJCaWVsYWssIExhd3JlbmNlIEYuIiwiQm9sbGVwYWxsaSwgU2FpbGFsaXRoYSIsIkJvbm55Y2FzdGxlLCBMb3JpIEwuIiwiQm9yay1KZW5zZW4sIEpldHRlIiwiQnJhZGZpZWxkLCBKb25hdGhhbiBQLiIsIkJyYWRmb3JkLCBZdWtpIiwiQnJhdW5kLCBQZXRlciBTLiIsIkJyb2R5LCBKZW5uaWZlciBBLiIsIkJ1cmdkb3JmLCBLcmlzdG9mZmVyIFMuIiwiQ2FkZSwgQnJpYW4gRS4iLCJDYWksIEh1aSIsIkNhaSwgUWl1eWluIiwiQ2FtcGJlbGwsIEFyY2hpZSIsIkNhw7FhZGFzLUdhcnJlLCBNYXJpc2EiLCJDYXRhbW8sIEV1bGFsaWEiLCJDaGFpLCBKaW4tRmFuZyIsIkNoYWksIFhpYW9yYW4iLCJDaGFuZywgTGktQ2hpbmciLCJDaGFuZywgWWktQ2hlbmciLCJDaGVuLCBDaGllbi1Ic2l1biIsIkNoZXNpLCBBbGVzc2FuZHJhIiwiQ2hvaSwgU2V1bmcgSG9hbiIsIkNodW5nLCBSZW4tSHVhIiwiQ29jY2EsIE1hc3NpbWlsaWFubyIsIkNvbmNhcywgTWFyaWEgUGluYSIsIkNvdXR1cmUsIENocmlzdGlhbiIsIkN1ZWxsYXItUGFydGlkYSwgR2), the lipid GWAS summary statistics from the Global Lipids Genetics Consortium[^21^](https://web.endnote.com/citations/eyJkaXNwbGF5VGV4dCI6IjIxIiwiY2l0YXRpb25zIjpbeyJyZWNvcmQiOnsibnVtYmVyIjoiNzg5MCIsInJlYy1ndWlkIjoiYjk4M2NlNGItMzQ2ZS00Mzg2LWJhZDgtM2U4MmUxZmQ0MTQ3IiwicmVjLXVzbiI6IjQ1IiwiZGF0ZXMiOnsieWVhciI6IjIwMjEifSwicmVmLXR5cGUiOiIxNyIsImVsZWN0cm9uaWMtcmVzb3VyY2UtbnVtIjoiaHR0cHM6Ly9kb2kub3JnLzEwLjEwMzgvczQxNTg2LTAyMS0wNDA2NC0zIiwidGl0bGVzIjp7InRpdGxlIjoiVGhlIHBvd2VyIG9mIGdlbmV0aWMgZGl2ZXJzaXR5IGluIGdlbm9tZS13aWRlIGFzc29jaWF0aW9uIHN0dWRpZXMgb2YgbGlwaWRzIiwic2Vjb25kYXJ5LXRpdGxlIjoiTmF0dXJlIn0sImlzYm4iOiIwMDI4LTA4MzYiLCJ2b2x1bWUiOiI2MDAiLCJwYWdlcyI6IjY3NS02NzkiLCJjb250cmlidXRvcnMiOnsiYXV0aG9ycyI6eyJhdXRob3IiOlsiR3JhaGFtLCBTYXJhaCBFLiIsIkNsYXJrZSwgU2hvYSBMLiIsIld1LCBLdWFuLUhhbiBILiIsIkthbm9uaSwgU3RhdnJvdWxhIiwiWmFqYWMsIEdyZWcgSi4gTS4iLCJSYW1kYXMsIFNod2V0YSIsIlN1cmFra2EsIElkYSIsIk50YWxsYSwgSW9hbm5hIiwiVmVkYW50YW0sIFNhaWxhamEiLCJXaW5rbGVyLCBUaG9tYXMgVy4iLCJMb2NrZSwgQWRhbSBFLiIsIk1hcm91bGksIEVpcmluaSIsIkh3YW5nLCBNaSBZZW9uZyIsIkhhbiwgU29oZWUiLCJOYXJpdGEsIEFraXJhIiwiQ2hvdWRodXJ5LCBBbmFueW8iLCJCZW50bGV5LCBBbXkgUi4iLCJFa29ydSwgS2VubmV0aCIsIlZlcm1hLCBBbnVyYWciLCJUcml2ZWRpLCBCaGF2aSIsIk1hcnRpbiwgSGlsYXJ5IEMuIiwiSHVudCwgS2FyZW4gQS4iLCJIdWksIFFpbiIsIktsYXJpbiwgRGVyZWsiLCJaaHUsIFhpYW5nIiwiVGhvcmxlaWZzc29uLCBHdWRtYXIiLCJIZWxnYWRvdHRpciwgQW5uYSIsIkd1ZGJqYXJ0c3NvbiwgRGFuaWVsIEYuIiwiSG9sbSwgSGlsbWEiLCJPbGFmc3NvbiwgSXNsZWlmdXIiLCJBa2l5YW1hLCBNYXNhdG8iLCJTYWthdWUsIFNhb3JpIiwiVGVyYW8sIENoaWthc2hpIiwiS2FuYWksIE1hc2FoaXJvIiwiWmhvdSwgV2VpIiwiQnJ1bXB0b24sIEJlbiBNLiIsIlJhc2hlZWQsIEh1bWFpcmEiLCJSdW90c2FsYWluZW4sIFNhbm5pIEUuIiwiSGF2dWxpbm5hLCBBa2kgUy4iLCJWZXR1cmksIFlvZ2FzdWRoYSIsIkZlbmcsIFFpcGluZyIsIlJvc2VudGhhbCwgRWxpc2FiZXRoIEEuIiwiTGluZ3JlbiwgVG9kZCIsIlBhY2hlY28sIEplbm5pZmVyIEFsbGVuIiwiUGVuZGVyZ3Jhc3MsIFNhcmFoIEEuIiwiSGFlc3NsZXIsIEplZmZyZXkiLCJHaXVsaWFuaW5pLCBGcmFuY28iLCJCcmFkZm9yZCwgWXVraSIsIk1pbGxlciwgSmFzb24gRS4iLCJDYW1wYmVsbCwgQXJjaGllIiwiTGluLCBLdWFuZyIsIk1pbGx3b29kLCBJb25hIFkuIiwiSGluZHksIEdlb3JnZSIsIlJhc2hlZWQsIEFzaWYiLCJGYXVsLCBKZXNzaWNhIEQuIiwiWmhhbywgV2VpIiwiV2VpciwgRGF2aWQgUi4iLCJUdXJtYW4sIENvbnN0YW5jZSIsIkh1YW5nLCBIb25neWFuIiwiR3JhZmYsIE1hcmlhZW), and blood pressure GWAS summary statistics[^22^](https://web.endnote.com/citations/eyJkaXNwbGF5VGV4dCI6IjIyIiwiY2l0YXRpb25zIjpbeyJiaWJsaW9Db250ZW50IjpbeyJwYWdlcyI6Ijc3OC03OTEiLCJyZWZlcmVuY2VUeXBlIjoiMTciLCJhdXRob3JzIjpbIktlYXRvbiwgSmFjb2IgTS4iLCJLYW1hbGksIFpvaGEiLCJYaWUsIFRpYW4iLCJWYWV6LCBBaG1hZCIsIldpbGxpYW1zLCBBcmllbCIsIkdvbGV2YSwgU2xhdmluYSBCLiIsIkFuaSwgQWxpcmV6YSIsIkV2YW5nZWxvdSwgRXZhbmdlbG9zIiwiSGVsbHdlZ2UsIEphY2tseW4gTi4iLCJZZW5nbywgTG9pYyIsIllvdW5nLCBXaWxsaWFtIEouIiwiVHJheWxvciwgTWF0dGhldyIsIkdpcmksIEF5dXNoIiwiWmhlbmcsIFpoaWxpIiwiWmVuZywgSmlhbiIsIkNoYXNtYW4sIERhbmllbCBJLiIsIk1vcnJpcywgQW5kcmV3IFAuIiwiQ2F1bGZpZWxkLCBNYXJrIEouIiwiSHdhbmcsIFNoaWgtSmVuIiwiLi4uIiwiV2FycmVuLCBIZWxlbiBSLiJdLCJpc2JuIjoiMTA2MS00MDM2Iiwidm9sdW1lIjoiNTYiLCJyc3htbCI6IjxyZWNvcmQ%2BPHJlZi10eXBlPjE3PC9yZWYtdHlwZT48Y29udHJpYnV0b3JzPjxhdXRob3JzPjxhdXRob3I%2BS2VhdG9uLCBKYWNvYiBNLjwvYXV0aG9yPjxhdXRob3I%2BS2FtYWxpLCBab2hhPC9hdXRob3I%2BPGF1dGhvcj5YaWUsIFRpYW48L2F1dGhvcj48YXV0aG9yPlZhZXosIEFobWFkPC9hdXRob3I%2BPGF1dGhvcj5XaWxsaWFtcywgQXJpZWw8L2F1dGhvcj48YXV0aG9yPkdvbGV2YSwgU2xhdmluYSBCLjwvYXV0aG9yPjxhdXRob3I%2BQW5pLCBBbGlyZXphPC9hdXRob3I%2BPGF1dGhvcj5FdmFuZ2Vsb3UsIEV2YW5nZWxvczwvYXV0aG9yPjxhdXRob3I%2BSGVsbHdlZ2UsIEphY2tseW4gTi48L2F1dGhvcj48YXV0aG9yPlllbmdvLCBMb2ljPC9hdXRob3I%2BPGF1dGhvcj5Zb3VuZywgV2lsbGlhbSBKLjwvYXV0aG9yPjxhdXRob3I%2BVHJheWxvciwgTWF0dGhldzwvYXV0aG9yPjxhdXRob3I%2BR2lyaSwgQXl1c2g8L2F1dGhvcj48YXV0aG9yPlpoZW5nLCBaaGlsaTwvYXV0aG9yPjxhdXRob3I%2BWmVuZywgSmlhbjwvYXV0aG9yPjxhdXRob3I%2BQ2hhc21hbiwgRGFuaWVsIEkuPC9hdXRob3I%2BPGF1dGhvcj5Nb3JyaXMsIEFuZHJldyBQLjwvYXV0aG9yPjxhdXRob3I%2BQ2F1bGZpZWxkLCBNYXJrIEouPC9hdXRob3I%2BPGF1dGhvcj5Id2FuZywgU2hpaC1KZW48L2F1dGhvcj48YXV0aG9yPktvb25lciwgSmFzcGFsIFMuPC9hdXRob3I%2BPGF1dGhvcj5Db25lbiwgRGF2aWQ8L2F1dGhvcj48YXV0aG9yPkF0dGlhLCBKb2huIFIuPC9hdXRob3I%2BPGF1dGhvcj5Nb3JyaXNvbiwgQWxhbm5hIEMuPC9hdXRob3I%2BPGF1dGhvcj5Mb29zLCBSdXRoIEouIEYuPC9hdXRob3I%2BPGF1dGhvcj5LcmlzdGlhbnNzb24sIEthdGk8L2F1dGhvcj48YXV0aG9yPlNjaG1pZHQsIFJlaW5ob2xkPC9hdXRob3I%2BPGF1dGhvcj5IaWNrcywgQW5kcmV3IEEuPC9hdXRob3I%2BPGF1dGhvcj5QcmFtc3RhbGxlciwgUGV0ZXIgUC48L2F1dGhvcj48YXV0aG9yPk5lbHNvbiwgQ2hyaXN0b3BoZXIgUC48L2F1dGhvcj48YX). After downloading these files, we restricted them to variants with P < 5 × 10^-8^, followed by converting genome coordinates from GRCh37 to GRCh38 where applicable. We defined genome-wide significant loci by iteratively spanning the ±500 kb region around the most significant variant and merging overlapping regions until no genome-wide significant variants were detected within ±1 Mb. Also we downloaded the locus information from the Supplementary Tale 2, 3, and 6 in the Million Veteran Program (MVP) Phenome-Wide Association Study (PheWAS)[^2^](https://web.endnote.com/citations/eyJkaXNwbGF5VGV4dCI6IjIiLCJjaXRhdGlvbnMiOlt7ImJpYmxpb0NvbnRlbnQiOlt7ImF1dGhvcnMiOlsiVmVybWEsIEFudXJhZyIsIkh1ZmZtYW4sIEplbm5pZmVyIEUuIiwiUm9kcmlndWV6LCBBbGV4IiwiQ29uZXJ5LCBNaXRjaGVsbCIsIkxpdSwgTW9sZWkiLCJIbywgWXVrLUxhbSIsIktpbSwgWW91bmdkYWUiLCJIZWlzZSwgRGF2aWQgQS4iLCJHdWFyZSwgTGluZHNheSIsIlBhbmlja2FuLCBWaWR1bCBBeWFrdWxhbmdhcmEiLCJHYXJjb24sIEhlbGVuZSIsIkxpbmFyZXMsIEZyYW5jaWVsIiwiQ29zdGEsIExhdXJlbiIsIkdvZXRoZXJ0LCBJYW4iLCJUaXB0b24sIFJ5YW4iLCJIb25lcmxhdywgSmFjcXVlbGluZSIsIkRhdmllcywgTGF1cmEiLCJXaGl0Ym91cm5lLCBTdGFjZXkiLCJDb2hlbiwgSmVyZW15IiwiLi4uIiwiTGlhbywgS2F0aGVyaW5lIFAuIl0sImlzYm4iOiIwMDM2LTgwNzUiLCJ2b2x1bWUiOiIzODUiLCJyc3htbCI6IjxyZWNvcmQ%2BPHJlZi10eXBlPjE3PC9yZWYtdHlwZT48Y29udHJpYnV0b3JzPjxhdXRob3JzPjxhdXRob3I%2BVmVybWEsIEFudXJhZzwvYXV0aG9yPjxhdXRob3I%2BSHVmZm1hbiwgSmVubmlmZXIgRS48L2F1dGhvcj48YXV0aG9yPlJvZHJpZ3VleiwgQWxleDwvYXV0aG9yPjxhdXRob3I%2BQ29uZXJ5LCBNaXRjaGVsbDwvYXV0aG9yPjxhdXRob3I%2BTGl1LCBNb2xlaTwvYXV0aG9yPjxhdXRob3I%2BSG8sIFl1ay1MYW08L2F1dGhvcj48YXV0aG9yPktpbSwgWW91bmdkYWU8L2F1dGhvcj48YXV0aG9yPkhlaXNlLCBEYXZpZCBBLjwvYXV0aG9yPjxhdXRob3I%2BR3VhcmUsIExpbmRzYXk8L2F1dGhvcj48YXV0aG9yPlBhbmlja2FuLCBWaWR1bCBBeWFrdWxhbmdhcmE8L2F1dGhvcj48YXV0aG9yPkdhcmNvbiwgSGVsZW5lPC9hdXRob3I%2BPGF1dGhvcj5MaW5hcmVzLCBGcmFuY2llbDwvYXV0aG9yPjxhdXRob3I%2BQ29zdGEsIExhdXJlbjwvYXV0aG9yPjxhdXRob3I%2BR29ldGhlcnQsIElhbjwvYXV0aG9yPjxhdXRob3I%2BVGlwdG9uLCBSeWFuPC9hdXRob3I%2BPGF1dGhvcj5Ib25lcmxhdywgSmFjcXVlbGluZTwvYXV0aG9yPjxhdXRob3I%2BRGF2aWVzLCBMYXVyYTwvYXV0aG9yPjxhdXRob3I%2BV2hpdGJvdXJuZSwgU3RhY2V5PC9hdXRob3I%2BPGF1dGhvcj5Db2hlbiwgSmVyZW15PC9hdXRob3I%2BPGF1dGhvcj5Qb3NuZXIsIERhbmllbCBDLjwvYXV0aG9yPjxhdXRob3I%2BU2FuZ2FyLCBSYWh1bDwvYXV0aG9yPjxhdXRob3I%2BTXVycmF5LCBNaWNoYWVsPC9hdXRob3I%2BPGF1dGhvcj5XYW5nLCBYdWFuPC9hdXRob3I%2BPGF1dGhvcj5Eb2NodGVybWFubiwgRGFuaWVsIFIuPC9hdXRob3I%2BPGF1dGhvcj5EZXZpbmVuaSwgUG9vcm5pbWE8L2F1dGhvcj48YXV0aG9yPlNoaSwgWXVubGluZzwvYXV0aG9yPjxhdXRob3I%2BTmFuZGksIFRhcmFrIE5hdGg8L2F1dGhvcj48YXV0aG9yPkFzc2ltZXMsIFRoZW1pc3RvY2xlcyBMLjwvYXV0aG9yPjxhdXRob3I%2BQnJ1bmV0dGUsIENoYXJsZXMgQS48L2F1dGhvcj48YXV0aG9yPkNhcnJvbGwsIFJvYmVy), and from the Supplementary Table 1 and 7 in the meta-analysis of PheWAS[^23^](https://web.endnote.com/citations/eyJkaXNwbGF5VGV4dCI6IjIzIiwiY2l0YXRpb25zIjpbeyJiaWJsaW9Db250ZW50IjpbeyJlbGVjdHJvbmljUmVzb3VyY2VOdW1iZXIiOiIxMC4xMTAxLzIwMjUuMDQuMTguMjUzMjYwNzQiLCJncm91cEd1aWRzIjpbXSwicmVmZXJlbmNlVHlwZSI6IjEzIiwiYXV0aG9ycyI6WyJMZXZpbiwgTWljaGFlbCBHIiwiS295YW1hLCBTYXRvc2hpIiwiV29lcm5lciwgSmFrb2IiLCJaaGFuZywgRGF2aWQgWSIsIlJvZHJpZ3VleiwgQWxleGlzIiwiTmFuZGksIFRhcmFrIiwiVHJ1b25nLCBCdXUiLCJBYnJhbW93aXR6LCBTYXJhaCBBIiwiR3VwdGEsIEhyaXR2aWsiLCJLYW1pbmVuaSwgSGltYW5pIiwiSG9ybnNieSwgV2hpdG5leSIsIkxpLCBaaWxpbmdoYW4iLCJDb2hyb24sIFRheWxvciIsIkh1ZmZtYW4sIEplbm5pZmVyIEUiLCJFbGxpbm9yLCBQYXRyaWNrIiwiS2ltLCBEb2t5b29uIiwiTGlhbywgS2F0aGVyaW5lIFAiLCJNYWRkdXJpLCBSYXZpIEsiLCJWb2lnaHQsIEJlbmphbWluIEYiLCIuLi4iLCJOYXRhcmFqYW4sIFByYWRlZXAiXSwicnN4bWwiOiI8cmVjb3JkPjxyZWYtdHlwZT4xMzwvcmVmLXR5cGU%2BPGNvbnRyaWJ1dG9ycz48YXV0aG9ycz48YXV0aG9yPkxldmluLCBNaWNoYWVsIEc8L2F1dGhvcj48YXV0aG9yPktveWFtYSwgU2F0b3NoaTwvYXV0aG9yPjxhdXRob3I%2BV29lcm5lciwgSmFrb2I8L2F1dGhvcj48YXV0aG9yPlpoYW5nLCBEYXZpZCBZPC9hdXRob3I%2BPGF1dGhvcj5Sb2RyaWd1ZXosIEFsZXhpczwvYXV0aG9yPjxhdXRob3I%2BTmFuZGksIFRhcmFrPC9hdXRob3I%2BPGF1dGhvcj5UcnVvbmcsIEJ1dTwvYXV0aG9yPjxhdXRob3I%2BQWJyYW1vd2l0eiwgU2FyYWggQTwvYXV0aG9yPjxhdXRob3I%2BR3VwdGEsIEhyaXR2aWs8L2F1dGhvcj48YXV0aG9yPkthbWluZW5pLCBIaW1hbmk8L2F1dGhvcj48YXV0aG9yPkhvcm5zYnksIFdoaXRuZXk8L2F1dGhvcj48YXV0aG9yPkxpLCBaaWxpbmdoYW48L2F1dGhvcj48YXV0aG9yPkNvaHJvbiwgVGF5bG9yPC9hdXRob3I%2BPGF1dGhvcj5IdWZmbWFuLCBKZW5uaWZlciBFPC9hdXRob3I%2BPGF1dGhvcj5FbGxpbm9yLCBQYXRyaWNrPC9hdXRob3I%2BPGF1dGhvcj5LaW0sIERva3lvb248L2F1dGhvcj48YXV0aG9yPkxpYW8sIEthdGhlcmluZSBQPC9hdXRob3I%2BPGF1dGhvcj5NYWRkdXJpLCBSYXZpIEs8L2F1dGhvcj48YXV0aG9yPlZvaWdodCwgQmVuamFtaW4gRjwvYXV0aG9yPjxhdXRob3I%2BVmVybWEsIEFudXJhZzwvYXV0aG9yPjxhdXRob3I%2BRGFtcmF1ZXIsIFNjb3R0IE08L2F1dGhvcj48YXV0aG9yPk5hdGFyYWphbiwgUHJhZGVlcDwvYXV0aG9yPjwvYXV0aG9ycz48L2NvbnRyaWJ1dG9ycz48dGl0bGVzPjx0aXRsZT5HZW5vbWUtV2lkZSBBc3Nlc3NtZW50IG9mIFBsZWlvdHJvcHkgQWNyb3NzICZndDsxMDAwIFRyYWl0cyBmcm9tIEdsb2JhbCBCaW9iYW5rczwvdGl0bGU%2BPC90aXRsZXM%2BPGRhdGVzPjx5ZWFyPjIwMjU8L3llYXI%2BPC9kYXRlcz48ZWxlY3Ryb25pYy1yZXNvdXJjZS1udW0%2BMTAuMTEwMS8yMDI1LjA0). Finally we downloaded all variant-phenotype associations from the Open Targets Platform and GWAS catalog . We combined these dataset to define known loci-phenotype pairs. All URLs are shown in the **Supplementary Data 46**.

#### Supplementary Methods 9. Liftover from GRCh37 to GRCh38

We used the UCSC liftOver to convert genome coordinates from GRCh37 to GRCh38 using the input BED file. To implement the liftover, a chain format file was obtained from the website (**Supplementary Data 46**). The variants on alternative contigs were excluded.

#### Supplementary Methods 10. Phenotype construction for PheWAS

We used the R package PheWAS (v.1.0) to create disease phecodes mapped from various ICD-10 billing codes. We required at least one instance of an ICD code to define a sample as a case, while all other samples were considered controls for the given phecode. This analysis was restricted to individuals who gave consent for use of EHR. Prevalent and incident cases were pooled[^24^](https://web.endnote.com/citations/eyJkaXNwbGF5VGV4dCI6IjI0IiwiY2l0YXRpb25zIjpbeyJiaWJsaW9Db250ZW50IjpbeyJlbGVjdHJvbmljUmVzb3VyY2VOdW1iZXIiOiIxMC4xMDM4L3M0MTU4OC0wMjQtMDE4OTQtNSIsImdyb3VwR3VpZHMiOltdLCJzZWNvbmRhcnlUaXRsZSI6Ik5hdHVyZSBHZW5ldGljcyIsInJlY29yZFN0YXR1cyI6ImFjdGl2ZSIsImlzYm4iOiIxMDYxLTQwMzYiLCJhdXRob3JzIjpbIkp1cmdlbnMsIFNlYW4gSi4iLCJXYW5nLCBYaW4iLCJDaG9pLCBTZXVuZyBIb2FuIiwiV2VuZywgTHUtQ2hlbiIsIktveWFtYSwgU2F0b3NoaSIsIlBpcnJ1Y2NlbGxvLCBKYW1lcyBQLiIsIk5ndXllbiwgVHJhbmciLCJTbWFkYmVjaywgUGF0cmljayIsIkphbmcsIERvbmdrZXVuIiwiQ2hhZmZpbiwgTWFyayIsIldhbHNoLCBSb2RkeSIsIlJvc2VsbGksIENhcm9saW5hIiwiRWxsaW90dCwgQW1hbmRhIEwuIiwiV2lqZGV2ZWxkLCBMZW9ub29yIEYuIEouIE0uIiwiQmlkZGluZ2VyLCBLaXJhbiBKLiIsIkthbnksIFNoaW53YW4iLCJSw6Rtw7YsIEpvZWwgVC4iLCJOYXRhcmFqYW4sIFByYWRlZXAiLCJBcmFnYW0sIEtyaXNobmEgRy4iLCIuLi4iLCJFbGxpbm9yLCBQYXRyaWNrIFQuIl0sInJzeG1sIjoiPHJlY29yZD48cmVmLXR5cGU%2BMTc8L3JlZi10eXBlPjxjb250cmlidXRvcnM%2BPGF1dGhvcnM%2BPGF1dGhvcj5KdXJnZW5zLCBTZWFuIEouPC9hdXRob3I%2BPGF1dGhvcj5XYW5nLCBYaW48L2F1dGhvcj48YXV0aG9yPkNob2ksIFNldW5nIEhvYW48L2F1dGhvcj48YXV0aG9yPldlbmcsIEx1LUNoZW48L2F1dGhvcj48YXV0aG9yPktveWFtYSwgU2F0b3NoaTwvYXV0aG9yPjxhdXRob3I%2BUGlycnVjY2VsbG8sIEphbWVzIFAuPC9hdXRob3I%2BPGF1dGhvcj5OZ3V5ZW4sIFRyYW5nPC9hdXRob3I%2BPGF1dGhvcj5TbWFkYmVjaywgUGF0cmljazwvYXV0aG9yPjxhdXRob3I%2BSmFuZywgRG9uZ2tldW48L2F1dGhvcj48YXV0aG9yPkNoYWZmaW4sIE1hcms8L2F1dGhvcj48YXV0aG9yPldhbHNoLCBSb2RkeTwvYXV0aG9yPjxhdXRob3I%2BUm9zZWxsaSwgQ2Fyb2xpbmE8L2F1dGhvcj48YXV0aG9yPkVsbGlvdHQsIEFtYW5kYSBMLjwvYXV0aG9yPjxhdXRob3I%2BV2lqZGV2ZWxkLCBMZW9ub29yIEYuIEouIE0uPC9hdXRob3I%2BPGF1dGhvcj5CaWRkaW5nZXIsIEtpcmFuIEouPC9hdXRob3I%2BPGF1dGhvcj5LYW55LCBTaGlud2FuPC9hdXRob3I%2BPGF1dGhvcj5Sw6Rtw7YsIEpvZWwgVC48L2F1dGhvcj48YXV0aG9yPk5hdGFyYWphbiwgUHJhZGVlcDwvYXV0aG9yPjxhdXRob3I%2BQXJhZ2FtLCBLcmlzaG5hIEcuPC9hdXRob3I%2BPGF1dGhvcj5GbGFubmljaywgSmFzb248L2F1dGhvcj48YXV0aG9yPkJ1cnR0LCBOb8OrbCBQLjwvYXV0aG9yPjxhdXRob3I%2BQmV6emluYSwgQ29ubmllIFIuPC9hdXRob3I%2BPGF1dGhvcj5MdWJpdHosIFN0ZXZlbiBBLjwvYXV0aG9yPjxhdXRob3I%2BTHVuZXR0YSwgS2F0aHJ5biBMLjwvYXV0aG9yPjxhdXRob3I%2BRWxsaW5vciwgUGF0cmljayBULjwvYXV0aG9yPjwvYXV0aG9ycz48). We excluded any phecode with < 100 cases.

#### Supplementary Methods 11. Definition of novel gene-phenotype associations in RVAT

We downloaded all rare variant analysis test results from the Open Targets Platform and GeneBass. Also, we downloaded the summary statistics from the whole-exome imputation within the UKB study[^25^](https://web.endnote.com/citations/eyJkaXNwbGF5VGV4dCI6IjI1IiwiY2l0YXRpb25zIjpbeyJiaWJsaW9Db250ZW50IjpbeyJlbGVjdHJvbmljUmVzb3VyY2VOdW1iZXIiOiIxMC4xMDM4L3M0MTU4OC0wMjEtMDA4OTItMSIsImdyb3VwR3VpZHMiOltdLCJyZWNvcmRTdGF0dXMiOiJhY3RpdmUiLCJzZWNvbmRhcnlUaXRsZSI6Ik5hdHVyZSBHZW5ldGljcyIsImF1dGhvcnMiOlsiQmFydG9uLCBBbGlzb24gUi4iLCJTaGVybWFuLCBNYXh3ZWxsIEEuIiwiTXVrYW1lbCwgUm9uZW4gRS4iLCJMb2gsIFBvLVJ1Il0sImlzYm4iOiIxMDYxLTQwMzYiLCJ2b2x1bWUiOiI1MyIsInJzeG1sIjoiPHJlY29yZD48cmVmLXR5cGU%2BMTc8L3JlZi10eXBlPjxjb250cmlidXRvcnM%2BPGF1dGhvcnM%2BPGF1dGhvcj5CYXJ0b24sIEFsaXNvbiBSLjwvYXV0aG9yPjxhdXRob3I%2BU2hlcm1hbiwgTWF4d2VsbCBBLjwvYXV0aG9yPjxhdXRob3I%2BTXVrYW1lbCwgUm9uZW4gRS48L2F1dGhvcj48YXV0aG9yPkxvaCwgUG8tUnU8L2F1dGhvcj48L2F1dGhvcnM%2BPC9jb250cmlidXRvcnM%2BPHRpdGxlcz48dGl0bGU%2BV2hvbGUtZXhvbWUgaW1wdXRhdGlvbiB3aXRoaW4gVUsgQmlvYmFuayBwb3dlcnMgcmFyZSBjb2RpbmcgdmFyaWFudCBhc3NvY2lhdGlvbiBhbmQgZmluZS1tYXBwaW5nIGFuYWx5c2VzPC90aXRsZT48c2Vjb25kYXJ5LXRpdGxlPk5hdHVyZSBHZW5ldGljczwvc2Vjb25kYXJ5LXRpdGxlPjwvdGl0bGVzPjxkYXRlcz48eWVhcj4yMDIxPC95ZWFyPjwvZGF0ZXM%2BPHBhZ2VzPjEyNjAtMTI2OTwvcGFnZXM%2BPHZvbHVtZT41Mzwvdm9sdW1lPjxpc2JuPjEwNjEtNDAzNjwvaXNibj48ZWxlY3Ryb25pYy1yZXNvdXJjZS1udW0%2BPHN0eWxlIGZhY2U9XCJ1bmRlcmxpbmVcIj4xMC4xMDM4L3M0MTU4OC0wMjEtMDA4OTItMTwvc3R5bGU%2BPC9lbGVjdHJvbmljLXJlc291cmNlLW51bT48bnVtYmVyPjg8L251bWJlcj48cmVjLWd1aWQ%2BMWI5ODRjNjMtZDJkZi00MTJmLWIzYjEtYjlmYWVkNmI0MjVjPC9yZWMtZ3VpZD48cmVjLXVzbj40ODwvcmVjLXVzbj48L3JlY29yZD4iLCJwYWdlcyI6IjEyNjAtMTI2OSIsInJlZmVyZW5jZVR5cGUiOiIxNyIsIm51bWJlciI6IjgiLCJ5ZWFyIjoiMjAyMSIsInRpdGxlIjoiV2hvbGUtZXhvbWUgaW1wdXRhdGlvbiB3aXRoaW4gVUsgQmlvYmFuayBwb3dlcnMgcmFyZSBjb2RpbmcgdmFyaWFudCBhc3NvY2lhdGlvbiBhbmQgZmluZS1tYXBwaW5nIGFuYWx5c2VzIiwiZ3VpZCI6IjFiOTg0YzYzLWQyZGYtNDEyZi1iM2IxLWI5ZmFlZDZiNDI1YyJ9XSwicmVjb3JkIjp7ImNvbnRyaWJ1dG9ycyI6eyJhdXRob3JzIjp7ImF1dGhvciI6WyJCYXJ0b24sIEFsaXNvbiBSLiIsIlNoZXJtYW4sIE1heHdlbGwgQS4iLCJNdWthbWVsLCBSb25lbiBFLiIsIkxvaCwgUG8tUnUiXX19LCJwYWdlcyI6IjEyNjAtMTI2OSIsInZvbHVtZSI6IjUzIiwiaXNibiI6IjEwNjEtNDAzNiIsImVsZWN0cm9uaWMtcmVzb3VyY2UtbnVtIjp7InN0eWxlIjp7IiMiOiIxMC4xMDM4L3M0MTU4OC0wMjEtMDA4OTItMSIsIkBmYWNlIjoidW5kZXJs). We defined novel gene-phenotype pairs when it has not been reported in these dataset. All URLs are shown in the **Supplementary Data 46**.

### **Supplementary Figures**

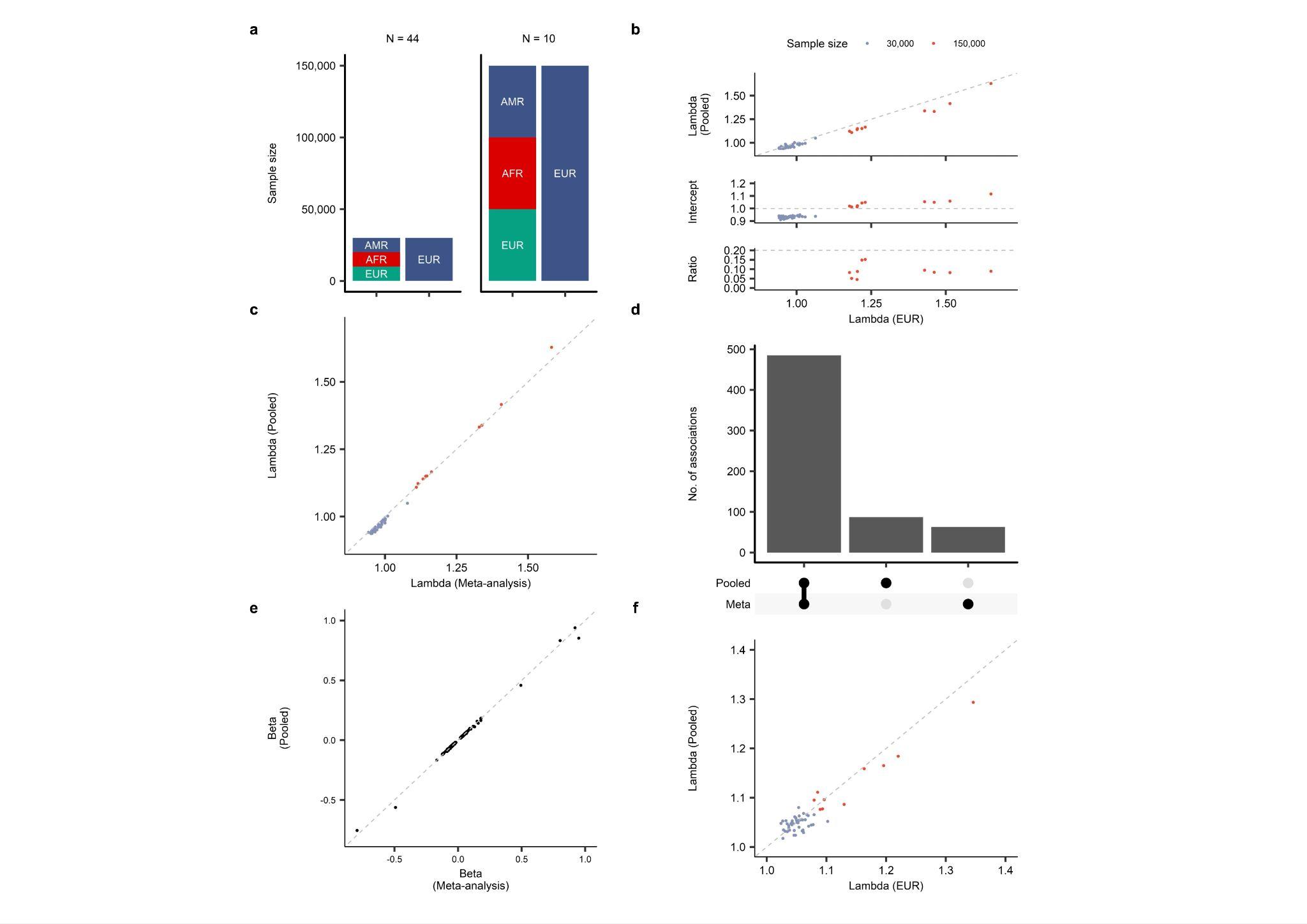

#### Supplementary Fig.1: Comparison of GWAS and RVAS between single-ancestry and pooled multi-ancestry frameworks.

**a,** Schematic overview of the study design evaluating single-ancestry versus pooled multi-ancestry approaches. Simulations were conducted across two configurations: a 100% EUR cohort and a pooled multi-ancestry cohort (comprising an equal 33%:33%:33% split of AFR, AMR, and EUR populations), each evaluated at sample sizes of 30,000 individuals (for 44 phenotypes) and 150,000 individuals (for 10 phenotypes). **b,** Comparison of λ_GC_ (EUR), LDSC intercept (EUR), the attenuation ratio (EUR), and λ_GC_ (pooled). The dashed lines indicate y = x (λ_GC_ [pooled]), y = 1.0 (LDSC intercept), and y = 0.2 (attenuation ratio), respectively. **c,** Comparison of λ_GC_ derived from meta-analysis with that from pooled analysis. The color scheme is the same as in **b**. **d,** The number of genome-wide significant loci in the meta-analysis and the pooled analysis. **e,** Comparison of P values between the meta-analysis and the pooled analysis. **f,** Comparison of λ_GC_ (95 percentile) between the European RVAS and the pooled RVAS. The color scheme is the same as in **b**. AFR, African-American; AMR, admixed American; EUR, European; GWAS, genome-wide association study; LDSC, linkage disequilibrium score regression; RVAS, Rare Variant Association Studies.

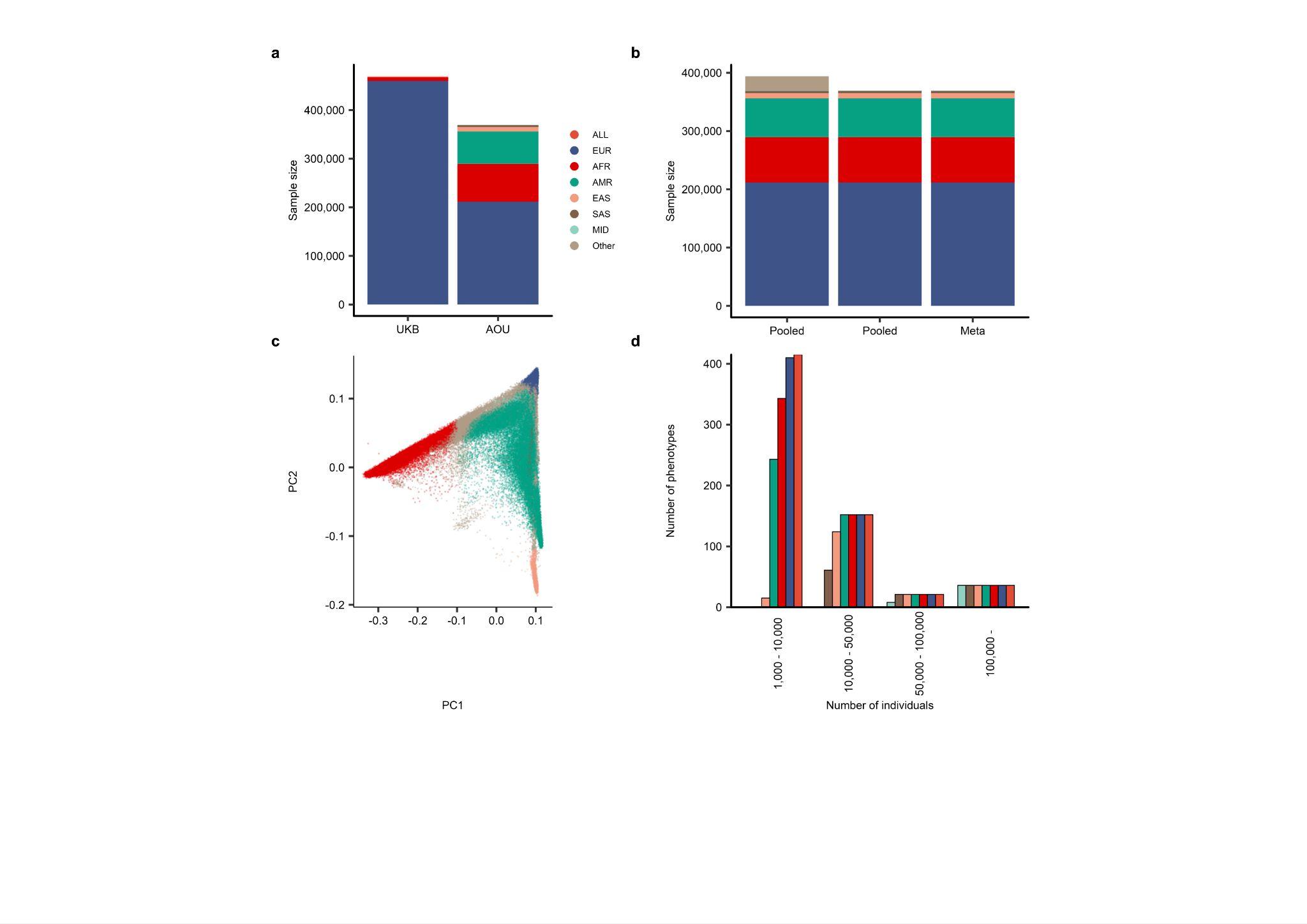

#### Supplementary Fig.2: The characteristics of AoU.

**a,** Comparison of sample sizes between AoU and UKB. The same color legend is used in **b**, **c**, and **d**. **b,** The principal component analysis (PCA) in AoU. **c,** The schematic representation of two pooled analyses and the meta-analysis. **d,** The sample size of each phenotype. AFR, African American; AMR, Admixed American; AoU, All of Us; EAS, East Asian; EUR, European; MID, Middle Eastern; PC, principal component; SAS, South Asian; UKB, UK Biobank.

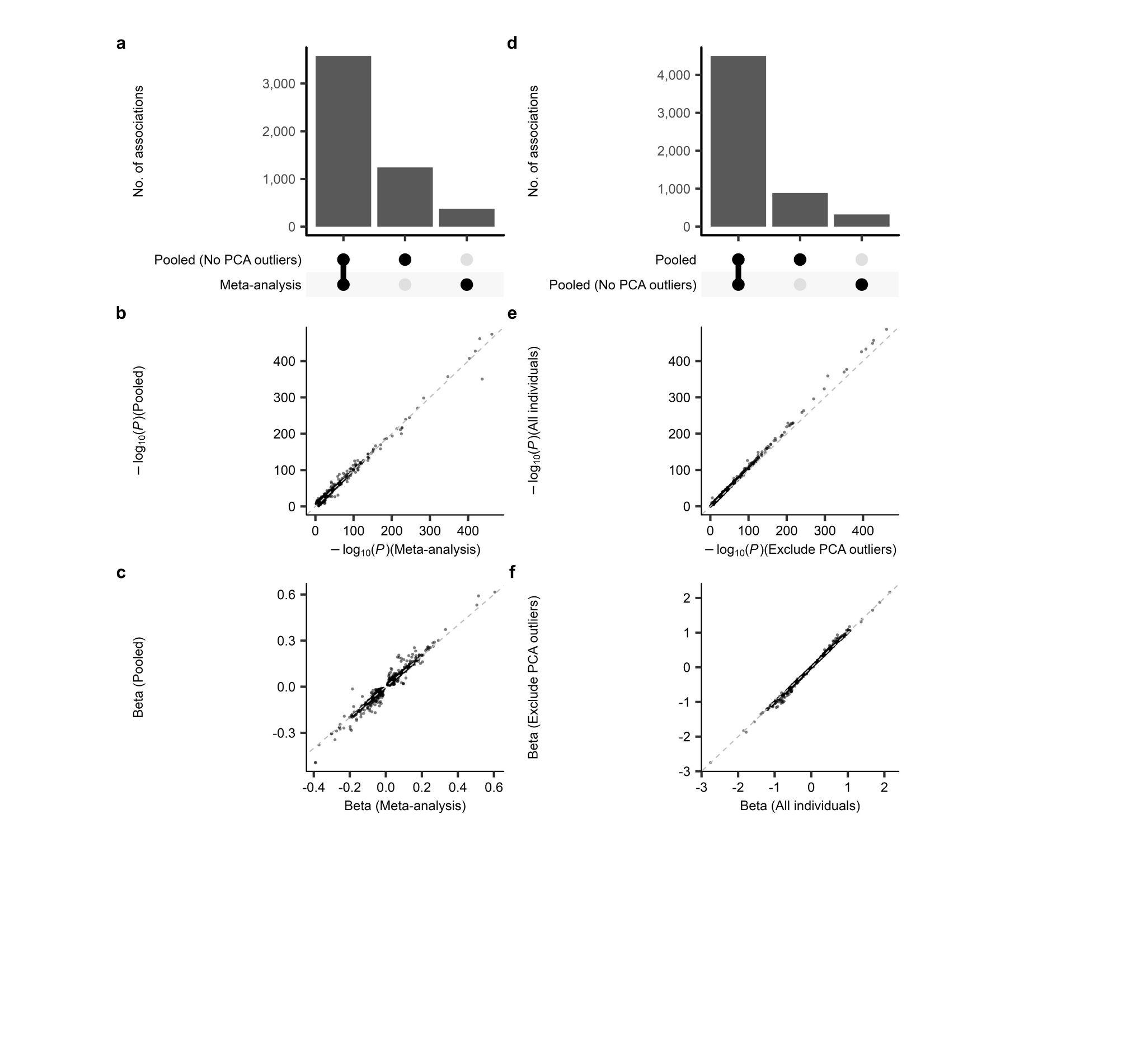

#### Supplementary Fig.3: Comparison of the pooled analysis with meta-analysis.

**a,** Number of genome-wide significant loci in the pooled analysis excluding PCA outliers and the meta-analysis. **b, c**, Comparison of P values (b) and beta (c) between the pooled analysis and the meta-analysis. **d,** The number of genome-wide significant loci in the pooled analysis with or without PCA outliers. **e, f,** Comparison of P values (e) and beta (f) between the pooled analysis with and without PCA outliers. PCA, principal component analysis.

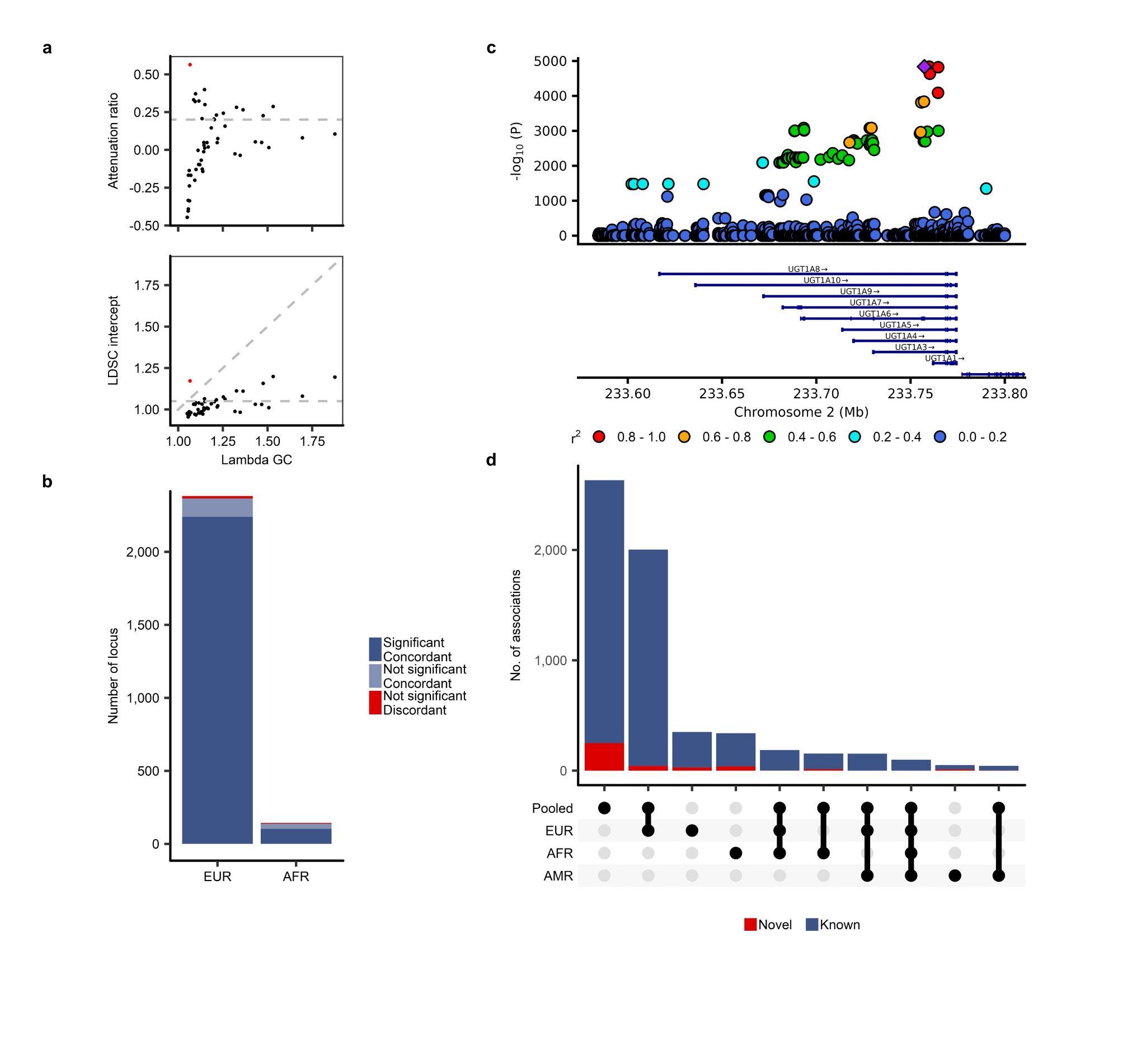

#### Supplementary Fig.4: GWAS results for 624 phenotypes.

**a,** The λ_GC_, LDSC intercept attenuation ratio calculated from 1,982 ancestry-phenotype pairs (624 phenotypes). The ancestry-phenotype pairs with λ_GC_ > 1.05 are shown. **b**, Replication analysis using UK Biobank for loci reaching genome-wide significance in AoU, with analyses performed separately in EUR and AFR ancestry groups. **c**, Locus plots for loci with P × 10-^4000^. **d**, Number of loci reaching significance in each ancestry group. Only ancestry groups with ≥ 10 loci were included in the plot. AFR, African, AMR, admixed-American, AoU, All of Us; EUR, European; GWAS, genome-wide association study; LDSC, linkage disequilibrium score regression.

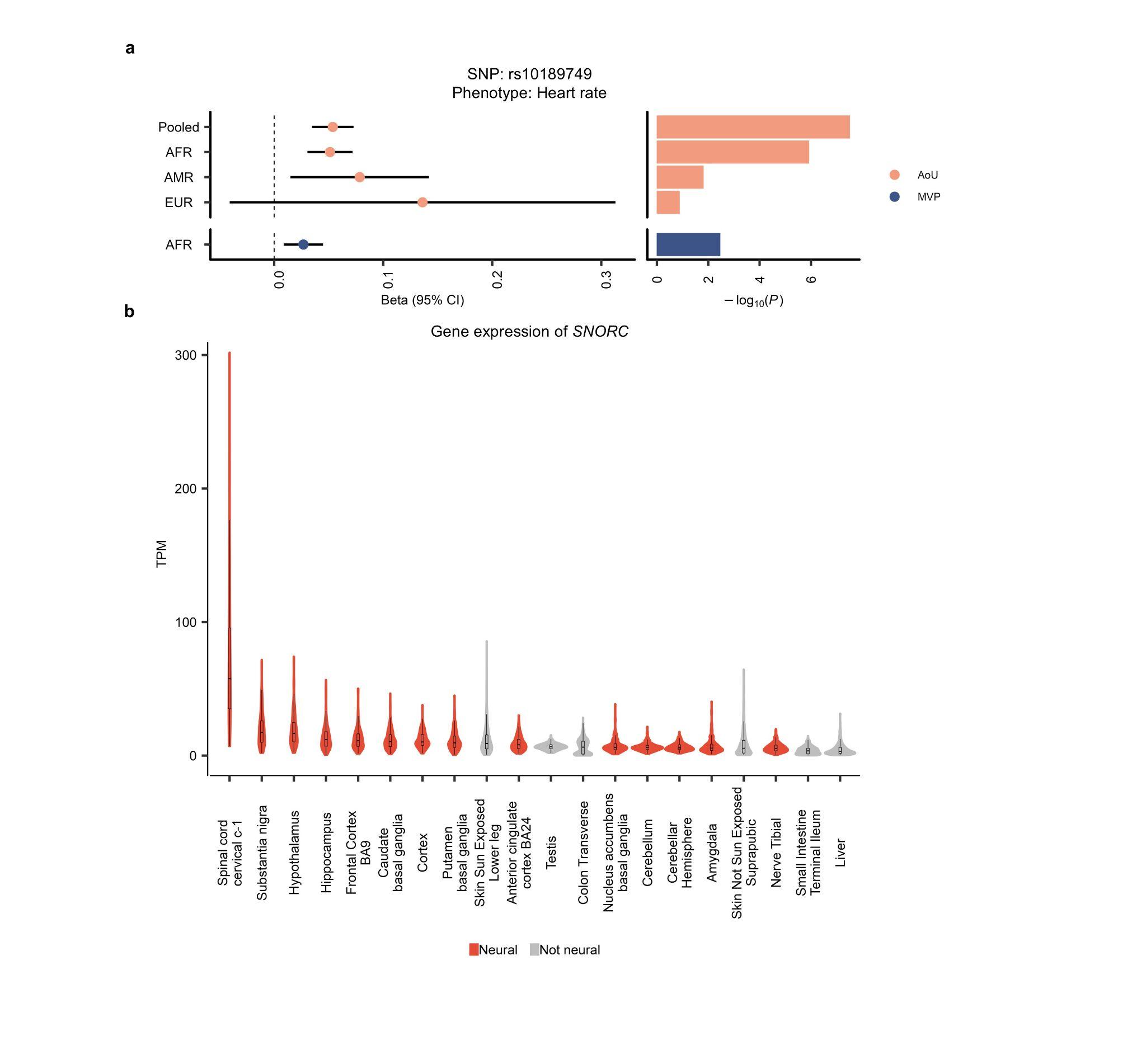

#### Supplementary Fig.5: The follow-up analysis of rs10189749.

**a,** Effect estimates and P values of rs10189749 in each ancestry among AoU and MVP. The error bar for the ORs indicates the 95% CIs. **b,** Top 20 expressions of *SNORC* in each tissue. The vertical axis indicates TPM in each tissue. AFR, African American; AMR, Admixed American; AoU, All of Us; CI, confidence interval; EUR, European; MVP, Million Veterans Program; OR, odds ratio; TPM, Transcripts Per Million.

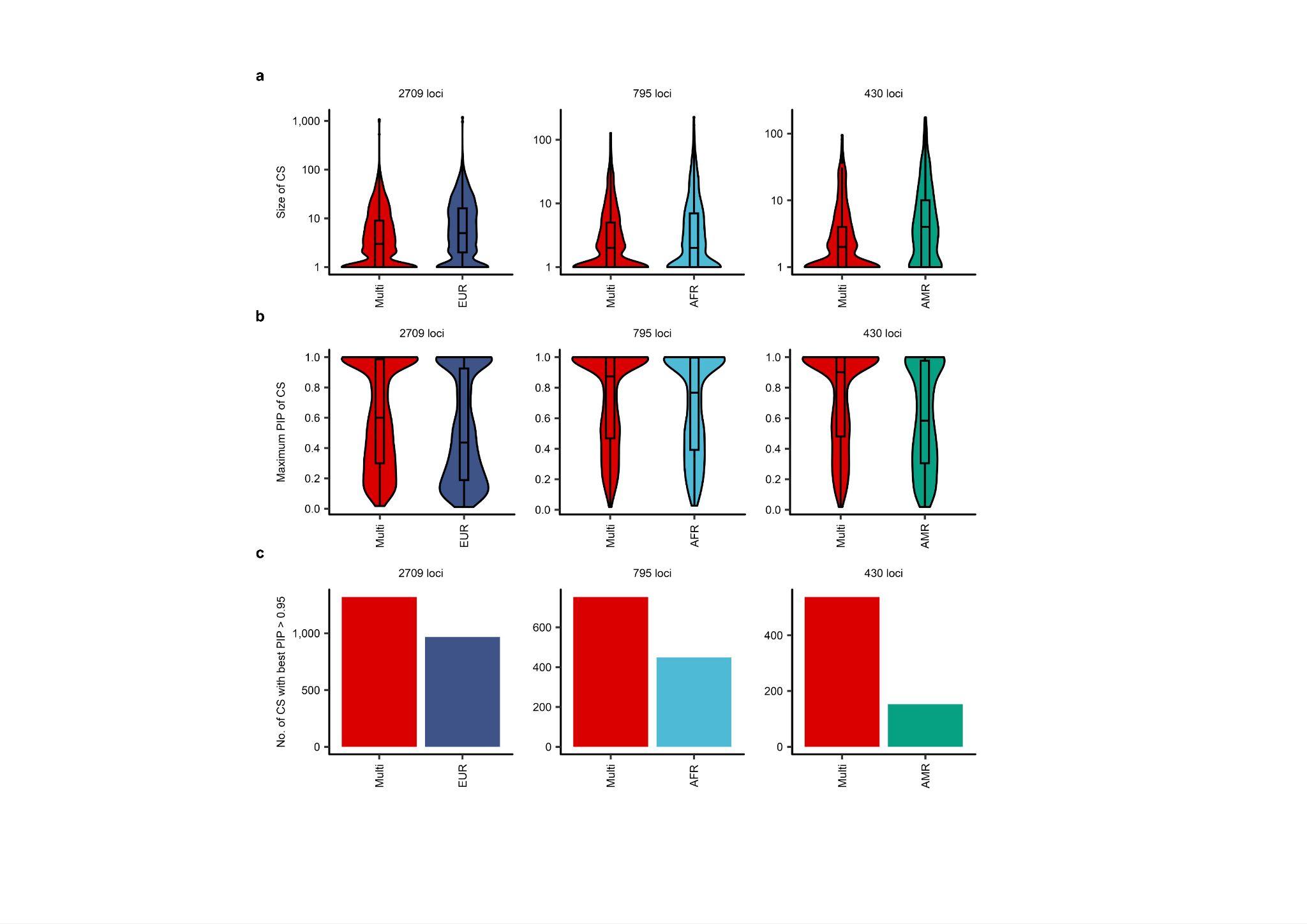

#### Supplementary Fig.6: Performance comparison between multi-ancestry and single-ancestry fine-mapping.

**a,** Comparison of size of CS between multi-ancestry and single-ancestry fine-mapping. The size of CS is defined as the number of variants included in the CS. The box plot shows the median value as the centerline; the box boundaries show the first and third quartiles and the whiskers extend 1.5 times the interquartile range in **a** and **b**. **b,** Comparison of maximum PIP of CS. **c,** The comparison of the number of CS with best PIP > 0.95. Effect estimate. AFR, African; AMR, Admixed American; CS, credible set; EUR, European; Multi, multi-ancestry; PIP, posterior inclusion probability.

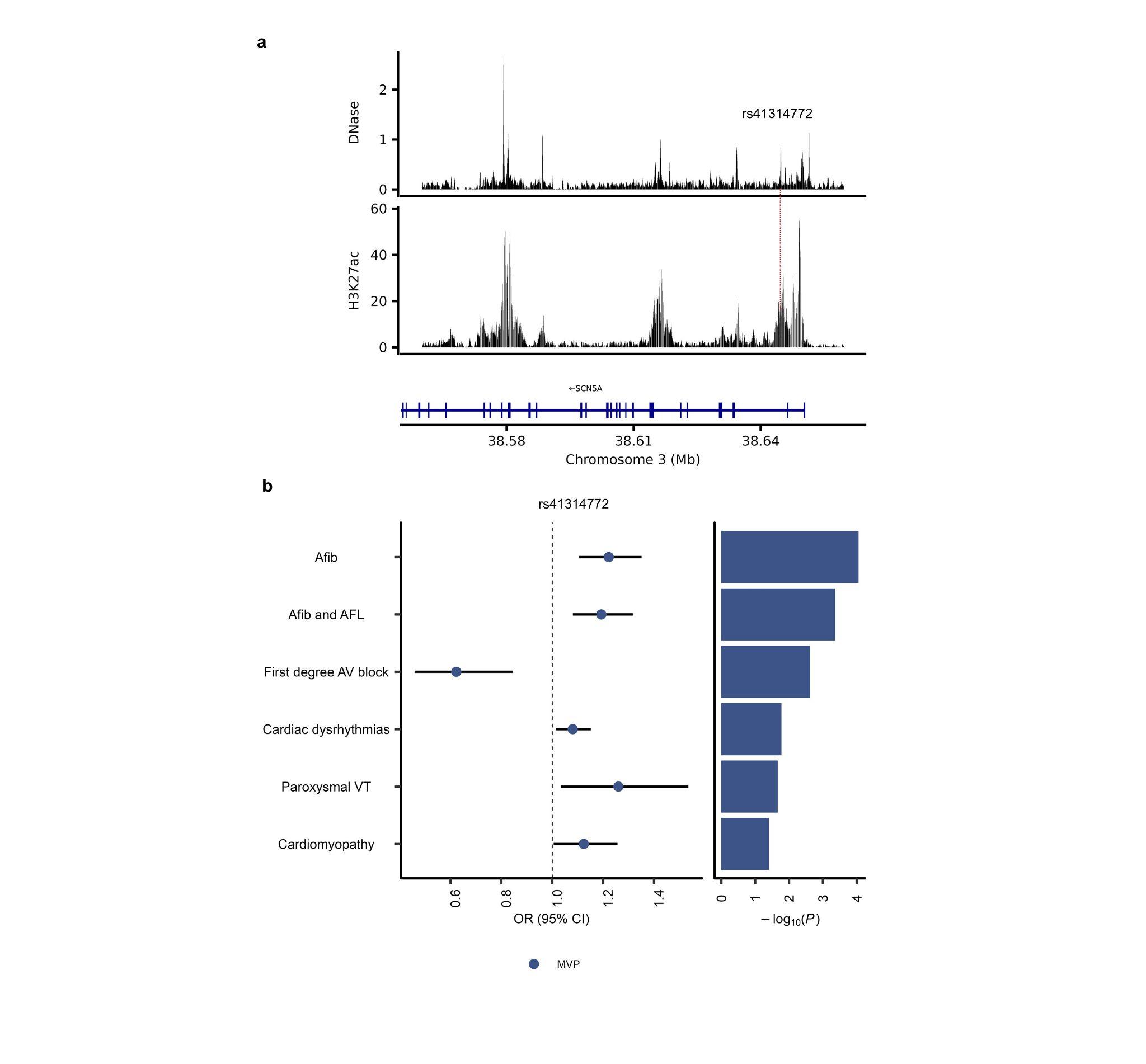

#### Supplementary Fig.7: The follow-up analysis of rs41314772.

**a,** DNase peak and H3K27ac peaks for the *SCN5A* locus. The horizontal axis indicates the genomic coordinates. The red line indicates the genomic location of rs41314772. DNase (ENCSR299QGI) and H3K27ac (ENCSR932QRC) peaks are based on the left ventricle (**Supplementary Data 14**). **b,** ORs and P values of rs41314772 in MVP. The error bar for the effect estimates indicates the 95% CIs. Afib, Atrial fibrillation; AFL, Atrial flutter; AV, atrioventricular; CI, confidence interval; DNase, Deoxyribonuclease; H3K27ac, Histone H3 Lysine 27 acetylation; MVP, Million Veterans Program; OR, odds ratio; VT, ventricular tachycardia.

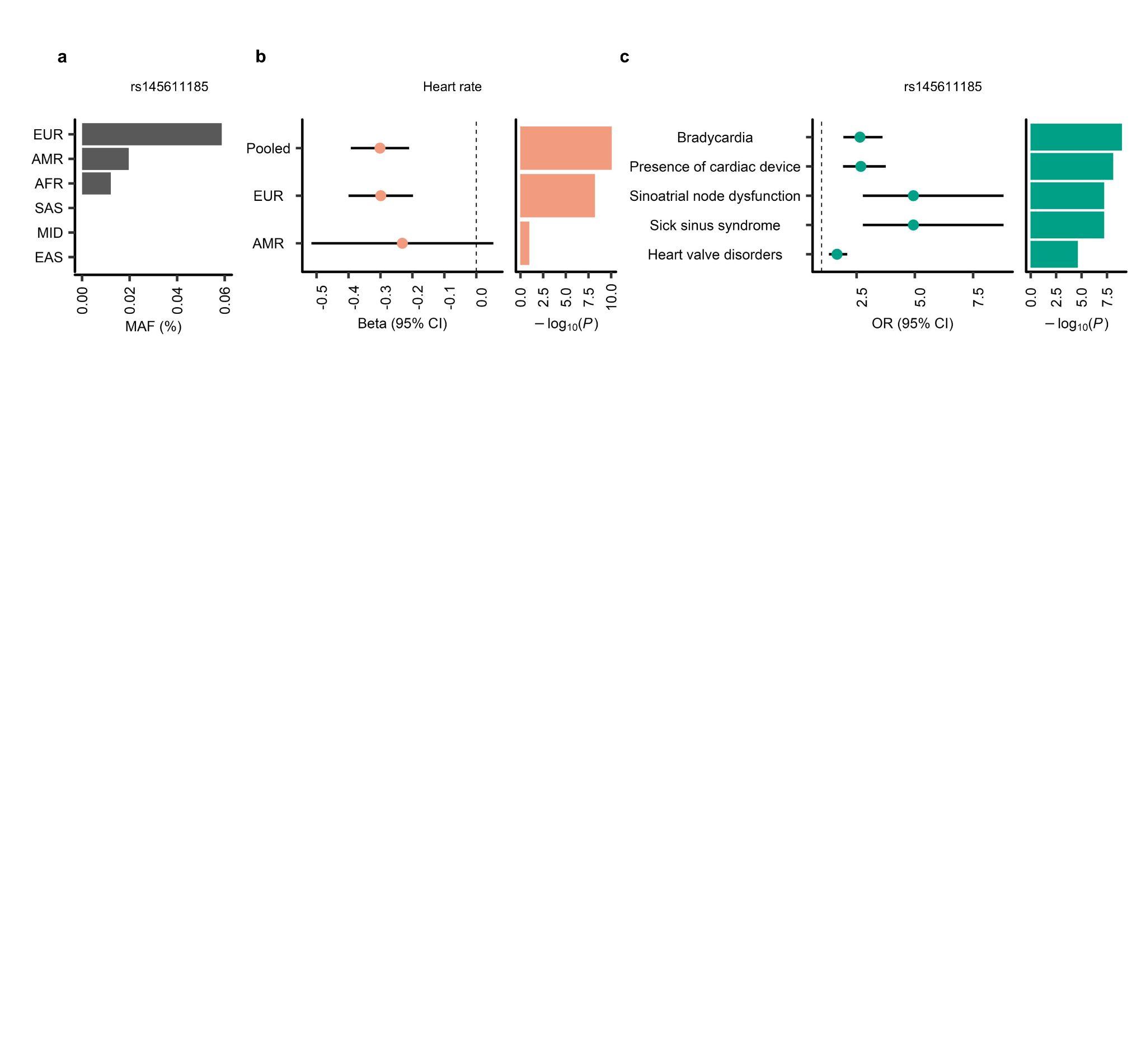

#### Supplementary Fig.8: The impact of rs145611185 on quantitative and binary traits in AoU and UKB.

**a,** The MAF of rs145611185 in each ancestry, derived from gnomAD v4.1.1. **b,** Effect estimates and P values of rs145611185 for heart rate in each ancestry among AoU. The error bar for the effect estimates indicates the 95% CIs. **c,** ORs and P values of rs145611185 for cardiovascular phenotypes in each UKB. The error bar for the ORs indicates the 95% CIs. AoU, AFR, African American; AMR, Admixed American; AoU, All of Us; CI, confidence interval; EAS, East Asian; EUR, European; MID, Middle Eastern; OR, odds ratio; SAS, South Asian; UKB, UK Biobank.

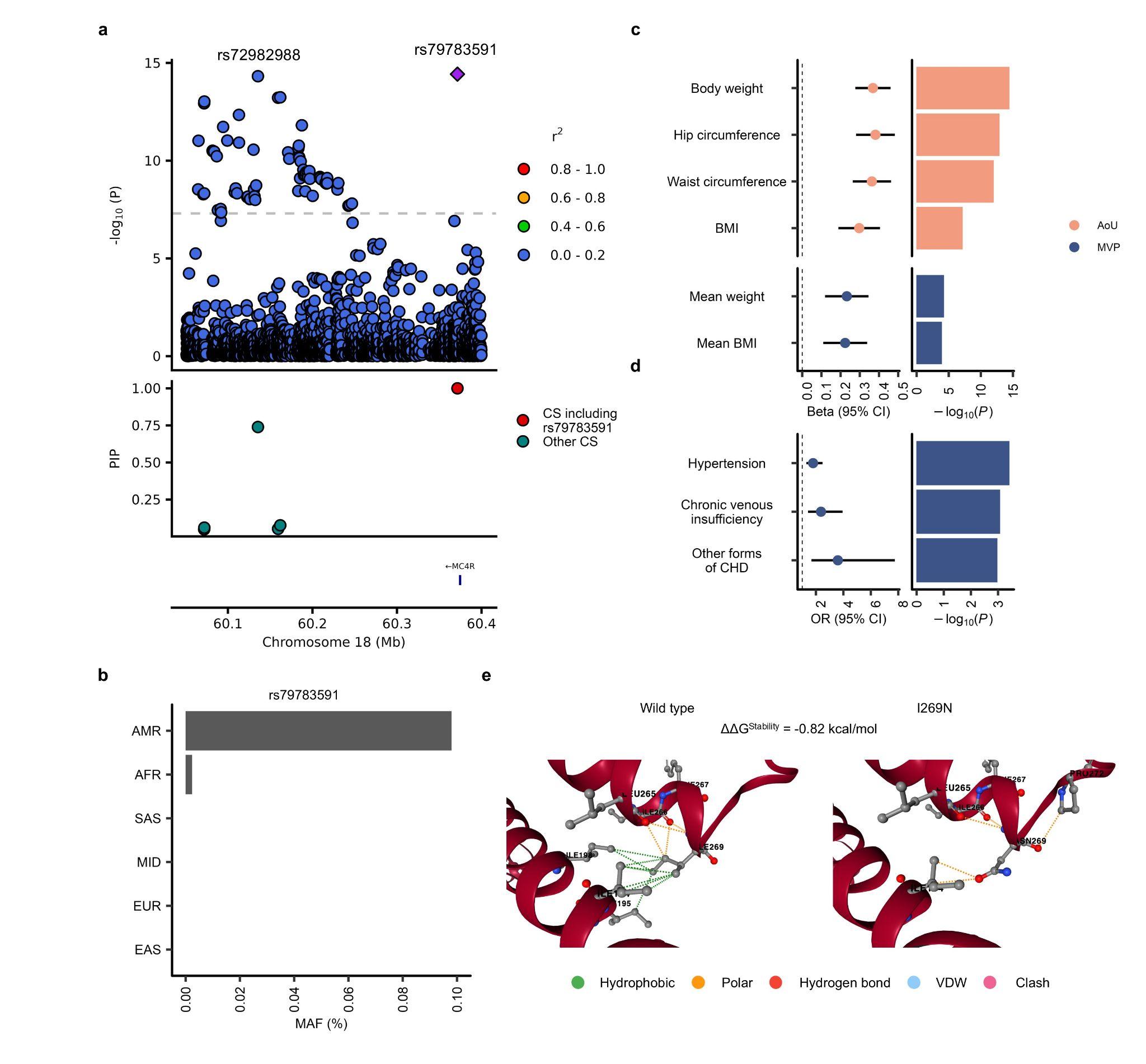

#### Supplementary Fig.9: The admixed-American specific missense variant in *MC4R* and obesity.

**a,** The locus plot and PIP for the *MC4R* locus. The horizontal axis indicates the genomic coordinates. **b,** The MAF of rs79783591 in each ancestry, derived from gnomAD v4.1.1. **c,** Effect estimates and P values of rs79783591 for obesity-related phenotypes in each ancestry among AoU and MVP. The error bar for the effect estimates indicates the 95% CI. **d,** ORs and P values of rs79783591 on binary phenotypes in MVP. The error bar for the ORs indicates the 95% CI. **e,** Results from DynaMut2. The negative ΔΔG^Stability^ indicates that the missense variant causes structural instability. AFR, African American; AMR, Admixed American; AoU, All of Us; BMI, body mass index; CHD, chronic heart disease; CI, confidence interval; CS, credible set; EAS, East Asian; EUR, European; MAF, minor allele frequency; MID, Middle Eastern; MVP, Million Veteran Program; OR, odds ratio; PIP, posterior inclusion probability; SAS, South Asian; VDW, van der Waals.

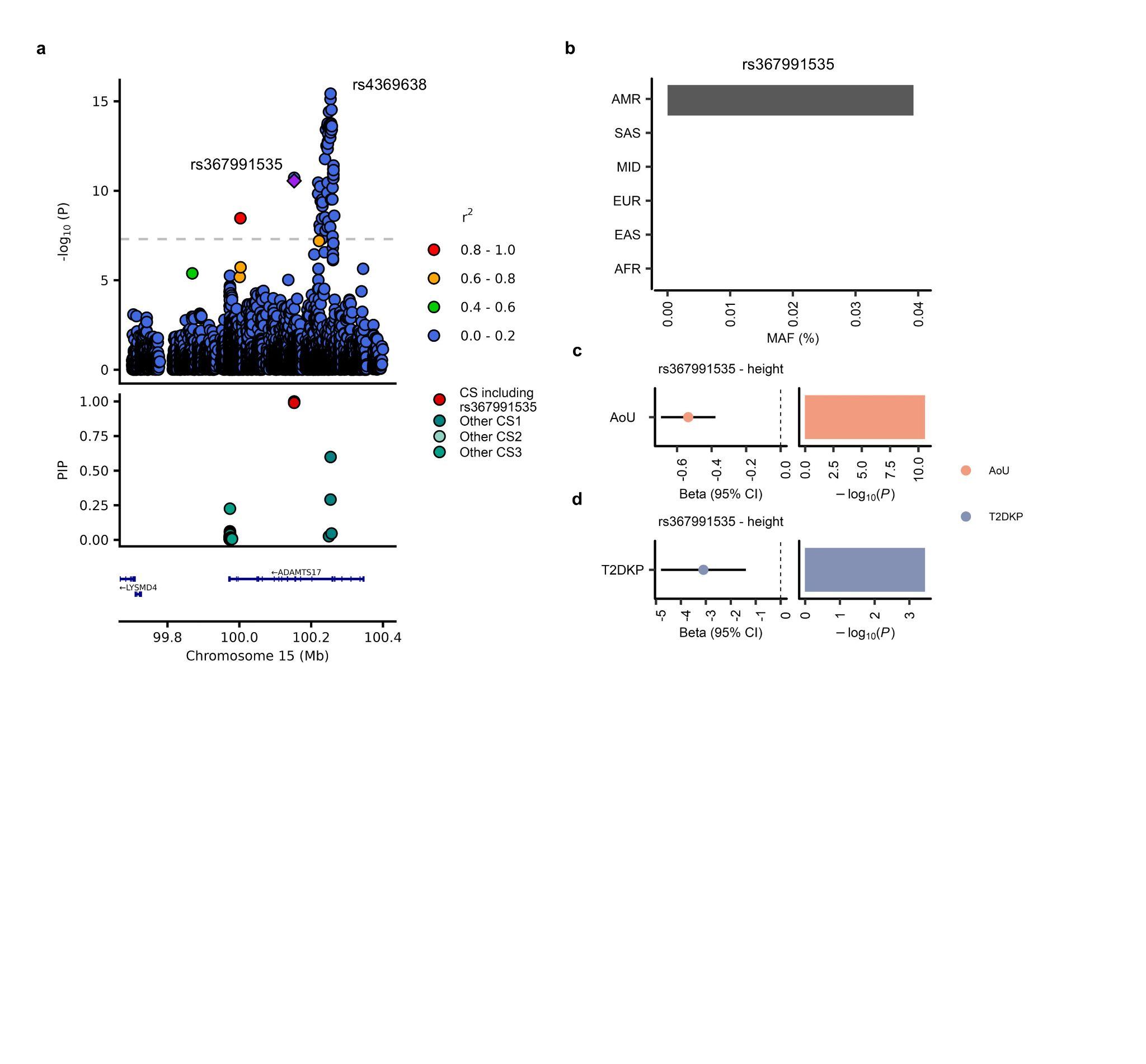

#### Supplementary Fig.10: The admixed-American specific missense variant in *ADAMTS17* and body height.

**a,** The locus plot and PIP for the *ADAMTS17* locus. The horizontal axis indicates the genomic coordinates. **b,** The MAF of rs367991535 in each ancestry, derived from gnomAD v4.1.1. **c, d,** Effect estimates and P values of rs367991535 for body height in AoU (**c**) and in T2D knowledge portal (**d**). Of note, phenotype data were inverse-rank normalized in AoU while not in the T2D knowledge portal, which explains the difference in beta. The error bar for the effect estimates indicates the 95% CI. AFR, African American; AMR, Admixed American; AoU, All of Us; CI, confidence interval; CS, credible set; EAS, East Asian; EUR, European; MAF, minor allele frequency; MID, Middle Eastern; PIP, posterior inclusion probability; SAS, South Asian; T2DKP, Type 2 Diabetes Knowledge Portal.

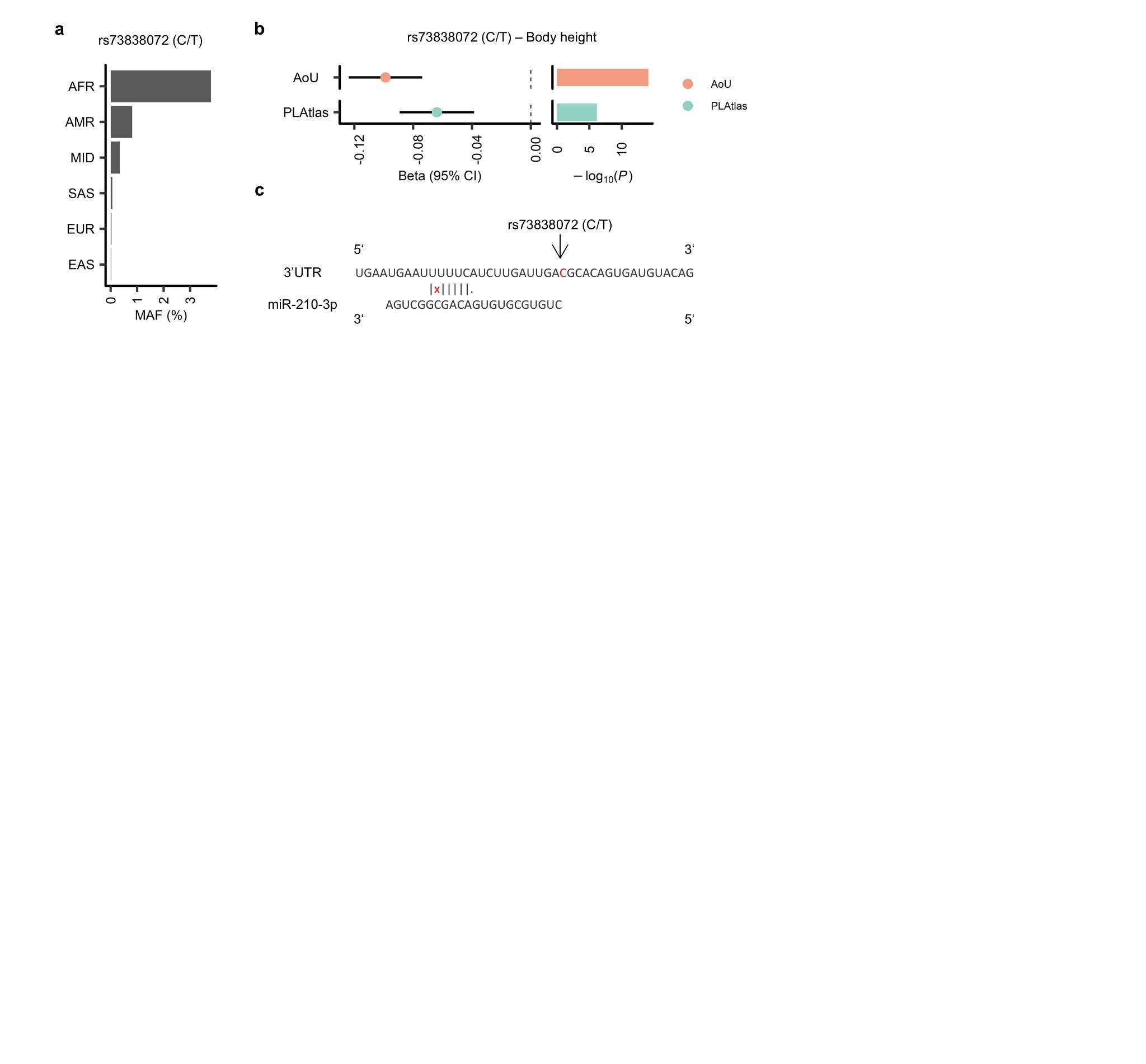

#### Supplementary Fig.11: rs73838072 affects the binding site of miR-210-3p.

**a,** The MAF of rs73838072 in each ancestry, derived from gnomAD v4.1.1. **b,** Effect estimates and P values of rs73838072 for body height in AFR among AoU and MVP. The errors bar for the effect estimates indicates the 95% CIs. **c,** Binding site of miR-210-3p. “|” indicates matching, “X” mismatching, and “.” wobble matching. AFR, African American; AMR, Admixed American; AoU, All of Us; CI, confidence interval; EAS, East Asian; EUR, European; MAF, minor allele frequency; MID, Middle Eastern; SAS, South Asian; UTR, untranslated region.

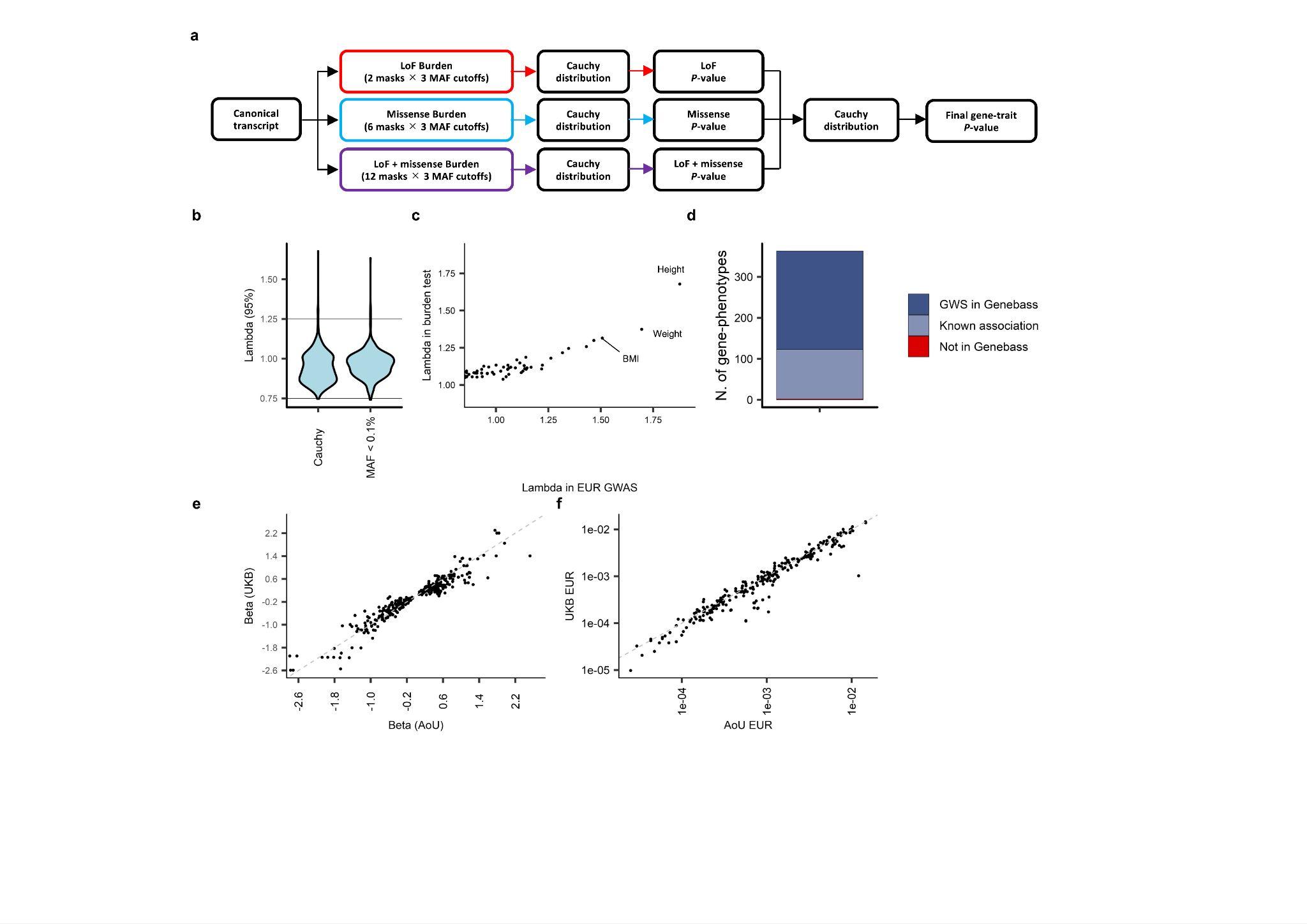

#### Supplementary Fig.12: Analytical pipeline and quality control for RVAS.

**a,** Analytical scheme to produce a single test statistic for a given gene-phenotype pair. **b,** λ_95%_ derived from 624 phenotypes using the final gene-trait P value. We repeated the same analysis using masks with MAF < 0.1%. **c,** The association between λ_95%,burden_ and λ_GC,GWAS_. We annotated top 3 phenotypes with inflated λ_95%,burden_. **d,** Replication analysis using Open Targets Platform and GeneBass. **e,** Comparison of effect estimates between AoU and UKB. **f,** Comparison of cumulative MAF between AoU EUR and UKB EUR. AoU, All of Us; GWS, genome-wide significant; LoF, Loss-of-Function; MAF, minor allele frequency; RVAS, Rare Variant Association Studies; UKB, UK Biobank.

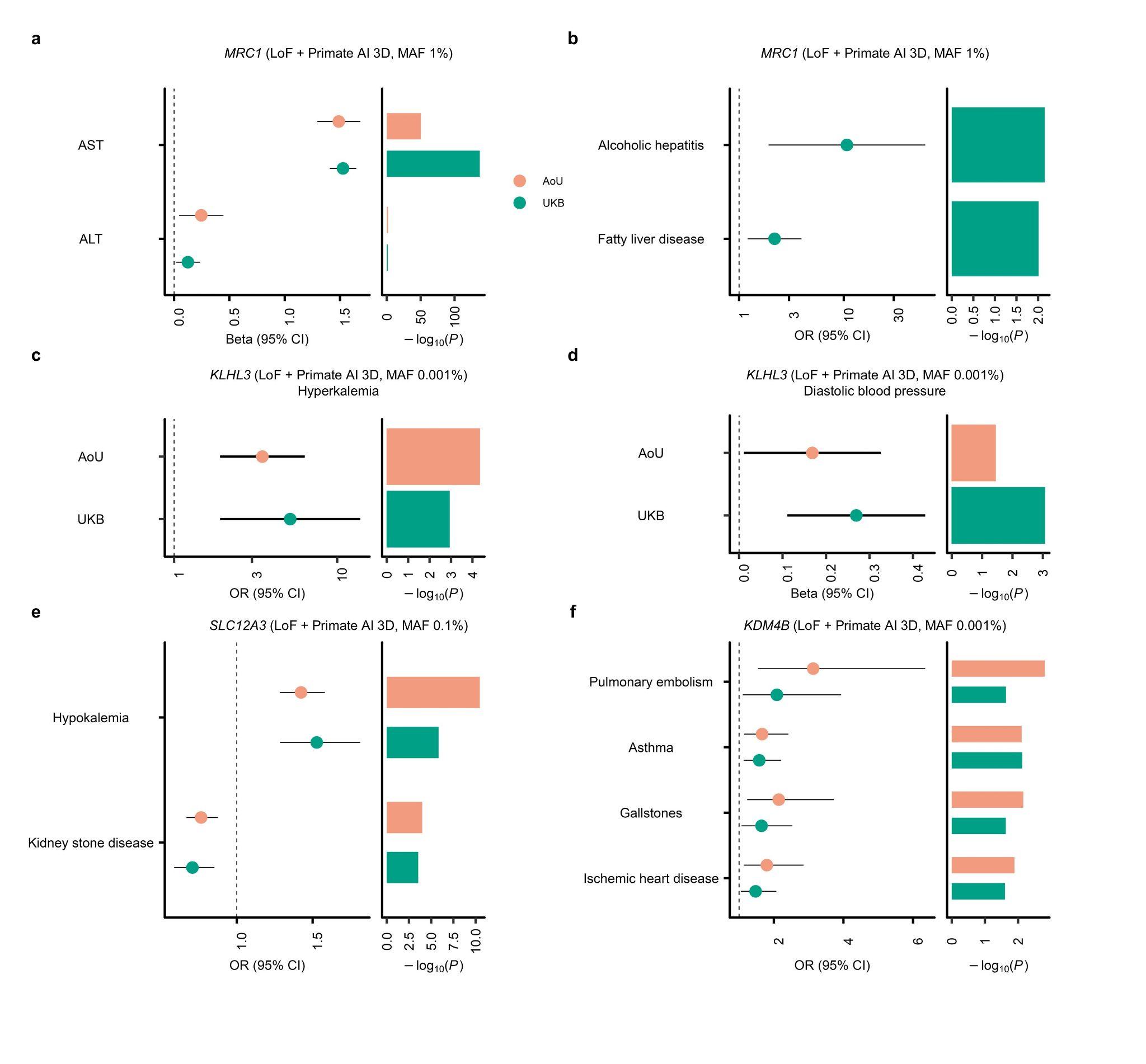

#### Supplementary Fig.13: Novel gene-phenotype associations.

**a,** Effect estimates and P values of *MRC1* with the best mask for liver function-related quantitative traits among AoU and UKB. The error bar for the effect estimates indicates the 95% CIs in all figures. **b,** Effect estimates and P values of *MRC1* with the best mask for liver function-related binary traits among UKB. **c, d,** Effect estimates and P values of *KLHL3* with the best mask for hyperkalemia (**c**) and diastolic blood pressure (**d**) among AoU and UKB. **e,** ORs and P values of *SLC12A3* with the best mask for hypokalemia and kidney stone disease among AoU and UKB. **f,** ORs and P values of *KDM4B* with the best mask for obesity-related phenotypes among AoU and UKB. The same color scheme is used in all the figures. ALT, Alanine aminotransferase; AST, Aspartate aminotransferase; AoU, All of Us; CI, confidence interval; LoF, Loss-of-Function; MAF, minor allele frequency; OR, odds ratio; UKB, UK Biobank.

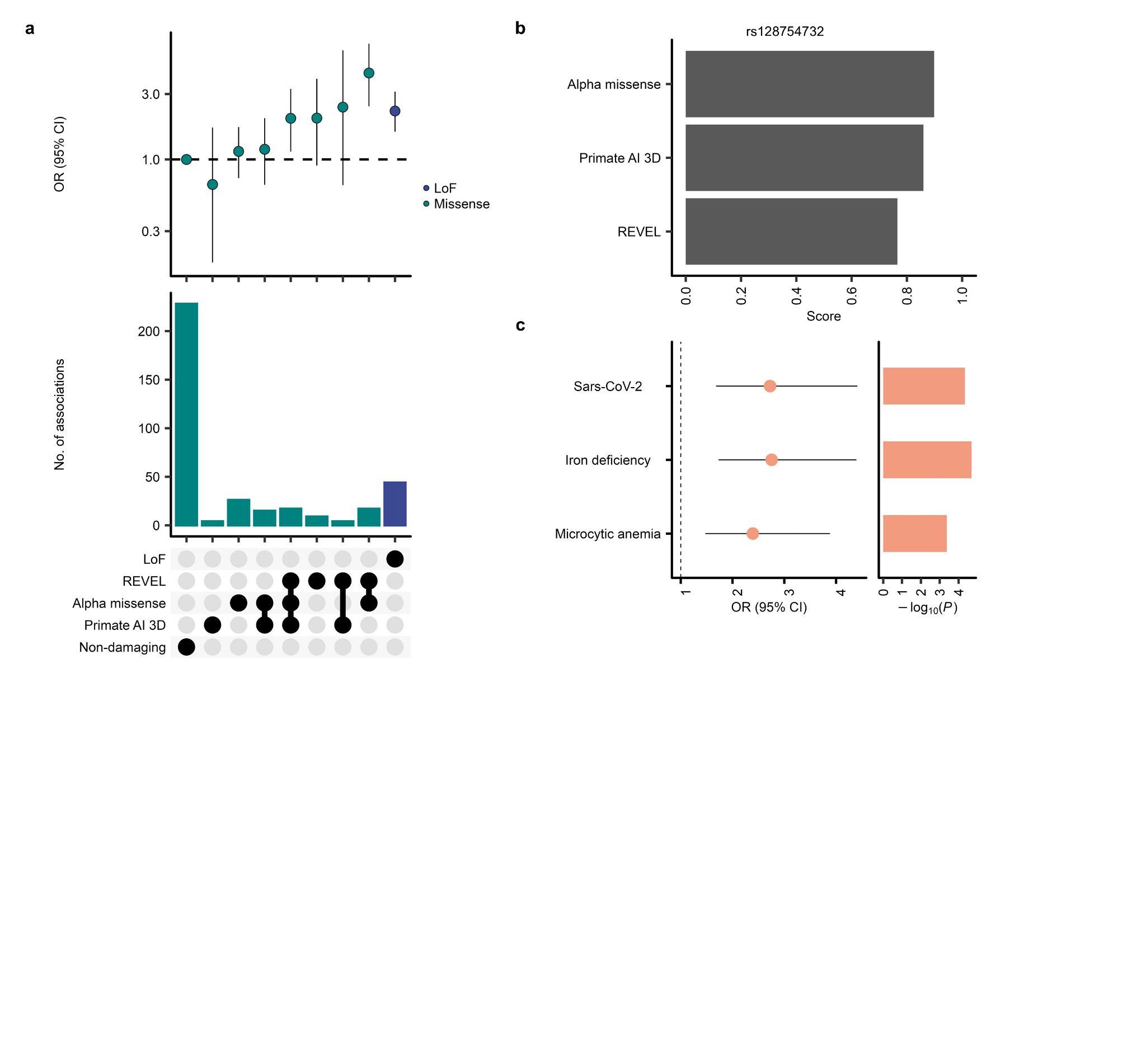

#### Supplementary Fig.14: Using multiple missense predictors identifies candidate causal variants.

**a,** Enrichment of LoF and damaging missense variants among significant variants with MAF < 1% in exon regions. The top panel shows the enrichment of indicated variant classes in the significant variants compared to non-damaging missense variants. The bottom panel shows the number of significant variants in the indicated variant classes. The error bar for the effect estimates indicates the 95% CIs in **a** and **c**. **b,** Results from the missense predictors for rs128754732. **c,** ORs and P values of rs1287547322 for binary traits in the pooled analysis. CI, confidence interval; LoF, Loss-of-Function; OR, odds ratio; Sars-Cov-2, Severe Acute Respiratory Syndrome Coronavirus 2.

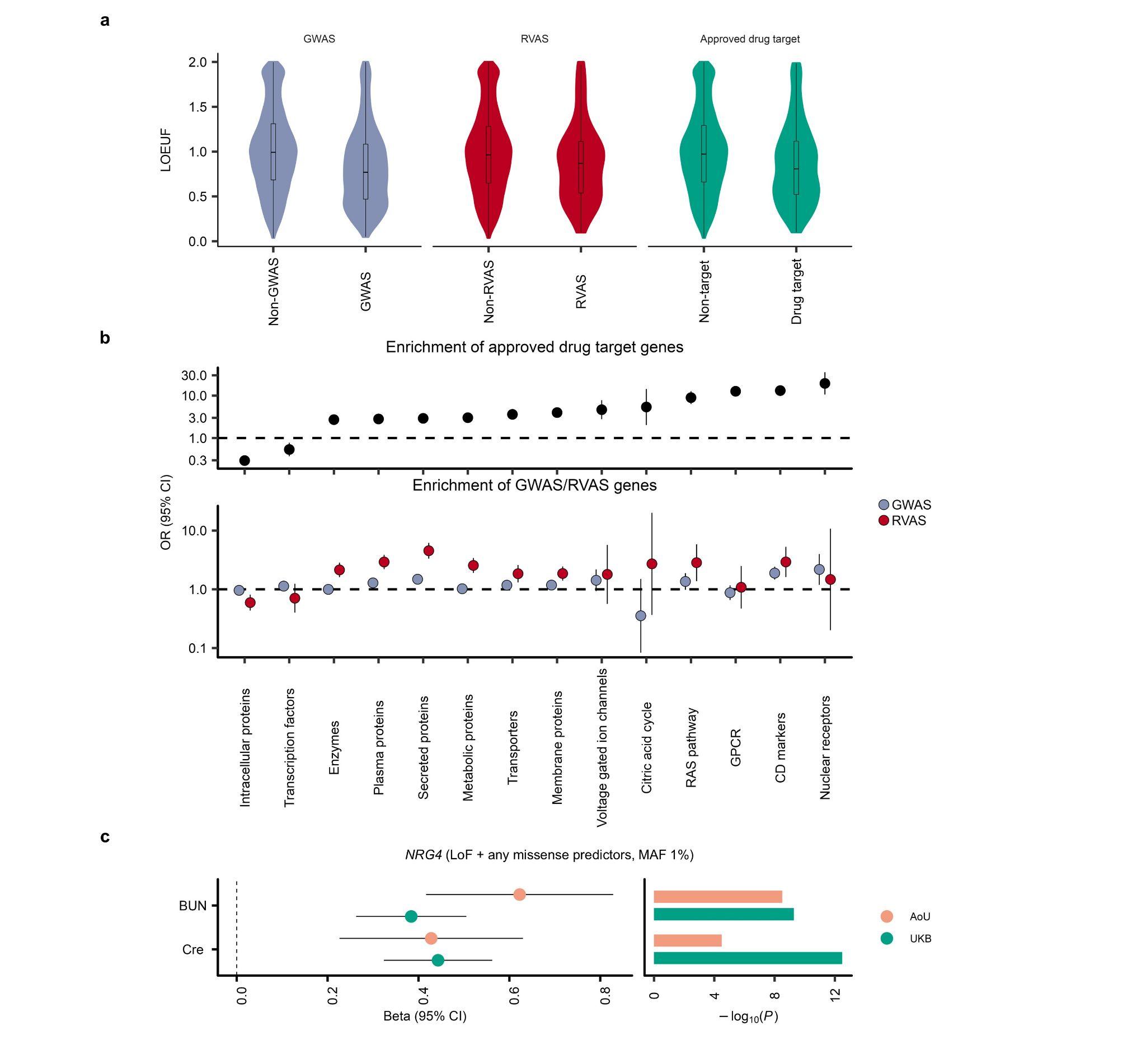

#### Supplementary Fig.15: The characteristics of GWAS genes and RVAS genes.

**a,** The difference of LOEUF among GWAS genes, RVAS genes, and approved drug target genes. The box plot shows the median value as the centerline; the box boundaries show the first and third quartiles and the whiskers extend 1.5 times the interquartile range. **b,** Top panel shows the enrichment in the approved drug target genes in the indicated protein classes. The bottom panel shows the enrichment of GWAS genes and RVAS genes in the indicated protein classes. The error bar for the effect estimates indicates the 95% CIs in **b** and **c.** **c,** Effect estimates and P values of *NRG4* (The best mask: LoF + any missense predictors, MAF 1%) on kidney-related traits among AoU and UKB. AoU, All of Us; BUN, blood urea nitrogen; CD, cluster of differentiation; CI, confidence interval; Cre, creatinine; GPCR, G protein-coupled receptor; GWAS, genome-wide association study; LOEUF, Loss-of-Function Observed/Expected Upper Bound Fraction; LoF, Loss-of-Function; MAF, minor allele frequency; OR, odds ratio; RVAS, rare variant association study; UKB, the United Kingdom Biobank.
